# Efficacy and Safety of NVX-CoV2373 in Adults in the United States and Mexico

**DOI:** 10.1101/2021.10.05.21264567

**Authors:** Lisa M. Dunkle, Karen L. Kotloff, Cynthia L. Gay, Germán Áñez, Jeffrey M. Adelglass, Alejandro Q. Barrat Hernández, Wayne L. Harper, Daniel M. Duncanson, Monica A. McArthur, Diana F. Florescu, R. Scott McClelland, Veronica Garcia-Fragoso, Robert A. Riesenberg, David B. Musante, David L. Fried, Beth E. Safirstein, Mark McKenzie, Robert J. Jeanfreau, Jeffrey K. Kingsley, Jeffrey A. Henderson, Dakotah C. Lane, Guillermo M. Ruíz-Palacios, Lawrence Corey, Kathleen M. Neuzil, Robert W. Coombs, Alex L. Greninger, Julia Hutter, Julie A. Ake, Katherine Smith, Wayne Woo, Iksung Cho, Gregory M. Glenn, Filip Dubovsky, for the 2019nCoV-301 Study Group

## Abstract

**BACKGROUND:** Vaccination using severe acute respiratory syndrome coronavirus 2 (SARS-CoV-2) spike (S) protein antigen has been effective in the prevention of coronavirus disease 2019 (Covid-19). NVX-CoV2373 is an adjuvanted, recombinant S protein nanoparticle vaccine that demonstrated clinical efficacy for prevention of Covid-19 in phase 2b/3 trials in the United Kingdom and South Africa.

**METHODS:** This phase 3, randomized, observer-blinded, placebo-controlled trial evaluated the efficacy and safety of NVX-CoV2373 in adults ≥18 years of age in the United States and Mexico during the first quarter of 2021. Participants were randomized in a 2:1 ratio to receive two doses of NVX-CoV2373 or placebo 21 days apart. The primary end point was vaccine efficacy (VE) against reverse transcriptase-polymerase chain reaction-confirmed Covid-19 in SARS-CoV-2-naïve participants ≥7 days after the second dose administration.

**RESULTS:** Of the 29,949 participants randomized between December 27, 2020, and February 18, 2021, 29,582 (median age: 47 years, 12.6% ≥65 years) received ≥1 dose: 19,714 received vaccine and 9868 placebo. In the per-protocol population, there were 77 Covid-19 cases; 14 among vaccine and 63 among placebo recipients (VE: 90.4%, 95% confidence interval [CI] 82.9 to 94.6, P<0.001). All moderate-to-severe cases occurred in placebo recipients, yielding VE of 100% (95% CI 87.0 to 100). Most sequenced viral genomes (48/61, 78.7%) were variants of concern (VOC) or interest (VOI), mainly represented by variant alpha/B.1.1.7 (31/35, 88.6% VOC identified). VE against any VOC/VOI was 92.6% (95% CI 83.6 to 96.7). Reactogenicity was mostly mild-to-moderate and transient, but more frequent in NVX-CoV2373 recipients and after the second dose. Serious adverse events were rare and evenly distributed between treatments.

**CONCLUSIONS:** NVX-CoV2373 was well tolerated and demonstrated a high overall VE (>90%) for prevention of Covid-19, with most cases due to variant strains.

(Funded by the Office of the Assistant Secretary for Preparedness and Response, Biomedical Advanced Research and Development Authority and the National Institute of Allergy and Infectious Diseases (NIAID), National Institutes of Health; PREVENT-19 ClinicalTrials.gov number, NCT04611802.)

---

The global pandemic of coronavirus disease 2019 (Covid-19) caused by the severe acute respiratory syndrome coronavirus 2 (SARS-CoV-2) has been met with an unprecedented response in vaccine development.^1^ Several Covid-19 vaccines based on different technologies are available, all of which use the SARS-CoV-2 spike (S) protein, usually based on the ancestral Wuhan strain, as an antigen.^2-5^ While these vaccines demonstrated remarkable reduction in disease burden in clinical trials, development efforts must continue to address global supply shortages and emergence of variants that impact virulence or susceptibility to vaccine-induced immunity (variants of concern, VOC, or variants of interest, VOI).^6-8^

NVX-CoV2373, a SARS-CoV-2 vaccine comprising full-length, stabilized pre-fusion, recombinant S (rS) protein trimers produced from the Wuhan-Hu-1 sequence, is assembled into nanoparticles co-formulated with a saponin-based adjuvant (Matrix-M™). The vaccine, stable at refrigerated temperature (2-8°C), represents an important product to address the continually evolving Covid-19 pandemic and the concurrent global vaccine shortage.^9^ NVX-CoV2373 offers the advantage of representing a traditional adjuvanted protein subunit-based platform that may reduce vaccine hesitancy and extend the durability of immune response.^10^

NVX-CoV2373 was well tolerated and immunogenic in adults^10,11^ and provided high vaccine efficacy (VE) against severe disease caused by VOC beta/B.1.351 in a phase 2b trial in South Africa,^12^ and against Covid-19 of any severity from VOC alpha/B.1.1.7 in a phase 3 trial in the United Kingdom (UK).^13^ We describe the results of PREVENT-19, a phase 3 trial of NVX-CoV2373 in adults ≥18 years of age in the United States and Mexico conducted during a period of predominantly VOC/VOI circulation.

## Methods

### TRIAL DESIGN, PARTICIPANTS, PROCEDURES, AND OVERSIGHT

A phase 3, randomized, observer-blinded, placebo-controlled trial conducted in 113 clinical sites in the United States and six in Mexico evaluated efficacy and safety of NVX-CoV2373. Participants received initial injections between December 27, 2020, and February 18, 2021, and were followed through April 19, 2021. Healthy adults ≥18 years of age or those with stable chronic medical conditions, including chronic pulmonary, renal, or cardiovascular disease, diabetes mellitus type 2, or well-controlled human immunodeficiency virus (HIV) infection, were eligible for participation. Key exclusion criteria included known previous laboratory-confirmed SARS-CoV-2 infection or known immunosuppression. Consistent with the US Food and Drug Administration (FDA) and funding agency guidance,^1,14^ enrollment targets were specified for inclusion of racial and ethnic minorities, and site selection prioritized accessibility to participants at high risk of acquisition or complications of Covid-19. Additional details regarding trial design, conduct, oversight, and analyses are provided in the Supplementary Appendix, the protocol, and statistical analysis plan.

Participants provided written informed consent before enrollment, were stratified by age (18 to ≤64 or ≥65 years), and randomized using a web-based interactive response system in a 2:1 ratio to receive two 0.5-mL intramuscular injections of either NVX-CoV2373 (5 μg rS SARS-CoV-2 vaccine adjuvanted with 50 μg Matrix-M™) or placebo 21 days apart. Only unblinded site personnel managed study vaccine logistics/preparation and had no other role in trial conduct.

Novavax was the trial sponsor and responsible for its design, development, and manufacture of NVX-CoV2373 clinical trial material. PREVENT-19 was part of the US Government-funded harmonized trial program. Site selection, monitoring, data collection and analysis, and preparation of the manuscript were conducted in collaboration with the Biomedical Advanced Research and Development Authority, the National Institute of Allergy and Infectious Diseases/National Institutes of Health (NIAID/NIH), the Covid-19 Prevention Network, and trial co-chairs.^15^ Trial data were available to all authors, who vouched for its accuracy and completeness and for fidelity to the trial protocol. The trial is ongoing, and investigators and Novavax clinical team remain blinded to participant-level treatment assignments. The protocol, amendments, and overall oversight were provided by institutional review boards/ethics committees according to local regulations. Safety oversight was provided by a NIAID/NIH-sponsored Protocol Safety Review Team, and safety, efficacy, and potential vaccine-induced harm were monitored through regular review of unblinded data by the NIAID/NIH-sponsored Data and Safety Monitoring Board (DSMB).^16^

### EFFICACY ASSESSMENTS

The primary end point was the efficacy of NVX-CoV2373 in preventing the first episode of reverse transcriptase-polymerase chain reaction (RT-PCR)-confirmed symptomatic mild, moderate, or severe Covid-19 (using FDA criteria,^14^ Table S3) with onset ≥7 days after the second injection in the per-protocol population of efficacy (PP-EFF). Symptoms of suspected Covid-19 (Table S2) were reported daily by participants using an electronic diary. When specified symptoms were reported for ≥2 consecutive days, participants were instructed to self-collect three daily nasal swabs and undergo in-clinic medical evaluation, including nasal swab collection for SARS-CoV-2 testing at a central laboratory. Confirmation of end point Covid-19 cases was achieved when ≥1 nasal swab was positive by SARS-CoV-2 RT-PCR at the central laboratory. Whole-genome sequencing (WGS) and clade/lineage assignment were performed on RT-PCR-positive samples with sufficient viral RNA load (Supplementary Appendix).

The key secondary efficacy end point was protection against RT-PCR-confirmed Covid-19 due to non-VOC/VOI strains, i.e., circulating strains (D614G) not containing VOC/VOI-defining mutations and considered most clinically similar to the ancestral Wuhan-Hu-1 strain. Other secondary end points included VE against moderate-to-severe Covid-19 and VE assessed across subgroups (race, ethnicity, age, and comorbidities associated with increased risk of severe Covid-19 according to Centers for Disease Control and Prevention [CDC] criteria,^17^ or risk of exposure related to occupation/lifestyle), and safety and reactogenicity. Exploratory efficacy end points included protection against RT-PCR-confirmed Covid-19 due to a VOC/VOI, as defined by CDC.^6^ Assessment of Covid-19 severity as an end point was evaluated by investigators and study physicians, and severe cases were confirmed by an external independent End Point Review Committee blinded to treatment assignment.

### SAFETY ASSESSMENTS

Solicited local and systemic adverse events (AEs) were collected via electronic diary for 7 days following each injection. Participants were assessed for unsolicited AEs from the first dose through 28 days after the second dose; serious adverse events (SAEs), adverse events of special interest (AESIs), and medically attended adverse events (MAAEs) were assessed from the first dose through data cutoff.

### STATISTICAL ANALYSIS

#### Efficacy analysis

The PP-EFF used for all efficacy analyses included participants who had no evidence of past SARS-CoV-2 infection at baseline until ≥7 days after the second injection (i.e., negative serum anti-SARS-CoV-2 nucleoprotein and nasal swab negative by SARS-CoV-2 RT-PCR), who received both injections of assigned treatment, and had no major protocol deviations. Vaccine efficacy (VE%) was defined as (1 – RR) × 100, where RR is the relative risk of incidence rates between the two treatment groups. The estimated RR and two-sided 95% confidence interval (CI) were derived using Poisson regression with robust error variance. The pre-specified success criteria for the primary efficacy end point were defined as lower bound of two-sided 95% CI of VE >30% and point estimate of VE ≥50%.^14^

The trial was designed to evaluate efficacy based on the number of events expected to be required to achieve statistical significance for the primary outcome, originally estimated to be 144 primary end points. However, to maintain the viability of the trial and retain participants in the face of the nationwide vaccination campaign using vaccines under Emergency Use Authorization (EUA), a blinded crossover (participants originally randomized to placebo were administered NVX-CoV2373 and vice versa) was implemented, coinciding with the accumulation of a median 2-month safety follow-up, to ensure that all participants received active vaccine at the earliest time possible without compromising FDA-required placebo-controlled safety follow-up. The final analysis of the placebo-controlled portion of the study, approved by the DSMB prior to general access to unblinded data and reported here, includes the number of end point cases accrued prior to the crossover. As a result of the blinded crossover, a single efficacy analysis was conducted using the full two-sided type I error of 5% for the primary end point.

#### Safety analysis

Safety data from all participants who received at least one dose of study treatment was summarized descriptively. Solicited local and systemic AEs were assessed for severity using FDA criteria^18^ and duration after each injection. Unsolicited AEs were coded by preferred term and system organ class using the Medical Dictionary for Regulatory Activities (MedDRA), version 23.1, and summarized by severity and relationship to study vaccine.

## Results

### PARTICIPANTS

Of 31,588 participants screened, 29,949 were randomized between December 27, 2020, and February 18, 2021 (Figure 1). Among 29,582 participants who received at least one dose of NVX-CoV2373 (19,714) or placebo (9868) in the full analysis set (FAS), 25,452 (17,312 NVX-CoV2373 recipients and 8140 placebo recipients) met criteria for inclusion in the PP-EFF. Baseline demographics of the PP-EFF were well balanced between treatment groups; 48.2% self-identified as female, 75.9% as White, 11.0% as Black/African American, 6.2% as Native American/Alaska Native (including American Indians and Native Mexicans), 21.5% as Hispanic/Latino. One or more comorbid conditions^17^ were reported by 47.3%. The median age was 47 years and 11.8% were ≥65 years of age (Table 1). Baseline seropositivity/RT-PCR positivity rates in the FAS were 6.3% and 6.9% among NVX-CoV2373 and placebo recipients, respectively (Table S4).

**Figure 1.**
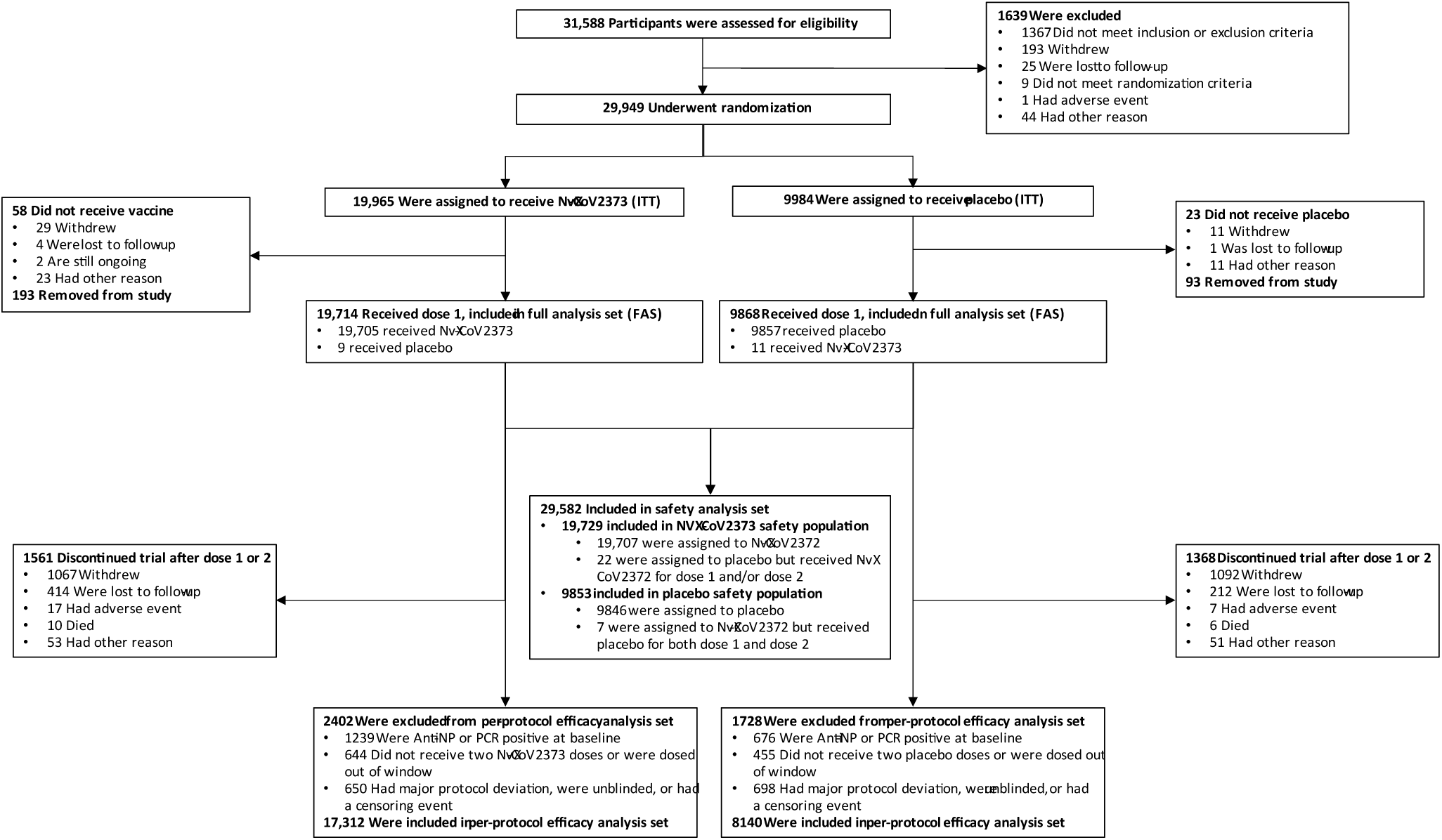
Trial Disposition. The full analysis set (FAS) included all participants who were randomly assigned to treatment and received at least one dose, regardless of protocol violations or missing data, and are analyzed according to the trial vaccine group as randomized.

**Table 1.**
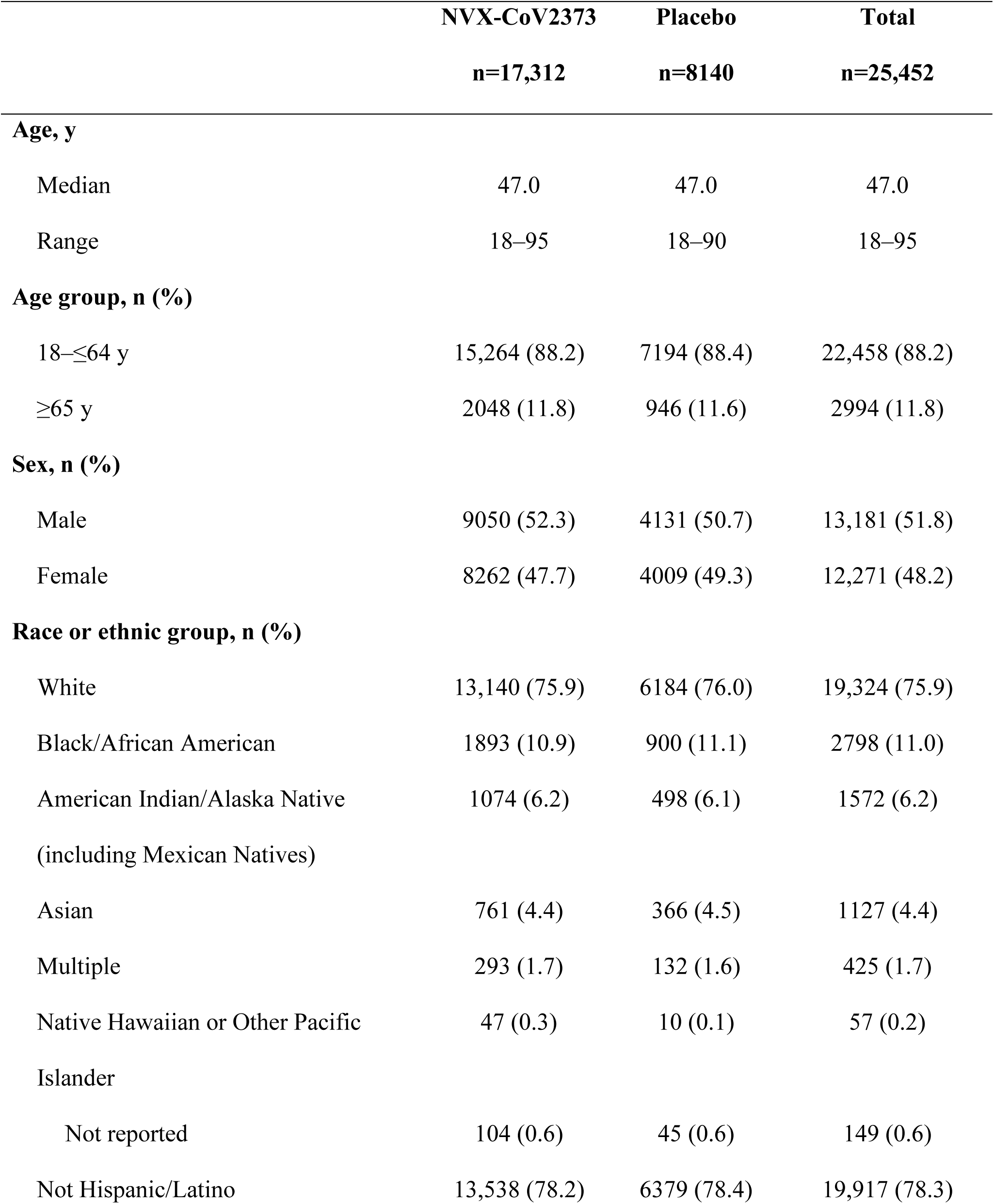
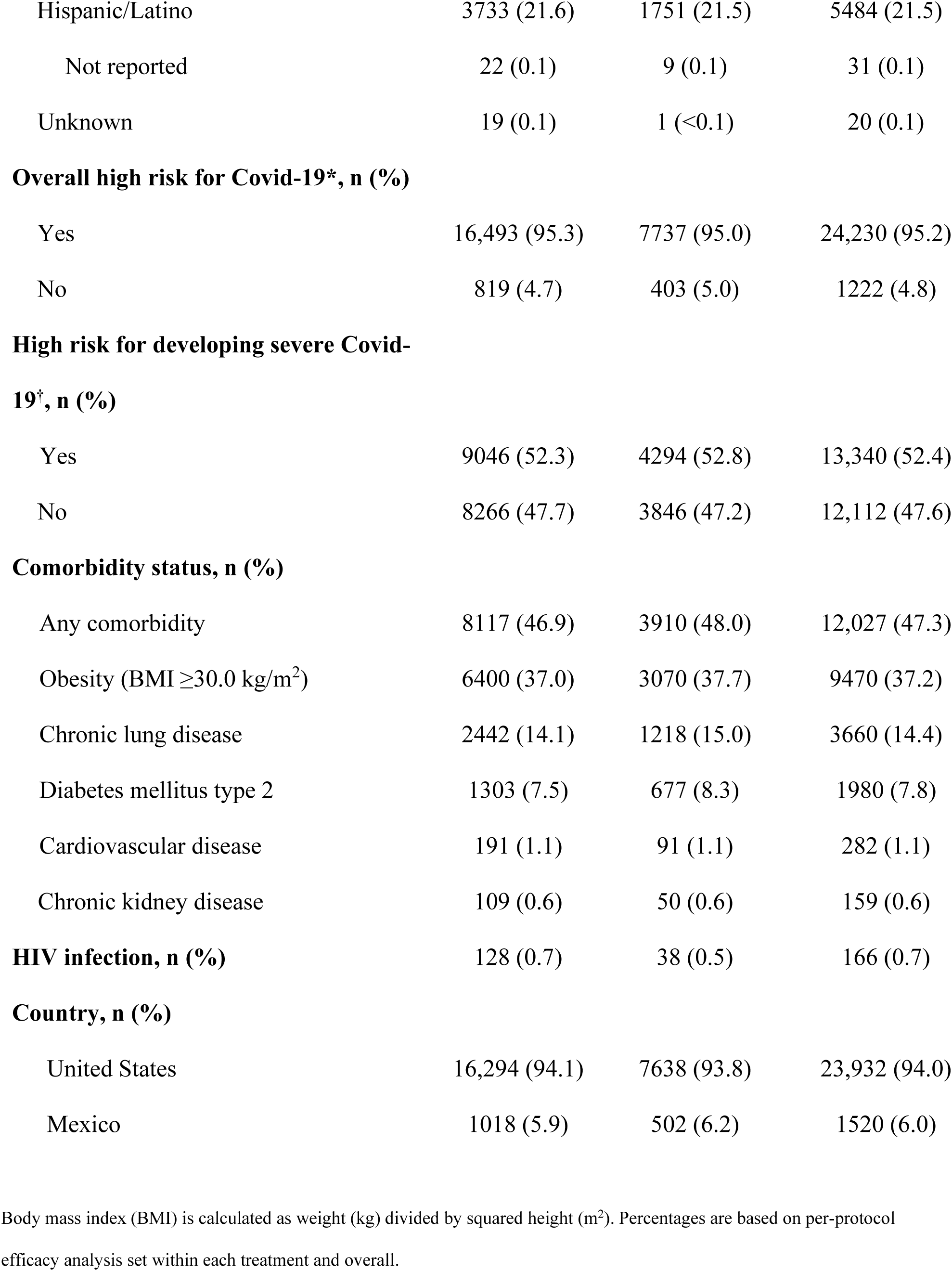
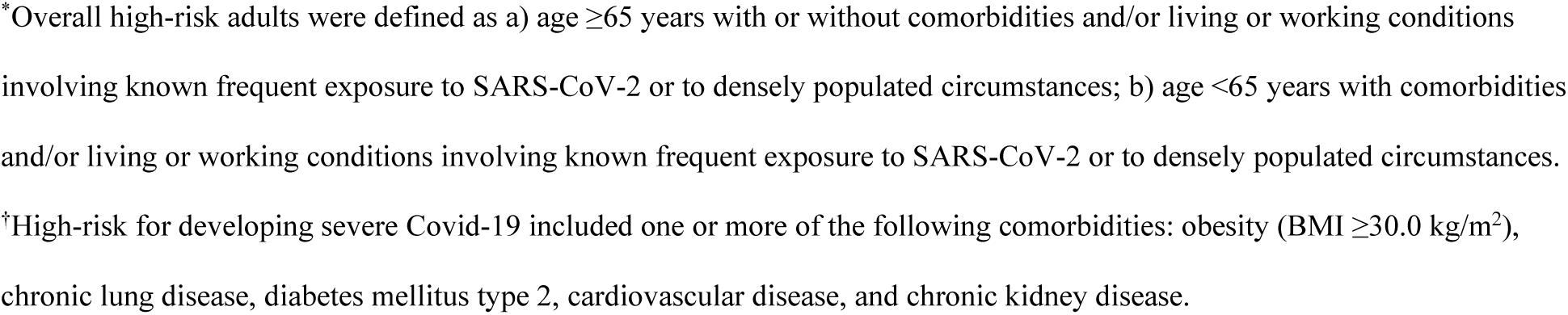
Demographics and baseline characteristics (per-protocol efficacy population)

|  | **NVX-CoV2373** | **Placebo** | **Total** |
| --- | --- | --- | --- |
|  | **n=17,312** | **n=8140** | **n=25,452** |
| **Age, y** |  |  |  |
| Median | 47.0 | 47.0 | 47.0 |
| Range | 18–95 | 18–90 | 18–95 |
| **Age group, n (%)** |  |  |  |
| 18–≤64 y | 15,264 (88.2) | 7194 (88.4) | 22,458 (88.2) |
| ≥65 y | 2048 (11.8) | 946 (11.6) | 2994 (11.8) |
| **Sex, n (%)** |  |  |  |
| Male | 9050 (52.3) | 4131 (50.7) | 13,181 (51.8) |
| Female | 8262 (47.7) | 4009 (49.3) | 12,271 (48.2) |
| **Race or ethnic group, n (%)** |  |  |  |
| White | 13,140 (75.9) | 6184 (76.0) | 19,324 (75.9) |
| Black/African American | 1893 (10.9) | 900 (11.1) | 2798 (11.0) |
| American Indian/Alaska Native<br>(including Mexican Natives) | 1074 (6.2) | 498 (6.1) | 1572 (6.2) |
| Asian | 761 (4.4) | 366 (4.5) | 1127 (4.4) |
| Multiple | 293 (1.7) | 132 (1.6) | 425 (1.7) |
| Native Hawaiian or Other Pacific<br>Islander | 47 (0.3) | 10 (0.1) | 57 (0.2) |
| Not reported | 104 (0.6) | 45 (0.6) | 149 (0.6) |
| Not Hispanic/Latino | 13,538 (78.2) | 6379 (78.4) | 19,917 (78.3) |
| Hispanic/Latino | 3733 (21.6) | 1751 (21.5) | 5484 (21.5) |
| Not reported | 22 (0.1) | 9 (0.1) | 31 (0.1) |
| Unknown | 19 (0.1) | 1 (<0.1) | 20 (0.1) |
| **Overall high risk for Covid-19\*, n (%)** |  |  |  |
| Yes | 16,493 (95.3) | 7737 (95.0) | 24,230 (95.2) |
| No | 819 (4.7) | 403 (5.0) | 1222 (4.8) |
| **High risk for developing severe Covid-19†, n (%)** |  |  |  |
| Yes | 9046 (52.3) | 4294 (52.8) | 13,340 (52.4) |
| No | 8266 (47.7) | 3846 (47.2) | 12,112 (47.6) |
| **Comorbidity status, n (%)** |  |  |  |
| Any comorbidity | 8117 (46.9) | 3910 (48.0) | 12,027 (47.3) |
| Obesity (BMI $\geq 30.0$ kg/m<sup>2</sup>) | 6400 (37.0) | 3070 (37.7) | 9470 (37.2) |
| Chronic lung disease | 2442 (14.1) | 1218 (15.0) | 3660 (14.4) |
| Diabetes mellitus type 2 | 1303 (7.5) | 677 (8.3) | 1980 (7.8) |
| Cardiovascular disease | 191 (1.1) | 91 (1.1) | 282 (1.1) |
| Chronic kidney disease | 109 (0.6) | 50 (0.6) | 159 (0.6) |
| **HIV infection, n (%)** | 128 (0.7) | 38 (0.5) | 166 (0.7) |
| **Country, n (%)** |  |  |  |
| United States | 16,294 (94.1) | 7638 (93.8) | 23,932 (94.0) |
| Mexico | 1018 (5.9) | 502 (6.2) | 1520 (6.0) |

Body mass index (BMI) is calculated as weight (kg) divided by squared height (m<sup>2</sup>). Percentages are based on per-protocol efficacy analysis set within each treatment and overall.
\*Overall high-risk adults were defined as a) age $\geq 65$ years with or without comorbidities and/or living or working conditions involving known frequent exposure to SARS-CoV-2 or to densely populated circumstances; b) age $< 65$ years with comorbidities and/or living or working conditions involving known frequent exposure to SARS-CoV-2 or to densely populated circumstances.
<sup>†</sup>High-risk for developing severe Covid-19 included one or more of the following comorbidities: obesity (BMI $\geq 30.0$ kg/m<sup>2</sup>), chronic lung disease, diabetes mellitus type 2, cardiovascular disease, and chronic kidney disease.

### EFFICACY

In the FAS, the incidence of Covid-19 in the placebo group was 51.9 cases per 1000 person-years (95% CI 40.9 to 66.0), and cumulative incidence curves began separating around day 21 (Figure 2A). Among the 25,452 PP-EFF participants followed through April 19, 2021, 77 Covid-19 cases occurred (incidence 34.0 cases per 1000 person-years [95% CI 20.7 to 55.9] in placebo and 3.3 cases per 1000 person-years [95% CI 1.6 to 6.9] in NVX-CoV2373 recipients). The 14 cases in NVX-CoV2373 recipients and 63 in placebo recipients yielded VE of 90.4% (95% CI, 82.9 to 94.6) (Table S5). Cumulative incidence in NVX-CoV2373 recipients was higher during the first 42 days of follow-up, after which the incidence declined while continuing to increase in placebo recipients (Figure 2B).

**Figure 2.**
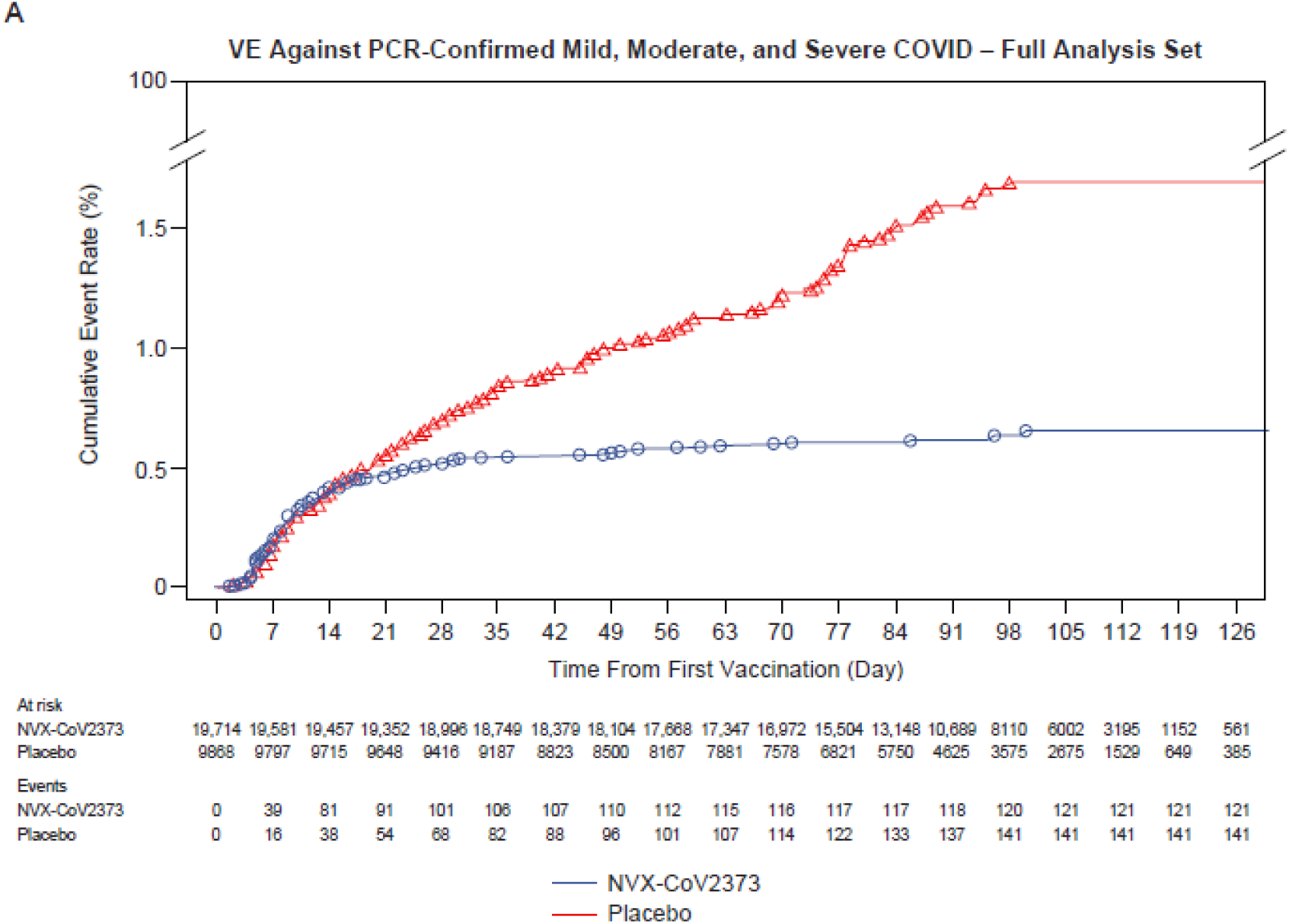
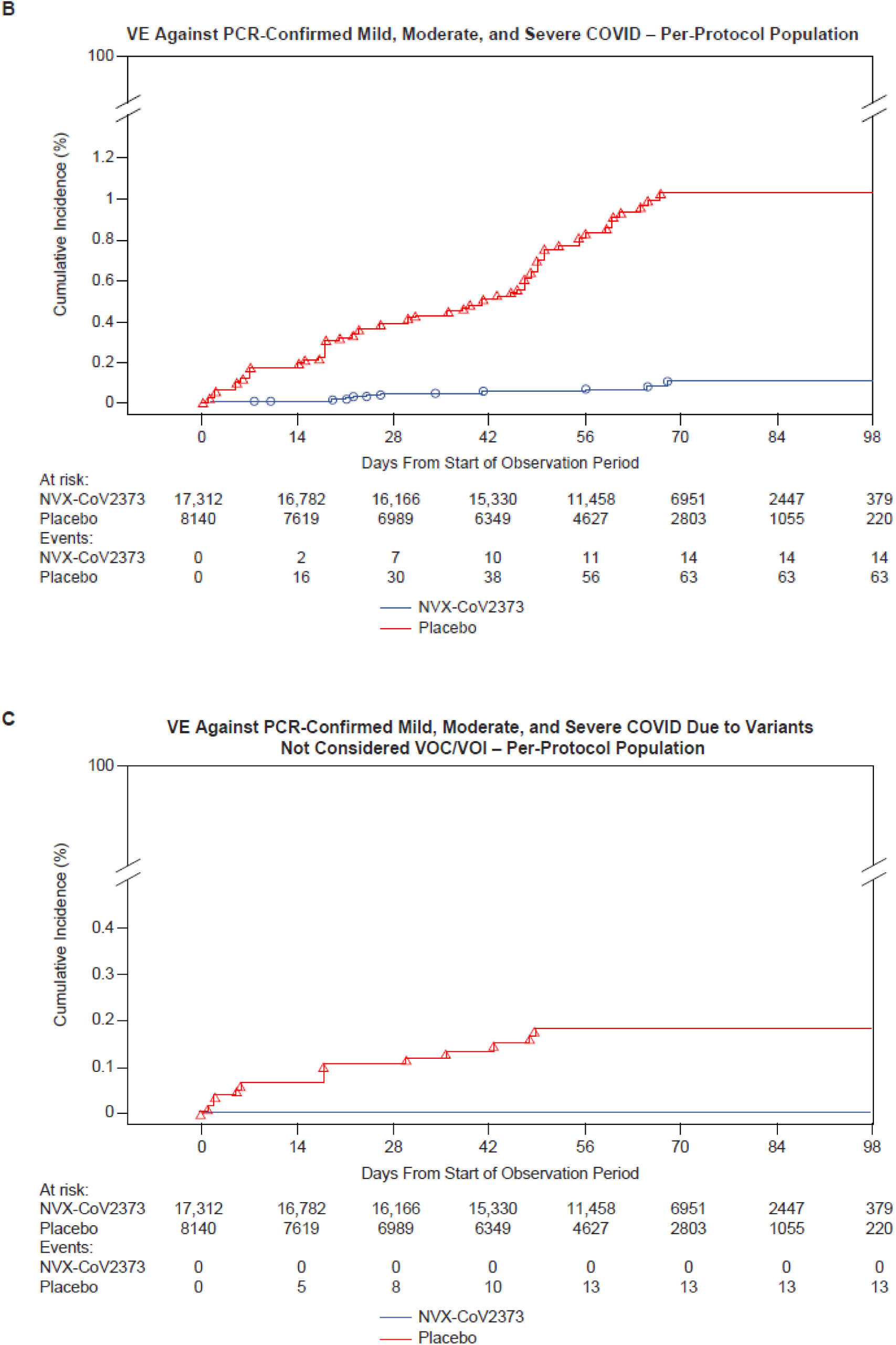
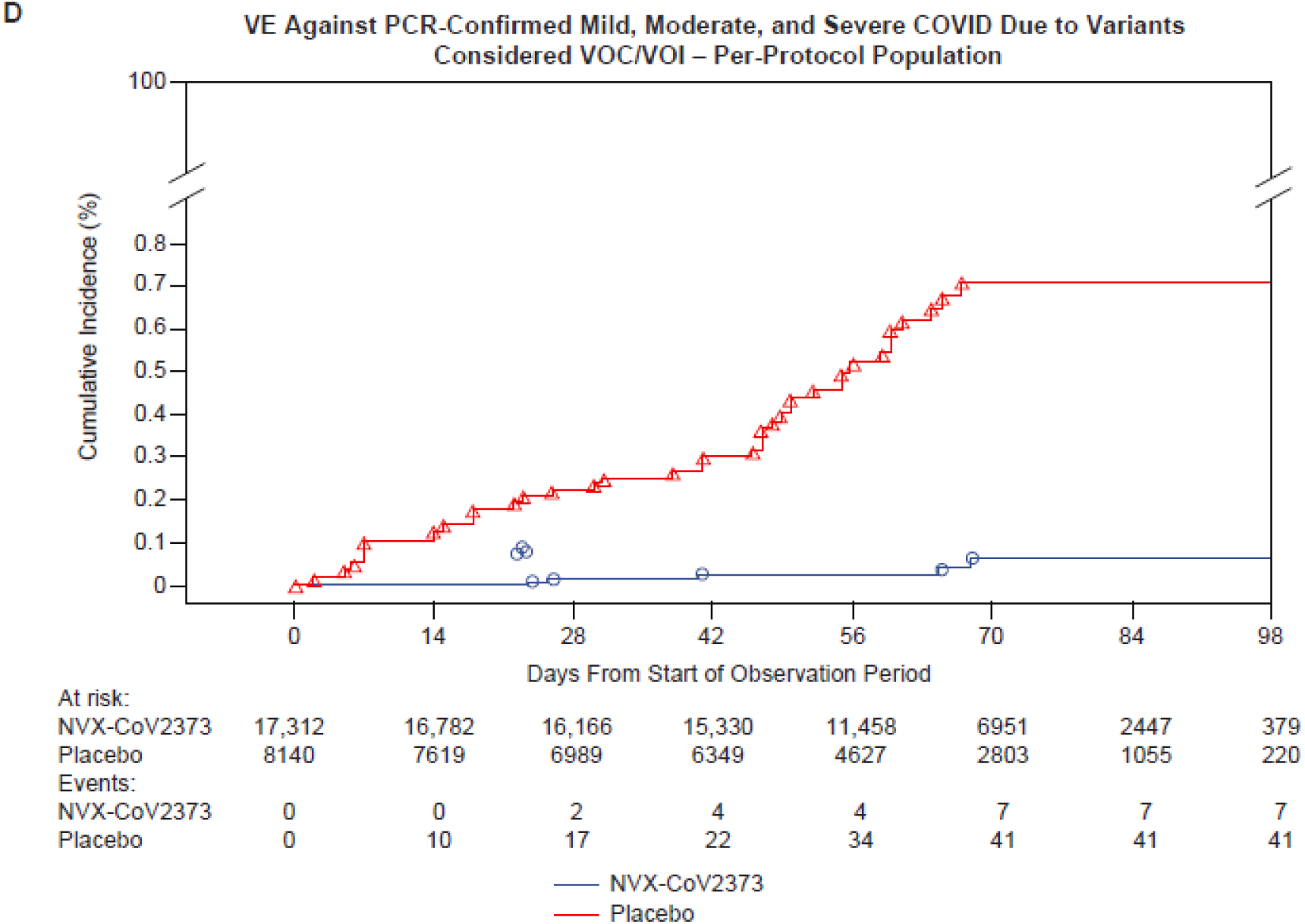
Cumulative Incidence Plots of Overall Efficacy of NVX-CoV2373 Against Symptomatic Covid-19. Shown is the cumulative incidence of symptomatic Covid-19. The time period for surveillance of the full analysis population was from the first dose of NVX-CoV2373 or placebo; per-protocol symptomatic Covid-19 cases were from at least 7 days after the second dose (i.e., Day 28) through approximately 4 months of follow-up, or unblinding or receipt of EUA vaccine. A) All participants, baseline seronegative/virologically negative, full analysis set; B) All participants, baseline seronegative/virologically negative, per-protocol population; C) All participants, baseline seronegative/virologically negative, per-protocol population with end points due to variants not considered VOC/VOI; and D) All participants, baseline seronegative/virologically negative, per-protocol population with end points due to variants considered VOC/VOI.

All cases reported in NVX-CoV2373 recipients were mild in severity, and all 14 moderate-to-severe cases occurred in the placebo group, yielding VE against moderate-to-severe Covid-19 of 100% (95% CI, 87 to 100) (Figure 3). VE against severe Covid-19 alone (N=4) was 100% (95% CI, 34.6 to 100, *post-hoc* analysis). VE among participants at high-risk for acquisition and/or complications was 91.0% (95% CI, 83.6 to 95.0). VE in several prespecified demographic subgroups demonstrated generally similar high degrees of efficacy. Hispanics/Latinos were the only demographic group with a lower VE (67.3%; 95% CI, 18.7 to 86.8) compared to other groups (Figure 3).

**Figure 3.**
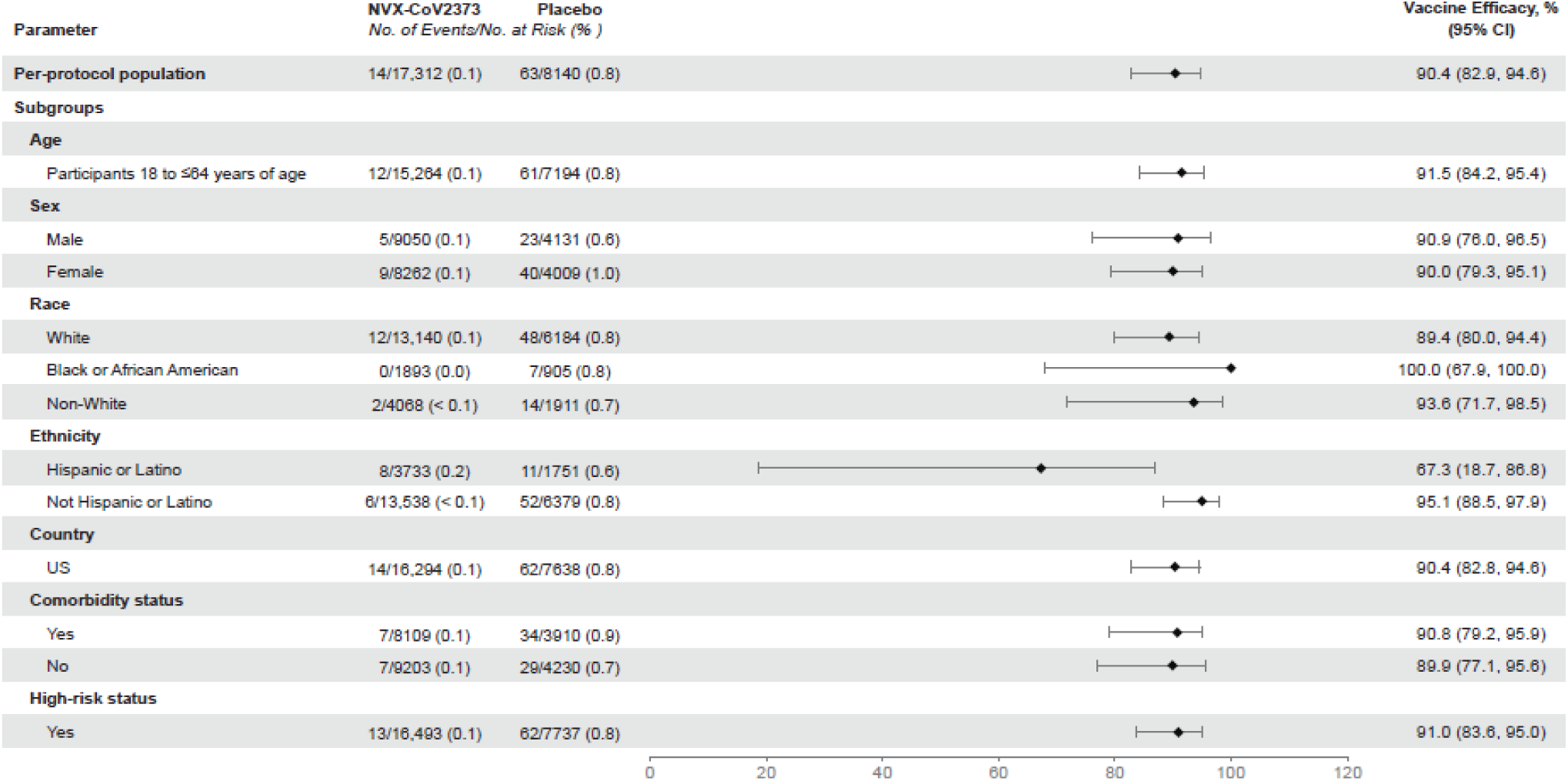
Vaccine Efficacy of NVX-CoV2373 in Specific Subgroups in the Per-Protocol Population. Vaccine efficacy was defined as 1 minus the relative risk (NVX-CoV2373 vs. placebo). “Non-White” race included end point cases from all other races to ensure that these subpopulations would be large enough for meaningful analyses. Comorbidity assessment is based on the Centers for Disease Control and Prevention definitions^17^ of those at increased risk for Covid-19. Overall high-risk participants include those ≥65 years of age, with chronic health conditions, or at increased risk of Covid-19 due to work or living conditions.

Nasal swabs from 61/77 (79.2%) end point cases yielded WGS. Of these, 35 VOC, 13 VOI, and 13 non-VOC/VOI were identified (Figure S1). Most VOC (31/35, 88.6%) were identified as variant alpha. VE against non-VOC/VOI was 100% (95% CI, 85.8 to 100) (Figure 2C), against any VOC/VOI was 92.6% (95% CI, 83.6 to 96.7) (Figure 2D), and against VOC alpha was 93.6% (95% CI, 81.7 to 97.8, *post-hoc* analysis).

### SAFETY

#### Reactogenicity

Solicited local and systemic AEs were predominantly mild-to-moderate and transient, but more frequent in NVX-CoV2373 than in placebo recipients, and more so after the second injection. After each dose, the most frequently reported solicited local AEs were tenderness and injection site pain. The median duration of these events was ≤2 days (Table S11). Severe (≥Grade 3) local reactions were infrequent (1.1% vs. <1% after dose 1, and 6.7% vs. <1% after dose 2 in the NVX-CoV2373 and placebo groups, respectively) (Figure 4 and Table S10).

**Figure 4.**
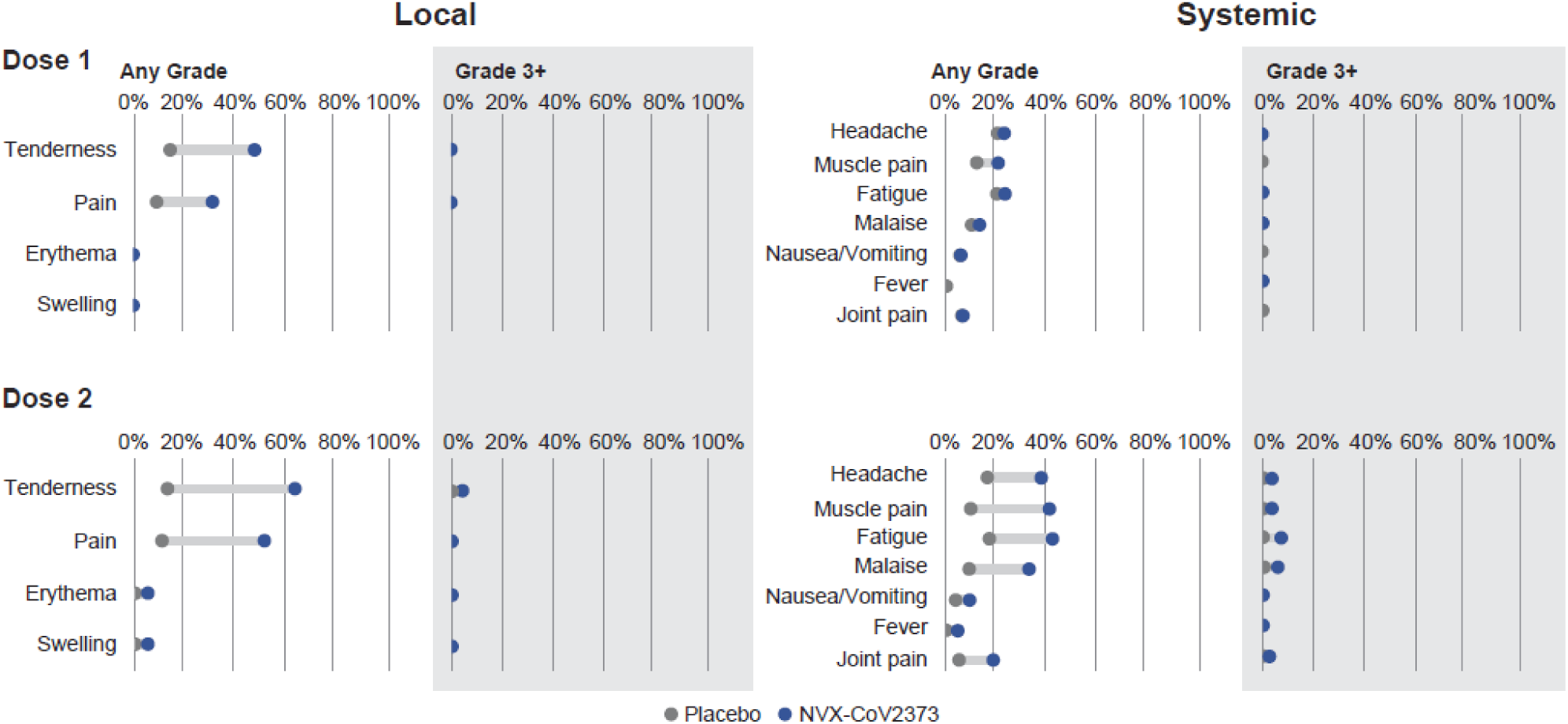
Solicited Local and Systemic Adverse Events. The percentage of participants in each treatment group with solicited local and systemic adverse events during the 7 days after each vaccination is plotted by FDA toxicity grade, either as any grade (mild, moderate, severe, or potentially life-threatening) or as Grade 3+ (severe or potentially life-threatening).^18^

The most common solicited systemic AEs: headache, myalgia, fatigue, and malaise were detected more frequently among NVX-CoV2373 recipients and after the second injection with a median duration of ≤1 day (Table S13). Fever of any severity was rare (<1%) and similarly distributed among vaccine and placebo groups after each dose. Severe systemic reactions (≥Grade 3) were infrequent (2.4% vs. 2.1% after dose 1, and 12.1% vs. 2.1% after dose 2 in the NVX-CoV2373 and placebo groups, respectively) (Figure 4 and Table S12).

#### Unsolicited AEs

Unsolicited AEs were slightly more frequent in vaccine than in placebo recipients (16.3% vs. 14.8%), although the imbalance appeared to include duplicate reporting of reactogenicity. There was a balanced frequency of MAAEs, SAEs, severe AEs, AESI related to Covid-19, and potential immune-mediated medical conditions between treatment groups (Table S9). There were no episodes of anaphylaxis, no evidence of vaccine-associated enhanced Covid-19, and no events that triggered prespecified pause rules. No episodes of Guillain Barré syndrome^19^ and no imbalance in myocarditis/pericarditis^20^ or thrombosis with thrombocytopenia syndrome (TTS)^21^ were observed (Tables S14-S16). All-cause mortality was balanced, nine (0.5%) among NVX-CoV2373 recipients and five (0.5%) in placebo recipients.

## Discussion

PREVENT-19 is an ongoing randomized, controlled trial assessing the efficacy of an adjuvanted rS SARS-CoV-2 vaccine in ≈30,000 participants. The trial includes a demographically diverse population in the United States and Mexico and provides strong evidence of high short-term VE of NVX-CoV2373 for the prevention of Covid-19 (>90%) and for prevention of moderate-to-severe disease (100%).

NVX-CoV2373 displayed a favorable safety profile within a median safety follow-up of 2 months, characterized by mild-to-moderate, transient reactogenicity. No safety concerns related to vaccination were seen, as demonstrated by comparable rates of unsolicited AEs (including SAE/severe AEs) between vaccine and placebo recipients. None of the safety signals under observation with other Covid-19 vaccines^19-21^ have been reported in PREVENT-19. Long-term safety monitoring is planned to continue throughout 24 months after initial vaccination.

PREVENT-19 expands observations from previous NVX-CoV2373 trials evaluating its efficacy in the evolving pandemic. SARS-CoV-2 has undergone extensive genetic evolution resulting in emergence of multiple variants that may cause evasion from S protein-based vaccine-induced immunity.^6-8^ NVX-CoV2373 showed VE >90% against VOC/VOI circulating during case accrual in the United States and Mexico, largely represented by the alpha VOC. Similar protection against this VOC was observed in the earlier phase 3 trial in the UK^12^ (93.6% in PREVENT-19 and 86.3% in the UK trial). Moreover, the efficacy against the alpha VOC in both trials was similar to the VE against the 13 non-VOC/VOI cases identified in PREVENT-19 (i.e., variants that may more closely resemble the ancestral Wuhan-Hu-1 S sequence from which the vaccine antigen was derived), suggesting that the vaccine induces protective immunity to a broader spectrum of variants beyond the prototype strain.^12^

Some additional strengths of the PREVENT-19 trial include demographic diversity, which permitted an assessment of VE across racial and ethnic minorities reflecting the US and Mexican populations particularly impacted by the pandemic. VE among Hispanics/Latinos in the United States was lower than in other demographic groups, which could be related to chance alone or to unidentified viral or host factors. However, all end point cases in Hispanic/Latino NVX-CoV2373-recipients were mild in severity.

Several limitations of this study are noteworthy. EUA vaccines became available concurrently with PREVENT-19’s initiation, reducing elderly participant enrollment, resulting in only four end point cases in participants ≥65 years of age and preventing a meaningful estimation of VE. However, the UK phase 3 trial enrolled enough older adults to establish a VE of 88.9%, comparable to younger adults.^13^

PREVENT-19 was the first to implement a blinded crossover ≈3-4 months after the first vaccination series to allow all trial participants to receive NVX-CoV2373, once VE and required safety were established and reviewed by the DSMB. To address durability of VE and long-term safety after blinded crossover, hazard models have been proposed for subsequent analyses,^22^ which would allow comparisons between early- and later-vaccinated groups.

Lastly, as with all major Covid-19 vaccine trials, VE was assessed over a relatively short time during a rapidly evolving pandemic. Initial mRNA vaccine trials in the United States^2,3^ tested VE largely against viral variants not significantly different from the source vaccine antigen, demonstrating a high degree of VE against Covid-19 of any severity and against severe disease. The VE results from PREVENT-19 not only recapitulate the overall high VE observed during those trials, but additionally demonstrated high VE regardless of causal strain; 100% against non-VOC/VOI and 92.6% against VOC/VOI predominating during the case-accruing period.

NVX-CoV2373 represents a new platform that can be added to the portfolio of vaccines that are safe, well tolerated, and highly protective against contemporary SARS-CoV-2 strains. The extended stability and easy storage requirements (up to 6 months at refrigerator temperatures) make it ideally suited for global deployment. Further, the conventional and established adjuvanted protein-based formulation may address vaccine hesitancy concerns related to newer vaccine platforms.

The significant efficacy of NVX-CoV2373 in preventing moderate-to-severe disease as well as overall Covid-19 in people at high risk for Covid-19 acquisition and complications will make this vaccine a critical addition in control of the pandemic and its most serious health and economic consequences.

## Data Availability

Data Sharing Statement will be available with the full text of this article upon publication.

https://clinicaltrials.gov/ct2/show/NCT04611802?cond=NCT04611802&draw=2&rank=1

## Disclosures

Supported by Novavax, Inc., the Office of the Assistant Secretary for Preparedness and Response, Biomedical Advanced Research and Development Authority (BARDA) (contract number: OWS: Novavax’s Project Agreement No. 1 under its Medical CBRN Defense Consortium (MCDC) Base Agreement No. 2020-530; Department of Defense (DoD) No. W911QY20C0077) and the National Institute of Allergy and Infectious Diseases (NIAID); HIV Vaccine Trials Network (HVTN) Leadership and Operations Center (UM1 AI68614), the HVTN Statistics and Data Management Center (UM1 AI68635), the HVTN Laboratory Center (UM1 AI68618), the HIV Prevention Trials Network Leadership and Operations Center (UM1 AI68619), the AIDS Clinical Trials Group Leadership and Operations Center (UM1 AI68636), and the Infectious Diseases Clinical Research Consortium leadership is group (UM1 AI148684). No other potential conflict of interest relevant to this article was reported.

Disclosure forms provided by the authors will be available with the full text of this article upon publication.

A data-sharing statement provided by the authors will be available with the full text of this article upon publication.

## Acknowledgments

We especially thank all the study participants who volunteered for the study and who have contributed their clinical experience to the establishment of safety and efficacy of NVX-CoV-2373, as well as to the 2019nCoV-301 Study Group members (listed in the Supplementary Appendix); to the funders (BARDA, NIAID/NIH), the members of the NIAID Data and Safety Monitoring Board (DSMB), and the Protocol Safety Review Team (PSRT), whose diligent monitoring of all trial data contributed to ensure the safety and well-being of the trial participants; and NIAID/NIH colleagues who provided valuable infrastructure support for the NIAID-affiliated investigative sites; community leadership groups throughout the country who assisted with community engagement and recruitment; and unnamed colleagues at each of the sites who generously contributed to the trials in many ways, and all unnamed colleagues at Novavax, Inc., who worked tirelessly and gave unlimited efforts to the development, testing, and support of this trial. Editorial assistance on the preparation of this manuscript was provided by Phase Five Communications, supported by Novavax, Inc.

## References

1. Slaoui M, Hepburn M. Developing safe and effective covid vaccines — Operation Warp Speed’s strategy and approach. N Engl J Med 2020;383:1701–3.

2. Baden LR, El Sahly HM, Essink B, et al. Efficacy and safety of the mRNA-1273 SARS-CoV-2 vaccine. N Engl J Med 2021;384:403–16.

3. Polack FP, Thomas SJ, Kitchin N, et al. Safety and efficacy of the BNT162b2 mRNA Covid-19 vaccine. N Engl J Med 2020;383:2603–15.

4. Logunov DY, Dolzhikova IV, Shcheblyakov DV, et al. Safety and efficacy of an rAd26 and rAd5 vector-based heterologous prime-boost COVID-19 vaccine: an interim analysis of a randomised controlled phase 3 trial in Russia. Lancet 2021;397:671–81.

5. Voysey M, Clemens SAC, Madhi SA, et al. Safety and efficacy of the ChAdOx1 nCoV-19 vaccine (AZD1222) against SARS-CoV-2: an interim analysis of four randomised controlled trials in Brazil, South Africa, and the UK. Lancet 2021;397:99–111.

6. US Centers for Disease Control and Prevention. SARS-CoV-2 Variant Classifications and Definitions (https://www.cdc.gov/coronavirus/2019-ncov/variants/variant-info.html).

7. Harvey WT, Carabelli AM, Jackson B, et al. SARS-CoV-2 variants, spike mutations and immune escape. Nature Reviews Microbiology 2021;19:409–24.

8. Krause PR, Fleming TR, Longini IM, et al. SARS-CoV-2 Variants and Vaccines. N Engl J Med 2021;385:179–86.

9. Holder J. Tracking Coronavirus Vaccinations Around the World. The New York Times. September 30, 2021. (https://www.nytimes.com/interactive/2fid021/world/covid-vaccinations-tracker.html).

10. Keech C, Albert G, Cho I, et al. Phase 1–2 trial of a SARS-CoV-2 recombinant spike protein nanoparticle vaccine. N Engl J Med 2020;383:2320–32.

11. Formica N, Mallory R, Albert G, et al. Evaluation of a SARS-CoV-2 vaccine NVXCoV2373 in younger and older adults. medRxiv 2021.02.26.21252482. doi: https://doi.org/10.1101/2021.02.26.21252482.

12. Shinde V, Bhikha S, Hoosain Z, et al. Safety and efficacy of NVX-CoV2373 Covid-19 vaccine against the B.1.351 variant. N Engl J Med 2021;384:1899–1909.

13. Heath PT, Galiza EP, Baxter DN, et al. Safety and efficacy of the NVX-CoV2373 Covid-19 vaccine. 2021;385(13):1172–83.

14. US Food and Drug Administration. Guidance for Industry: Development and Licensure of Vaccines to Prevent COVID-19; June 2020. (https://www.fda.gov/media/139638/download).

15. Corey L, Mascola JR, Fauci AS, Collins FS. A strategic approach to COVID-19 vaccine R&D. Science 2020;368:948–50.

16. Joffe S, Babiker A, Ellenberg SS, et al. Data and safety monitoring of COVID-19 vaccine clinical trials. J Infect Dis 2021; doi: 10.1093/infdis/jiab263.

17. Centers for Disease Control and Prevention. People with certain medical conditions. Updated May 13, 2021. (https://www.cdc.gov/coronavirus/2019-ncov/need-extra-precautions/people-with-medical-conditions.html).

18. US Food and Drug Administration, Center for Biologics Evaluation and Research (US). Guidance for Industry: Toxicity Grading Scale for Healthy Adult and Adolescent Volunteers Enrolled in Preventive Vaccine Clinical Trials. September 2007. (https://www.fda.gov/media/73679/download).

19. US Food and Drug Administration. Coronavirus (COVID-19). Update: July 13, 2021. (https://www.fda.gov/news-events/press-announcements/coronavirus-covid-19-update-july-13-2021).

20. US Centers for Disease Control and Prevention. Myocarditis and Pericarditis After mRNA COVID-19 Vaccination. (https://www.cdc.gov/coronavirus/2019-ncov/vaccines/safety/myocarditis.html).

21. Hunter PR. Thrombosis after covid-19 vaccination. BMJ 2021;373:n958.

22. Follmann D, Fintzi J, Fay MP, et al. A deferred-vaccination design to assess durability of COVID-19 vaccine effect after the placebo group is vaccinated. Ann Intern Med 2021;174:1118–25.

