## Supplemental Appendix for "Efficacy and Safety of NVX-CoV2373 in Adults in the United States and Mexico"

### Supplementary Appendix

This appendix has been provided by the authors to give readers additional information about their work.
Supplement to: Dunkle LM, Kotloff KL, Gay CL, et al. Efficacy and Safety of NVX-CoV2373 in Adults in the United States and Mexico.

#### 2019nCoV-301 Study Group (Pubmed listed, in alphabetical order of institution affiliation)

| Affiliation/Funding* | Study Group | Location |
|---|---|---|
| **México** | | |
| Centro de Atención e Investigación Médica (CAIMED) | Jorge F. Méndez Galván, MD, Monica B. Carrascal, Adriana Sordo Duran, Laura Ruy Sanchez Guerrero, Martha Cecilia Gómora Madrid | Mexico City, Mexico |
| FAICIC Clinical Research | Alejandro Quintín Barrat Hernández, MD, Sharzhaad Molina Guizar, Denisse Alejandra González Estrada, Silvano Omar Martínez Pérez, MD, Zindy Yazmín Zárate Hinojosa, MD | Veracruz, Mexico |
| Instituto Nacional de Ciencias Médicas y Nutrición Salvador Zubirán | Guillermo Miguel Ruiz-Palacios, MD | Mexico City, Mexico |
| Instituto Nacional de Salud Pública | Aurelio Cruz-Valdez, PhD, Janeth, Pacheco-Flores, MD, Anyela Lara, MD, Secia Díaz-Miralrio | Cuernavaca, Mexico |
| PanAmerican Clinical Research México | María José Reyes Fentanes, MD, Jocelyn Zuleica Olmos Vega, MD, Daniela Pineda Méndez, MD, Karina Cano Martínez, MD, Winniberg Stephany Alvarez León | Querétaro, Mexico |
| PanAmerican Clinical Research México | Vida Veronica Ruiz Herrera, MD, Eduardo Gabriel Vázquez Saldaña, Laura Julia Camacho Choza, Karen Sofia Vega Orozco, Sandra Janeth Ortega Domínguez | Guadalajara, Mexico |
| Unidad de Atención Médica e Investigación en Salud (UNAMIS) | Jorge A. Chacón, MD, Juan J. Rivera, MD, Erika A. Cutz, MD, Maricruz E. Ortegón, MD, María I. Rivera, MD | Mérida, Mexico |
| **United States and Puerto Rico** | | |
| Accellacare | David Browder, MD, Cortney Burch, Terri Moye, Paul Bondy, MD, Lesley Browder, MD | Rocky Mount, NC |
| Accellacare | Rickey D. Manning, MD, James Wilson Hurst, MD, Rodney E. Sturgeon, MD, Paul H. Wakefield, MD, John A. Kirby, MD | Knoxville, TN |
| Accel Research Sites | James Andersen, MD, Szheckera Fearon, MSN, FNP-C, Rosa Negron, MD, Amy Medina, ADN, BS | Lakeland, FL |
| Accel Research Sites | Bruce Rankin, DO, John M. Hill, MD, Steven Shinn, MD, Vivek Rajasekhar, DO, Marshall Nash, MD | DeLand, FL |
| Achieve Clinical Research | Hayes Williams, MD, PhD, LaShondra Cade, Rhodna Fouts, Connie Moya | Birmingham, AL |
| Alliance for Multispecialty Research | Corey G. Anderson, MD, Naomi Devine, NP-C, James Ramsey, NP-C, Ashley Perez, David Tatelbaum | Tempe, AZ |
| Alliance for Multispecialty Research | Michael Jacobs, MD, Kathleen Menasche, LPN, Vincent Mirkil, MD | Las Vegas, NV |
| Anaheim Clinical Trials | Peter J. Winkle, MD, Amina Z. Haggag, MD, Michelle Haynes, Marysol Villegas, Sabina Raja | Anaheim, CA |
| Atlanta Center for Medical Research | Robert Riesenberg, MD, Stanford Plavin, MD, Mark Lerman, MD, Leana Woodside, DNP, NP-C, Maria Johnson, MD | Atlanta, GA |
| Baylor College of Medicine / NIAID (UM1AI148575) | C. Mary Healy, MD, Jennifer A. Whitaker, MD, Hana El Sahly, MD, Christine Akamine, MD, Wendy A. Keitel, MD, Robert L. Atmar, MD | Houston, TX |
| Biomedical Advanced Research and Development Authority (BARDA) | Richard Gorman, MD, Gary Horwith, MD, Robin Mason, MS, MBA | Washington, DC |
| Benchmark Research | Laurence Chu, MD, Michelle Chouteau, MD, Lisa Johnson, FNP, Tambra Dora | Austin, TX |
| Benchmark Research | Greg Hachigian, MD, Deborah Murray, FNP, Michael Cancilla, PA, Logan Ledbetter, PA, Masaru Oshita, MD | Sacramento, CA |
| Benchmark Research | William Seger, MD, Beverly Ewing, APRN, DNP, FNP-BC | Fort Worth, TX |
| Beth Israel Deaconess Medical Center / NIAID (UM1AI068614) | Kathryn E. Stephenson, MD, MPH, Chen Sabrina Tan, MD, Rebecca Zash, MD, Jessica L. Ansel, MSN, Kate Jaegle, MSN, Caitlin J. Guiney, MSN | Boston, MA |
| Black Hills Center for American Indian Health / Missouri Breaks Industries Research Inc / NIAID (UM1AI068614) | Jeffrey A. Henderson, MD, MPH, Marcia O'Leary, RN, Kendra Enright, RN, Jill Kessler, MS, Pete Ducheneaux, LPN, Asha Inniss, MS, APRN | Eagle Butte, SD |
| California Research Foundation | Donald M. Brandon, MD, William B. Davis, MD, Daniel T. Lawler, MD | San Diego, CA |
| Carolina Institute for Clinical Research | Yaa D. Oppong, MD, Ryan P. Starr, DO, Scott N. Syndergaard, DO, Rozeli Shelly, MD, Mashrur Islam Majumder | Fayetteville, NC |
| Cedar Crosse Research Center | Danny Sugimoto, MD, Jeffrey Dugas Sr., MD, Dolores Rijos, Sandra Shelton, Stephan Hong, MD | Chicago, IL |
| Cenexel RCA | Howard Schwartz, MD, Nelia Sanchez-Crespo, MD, Jennifer Schwartz, APRN, Terry Piedra, BS, Barbara Corral, APRN | Hollywood, FL |
| Centex Studies | Joel Solis, MD, Carmen Medina, PA, Westley Keating, PA | McAllen, TX |
| Clinical Neuroscience Solutions | Michael E. Dever, MD, Mitul Shah, MD, Michael Delgado, MD, Tameika Scott, DrPH | Orlando, FL |
| Clinical Neuroscience Solutions | Lisa S. Usdan, MD, Lora J. McGill, MD, Valerie K. Arnold, MD, Carolyn Scatamacchia, MSN, NP-C, Codi M. Anthony, DNP, APRN, PMHNP-BC | Memphis, TN |
| CommonSpirit Health Research Institute | Rajan Merchant, MD, Anelgine Crans Yoon, MD, Janet Hill, PA-C, Lucy Ng-Price, MA, Teri Thompson-Seim | Woodland, CA |
| Comprehensive Clinical Research | Ronald Ackerman, MD, Jamie Ackerman, Florida Aristy, APRN | West Palm Beach, FL |
| Covid-19 Prevention Network (CoVPN) | Lawrence Corey, MD, Kathleen M Neuzil, MD, MPH, Huub G Gelderblom, MD, PhD, Nzeera Ketter, Carrie Sopher | Seattle, WA |
| CRA Headlands | Jon Finley, MD, Nathan Segall, MD, Mildred Stull, APRN, FNP-C | Stockbridge, GA |
| DM Clinical Research | Vicki E. Miller, MD, MPH, Monica Murray, Blanca Gomez, Zainab Rizvi, Sonia Guerrero | Tomball, TX |
| Empire Clinical Research | Yogesh K. Paliwal, MD, Amit Paliwal, MD, Sarah Gordon, MS, Bryan Gordon, Cynthia Montano-Pereira | Pomona, CA |
| Headlands Research | Christopher Galloway, MD, Candice Montros, Lily Aleman, Samira Shairi, RN, Wesley Van Ever | Orlando, FL |
| Health Research of Hampton Roads | George H. Freeman, MD, Esther Laverne Harmon, ANP, Marshall A. Cross, MD, Kacie Sales, BSN, RN, Catherine Q. Gular, PharmD | Newport News, VA |
| HHS-DoD Countermeasures Acceleration Group | Matthew Hepburn, MD | Washington, DC |

| | | |
|---|---|---|
| HOPE Research Institute | Matthew Doust, MD, Nathan Alderson, PhD, Shana Harshell | Phoenix, AZ |
| Howard University Hospital / Howard University College of Medicine / NIAID (UM1AI068614) | Siham Mahgoub, MD, Celia Maxwell, MD, Thomas Mellman, MD, Karl M Thompson, PhD, Glenn Wortman, MD | Washington, DC |
| IACT Health | Jeff Kingsley, DO, April Pixler, LaKondria Curry, Sarah Afework, Austin Swanson | Columbus, GA |
| Jacksonville Center for Clinical Research | Jeffry Jacqmein, MD, Maggie Bowers, PA-C, Dawn Robison, APRN-C, Victoria Mosteller, MD, Janet Garvey, DNP | Jacksonville, FL |
| Johnson County Clin-Trials | Carlos Fierro, MD, Mary Easley, BSN, RN | Lenexa, KS |
| Joint Program Executive Office for Chemical, Biological, Radiological and Nuclear Defense's, US Department of Defense | Rebecca J. Kurnat | Washington, DC |
| Lynn Health Science Institute | Carl P. Griffin, MD, Raymond Cornelison, MD, Shanda Gower, APRN, CNP, William Schnitz, MD, Destiny S. Heinzig-Cartwright, BA | Oklahoma City, OK |
| Lynn Institute of the Ozarks | Derek Lewis, MD, Fred E. Newton, MD, Aeiress Duhart, Breana Watkins, Brandy Ball | Little Rock, AR |
| Lynn Institute of the Rockies | Ripley Hollister, MD, Jeremy Brown, DO, Melody Ronk, PA-C, Jill York, Shelby Pickle | Colorado Springs, CO |
| M3-Emerging Medical Research | David B. Musante, MD, William P. Silver, MD, Linda R. Belhorn, MD, Nicholas A. Viens, MD, David Dellaero, MD | Durham, NC |
| M3-Wake Research | Matthew Hong, MD, Wayne Harper, MD, Lisa Cohen, DO, Priti Patel, NP, Kendra Lisec, PA | Raleigh, NC |
| MD Clinical | Beth Safirstein, MD, Luz Zapata, MD, Lazaro Gonzalez, APRN, Evelyn Quevedo, APRN, Farah Irani, PhD | Hallandale Beach, FL |
| Medical Research International | Joseph Grillo, MD, Amy Potts, PA-C, MPH, Julie White, MBA | Oklahoma City, OK |
| Medical University of South Carolina | Patrick Flume, MD, Gary Headden, MD, Brandie Taylor, NP, Ashley Warden, Amy Chamberlain | Charleston, SC |
| MedPharmics | Robert Jeanfreau MD, Susan Jeanfreau MD | Metairie, LA |
| MedPharmics | Paul G. Matherne, MD, Amy Caldwell, RN, Jessica Stahl, Mandy Vowell, Lauren Newhouse | Gulfport, MS |
| Meharry Medical College / NIAID (UM1AI068614) | Vladimir Berthaud MD, MPH, Zudi-Mwak Takizala MD, MPH, MBA, Genevieve Beninati, FNP, Kimberly Snell, PharmD, Sherrie Baker, BS, James Walker, RN | Nashville, TN |
| Meridian Clinical Research | David Ensz, MD, Tavane Harrison, CNP, Meagan Miller, Janet Otto | Sioux City, IA |
| Meridian Clinical Research | Brandon Essink, MD, Roni Gray, APRN, Christine Wilson, Tiffany Nemecek, Hannah Harrington, MPH | Omaha, NE |
| Meridian Clinical Research | Charles Harper, MD, Keith Vrbicky, MD, Chelsie Nutsch, NP, Sally Eppenbach, NP, Wendell Lewis, NP | Norfolk, NE |
| Meridian Clinical Research | Jordan Whatley, MD, Christopher Dedon, APRN, FNP-C, Tana Bourgeois, RN, Lyndsea Folsom, Crystal Rowell, APRN, FNP-C | Baton Rouge, LA |
| Miami Veterans Affairs Medical Center / NIAID (UM1AI068614) | Gregory Holt, MD, Mehdi Mirsaeidi, MD, Rafael Calderon, MD, Paola Lichtenberger, MD, Jalima Quintero, RN, Becky Martinez, RN | Miami, FL |
| Morehouse School of Medicine / NIAID (UM1AI068614) | Lilly Immergluck, MD, Erica Johnson, PhD, Austin Chan, MD, Norberto Fas, MD, LaTeshia Thomas-Seaton, MS, APRN, Saadia Khizer, MD, MPH | Atlanta, GA |
| MultiCare Institute for Research and Innovation | Jonathan Staben, MD | Cheney, WA |
| National Institute of Allergy and Infectious Diseases (NIAID) / National Institutes of Health (NIH) | Tatiana Beresnev, MD, Maryam Jahromi, MD, Mary A. Marovich, MD, Julia Hutter, MD, Martha Nason, PhD, Julie Ledgerwood, DO, John Mascola, MD | Bethesda, MD |
| National Research Institute | Mark Leibowitz, MD, Fernanda Morales, Mike Delgado, Rosario Sanchez, Norma Vega | Los Angeles, CA |
| Novavax, Inc. | Lisa M. Dunkle, MD, Germán Áñez, MD, Gary Albert, Erin Coston, Chinar Desai, Haoua Dunbar, Mark Eickhoff, Jenina Garcia, Margaret Kautz, Angela Lee, Maggie Lewis, Irene McKnight, Joy Nelson, Patrick Newingham, Patty Price-Abbott, Patty Reed, Diana Vegas, Bethanie Wilkinson, PhD, Katherine Smith, MD, Wayne Woo, MS, Iksung Cho, MS, Gregory M. Glenn, MD, Filip Dubovsky, MD, MPH | Gaithersburg, MD |
| Omega Medical Research | David L. Fried, MD, Lynne A. Haughey, MSN, FNP, Ariana C. Stanton, PA-C, Lisa Stevens Rameaka, MD | Warwick, RI |
| Pharmacology Research Institute | David Rosenberg, MD, Lee Tomatsu, Viviana Gonzalez, Millie Manalo | Los Alamitos, CA |
| PMG Research of Bristol | Bernard Grunstra, MD, Donald Quinn, MD, Phillip Claybrook, MD, Shelby Olds, MD, Amy Dye | Bristol, TN |
| PMG Research of Wilmington | Kevin D. Cannon, MD, Mesha M. Chadwick, MD, Bailey Jordan, Morgan Hussey, Hannah Nevarez | Wilmington, NC |
| Ponce de Leon Center / NIAID (UM1AI068614) | Colleen F. Kelley, MD, MPH, Valeria D. Cantos MD, Michael Chung MD, Caitlin Moran, MD, MSc, Paulina Rebolledo, MD, Christina Bacher, PAC | Atlanta, GA |
| Ponce School of Medicine / NIAID (UM1AI148685) | Elizabeth Barranco-Santana, MD, Jessica Rodriguez, MD, Rafael Mendoza, MD, Karen Ruperto, MD, Odette Olivieri, MD, Enrique Ocaña, MD | Ponce, Puerto Rico |
| Preferred Research Partners | Paul E. Wylie, MD, Renea Henderson, DO, Natasa Jenson, MD, Fan Yang, MD, Amy Kelley, BSN, RN | Little Rock, AR |
| Providea Health Partners Elligo Health Research | Kenneth Finkelstein, DO, David Beckmann, MD, Tanya Hutchins, FNP, Sebastian Garcia Escallon, BA, Kristen Johnson | Evergreen Park, IL |
| Providence Clinical Research | Teresa S. Sligh, MD, Parul Desai, NP, Vincent Huynh, BSc, Carlos Lopez, MD, Erika Mendoza, BA | North Hollywood, CA |
| Research Your Health | Jeffrey Adelglass, MD, Jerome (Jerry) G. Naifeh, MD, Kristine Jane Kucera, PA-C, MPAS, DHS, Waseem Chughtai, BS, MBBS, Shireen Hasham Jaffer | Plano, TX |
| Rochester Clinical Research | Matthew G. Davis, MD, Jennifer Foley, Michelle Lyn Burgett, RN, Tammi Louise Shlotzhauer, MD, Sarah Michelle Ingalsbe-Geno, RPA-C | Rochester, NY |
| SIMEDHealth / SIMEDResearch | Daniel Duncanson, MD, Kelly Kush, Lori Nesbitt, Cora Sonnier, Jennifer McCarter | Gainesville, FL |

| | | |
|---|---|---|
| Sterling Research Group | Michael B. Butcher, MD, James Fry, PA-C, Donna Percy, RN, BSN, Karen Freudemann | Cincinnati, OH |
| Sterling Research Group | Bruce C. Gebhardt, MD, Padma N. Mangu, MD, Debra Beck Schroeck, MS, PA-C, Rajesh Kumar Davit, MD, Gayle D. Hennekes, PA-C, MPAS | Cincinnati, OH |
| Stony Brook University - Stony Brook Medicine / NIAID (UM1AI068614) | Benjamin J. Luft, MD, Melissa Carr, BA, Sharon Nachman, MD, Alison Pellecchia, BA, Candace Smith, PharmD, Bruno Valenti, NP | Commack, NY |
| Suncoast Research Associates | Maria I. Bermudez, MD, Noris Peraita, ARNP, Ernesto Delgado, ARNP, Alicia Arrazcaeta, Natalie Ramirez | Miami, FL |
| Suncoast Research Group | Mark E. Kutner, MD, Jorge Caso, MD, Janet Mendez, ARNP, Marianela Carvajal, ARNP, Carmen Amador, ARNP | Miami, FL |
| Sundance Clinical Research | Larkin Tyler Wadsworth III, MD, Horacio Marafioti, MD, Lyly Dang, DNP-BC, Lauren Clement, NP-C, Jennifer Berry, FNP-BC | St. Louis, MO |
| Synexus Clinical Research | Mohammed Allaw, MD, Georgettea Geuss, Chelsea Miles, NP, Zachary Bittner, Melody Werne | Evansville, IN |
| Synexus Clinical Research | Cornell Calinescu, MD, Shannon Rodman, Joshua Rindt | Henderson, NV |
| Synexus Clinical Research | Erin Cooksey, MD, Kristina Harrison, Deanna Cooper, Manisha Horton Amanda Philyaw | Anderson, SC |
| Synexus Clinical Research | William Jennings, MD, Hilario Alvarado, MD, Michele Baka, MD, Malina Regalado, NP | San Antonio, TX |
| Synexus Clinical Research | Linda Murray, DO | Pinellas Park, FL |
| Synexus Clinical Research | Sherif Naguib, MD, Justin Singletary, Sha-Wanda Richmond, Sarah Omodele, Emily Oppenheim | Atlanta, GA |
| Synexus Clinical Research | Joseph Newberg, MD, Laura Pearlman, MD, Reuben Martinez, Victoria Andriulis | Chicago, IL |
| Synexus Clinical Research | Paul J. Nugent, DO, Leonard Singer, MD, Jeanne Blevins, Meagan Thomas, Christine Hull | Cincinnati, OH |
| Synexus Clinical Research | Isabel Pereira, MD, Gina Rivero, Tracy Okonya, Frances Downing, Paulina Miller | Vista, CA |
| Synexus Clinical Research | Margaret Rhee, MD, Katherine Stapleton, Jeffrey Klein, Rosamond Hong, MD | Akron, OH |
| Synexus Clinical Research | Suzanne Swan, MD, Tami Wahlin, MD, Elizabeth Bennett, PA, Amy Salzl Sharine Phan | Richfield, MN |
| Synexus Clinical Research | Jewel Johnny White, MD, Amanda Occhino, Ruth Paiano APRN, Morgan McLaughlin APRN, Elisa Swieboda APRN | The Villages, FL |
| Texas Center for Drug Development | Veronica Garcia-Fragoso, MD, Maria Gabriela Becerra, MD, Cecilia Mckeown, Lisa Holloway, Toni White | Houston, TX |
| The Charlotte-Mecklenburg Hospital Authority d/b/a Atrium Health / NIAID (UM1AI068614) | Christine B. Turley, MD, Andrew McWilliams, MD, Tiffany Esinhart, PA-C, Natasha Montoya, APRN, Shamika Huskey, FNP, Leena Paul, FNP | Charlotte, NC |
| The Miriam Hospital / NIAID (UM1AI068636) | Karen Tashima, MD, Jennie Johnson, MD, Marguerite Neill, MD, Martha Sanchez, MD, Natasha Rybak, MD, Maria Mileno, MD | Providence, RI |
| UC Davis Health / NIAID (UM1AI068614) | Stuart H. Cohen, MD, Monica Ruiz, Dean M. Boswell, BS, Elizabeth E. Robison, BS, Trina L. Reynolds, BS, Sonja Neumeister, MPH | Sacramento, CA |
| Universidad de Puerto Rico - Recinto de Ciencias Médicas - Maternal Infant Studies Center (CEMI) / NIAID (UM1AI068636) | Carmen D. Zorrilla, MD, Juana Rivera, MD, MPH, Jessica Ibarra, MD, Iris García, BSN, RN, Dianca Sierra, BA, Wanda Ramon, BSPh | San Juan, Puerto Rico |
| University of Colorado Hospital CRS / NIAID (UM1AI068636) / NCATS (UL1TR002535, UM1AI069432) | Thomas B. Campbell, MD, Suzanne Fiorillo, MSPH, Rebecca Pitotti, RNP, Victoria Riedel Anderson, MS, Jose Castillo Mancilla, MD, Nga Le, PharmD | Aurora, CO |
| University of Iowa Medical Center / NIAID (UM1AI068614) / NCATS (UL1TR002537) | Patricia L. Winokur, MD, Dilek Ince, MD, Theresa Hegmann, PA, Jeffrey Meier, MD, Jack Stapleton, MD, Laura Stulken, PA | Iowa City, IA |
| University of Maryland School of Medicine / NIAID (UM1AI148689) | Monica McArthur, MD, PhD, Karen L. Kotloff, MD, Kathleen Neuzil, MD, Andrea Berry, MD, Milagritos Tapia, MD, Elizabeth Hammershaimb, MD, MS, Toni Robinson, RN, Rosa MacBryde, RN | Baltimore, MD |
| University of Minnesota / NIAID (UM1AI068614) | Susan Kline, MD, MPH, Joanne L. Billings, MD, MPH, Winston Cavert, MD, Les B. Forgosh. MD, Timothy W. Schacker, MD, Tyler D. Bold, MD, PhD | Minneapolis, MN |
| University of Missouri Health Care / NIAID (UM1AI148685) | Dima Dandachi, MD, MPH, Taylor Nelson, DO, Andres Bran, MD, Grant Geiger, S. Hasan Naqvi, MD | Columbia, MO |
| University of Nebraska Medical Center / NIAID (UM1AI068614) | Diana F Florescu, MD, Richard Starlin, MD, David Kline, MD, Andrea Zimmer, MD, Anum Abbas, MD, Natasha Wilson, APRN | Omaha, NE |
| University of North Carolina / NIAID (UM1AI068619) / University of North Carolina at Chapel Hill Center for AIDS Research (P30AI050410) / NC TraCS Institute (UL1TR002489) | Cynthia L. Gay, MD, MPH, Joseph J Eron, MD, Michael Sciaudone, MD, MPH, A. Lina Rosengren, MD, MPH, MS, John S Kizer, MD, Sarah E Rutstein, MD, PhD | Chapel Hill, NC |
| University of South Florida, Morsani College of Medine / NIAID (UM1AI068614) | Carina A. Rodriguez, MD, Elizabeth Bruce, MD, Claudia Espinosa, MD, Lisa J Sanders, MD, Kami Kim, MD, Denise Casey, RN | Tampa, FL |
| University of Texas Health Science Center San Antonio / NIAID (UM1AI068614) | Barbara S. Taylor, MD, MS, Thomas Patterson, MD, Ruth Serrano Pinilla, MD, Delia Bullock, MD, Philip Ponce, MD, Jan Patterson, MD | San Antonio, TX |
| University of Washington / Lummi Tribal Health Center / NIAID (UM1AI148573) | R. Scott McClelland, MD, MPH, Dakotah C. Lane, MD, Anna Wald, MD, MPH, Frank James, MD, Elizabeth Duke, MD, Kirsten Hauge, MPH, Jessica Heimonen, MPH | Seattle, WA |
| University of Washington | Robert W. Coombs, MD, PhD, Alex Greninger, MD, PhD, MS, MPhil, Pavitra Roychoudhury, PhD, Erin A. Goecker, MS, Yunda Huang, PhD, Youyi Fong, PhD | Seattle, WA |
| VA Ann Arbor Healthcare System / NIAID (UM1AI068614) | Carol Kauffman, MD, Kathleen Linder, MD, Kimberly Nofz, BSN, Andrew McConnell, BS | Ann Arbor, MI |

| | | |
|---|---|---|
| Velocity Clinical Research | Robert J. Buynak, MD, Angella Webb, APRN, Taryn Petty, FNP, Stephanie Andree, FNP | Valparaiso, IN |
| Velocity Clinical Research | Judith Kirstein, MD, Marcia Bernard, Erica Sanchez, Nolan Mackey, Clarisse Baudelaire | Banning, CA |
| Velocity Clinical Research | Gregg Lucksinger, MD, Jaleh Ostovar, NP | Medford, OR |
| Velocity Clinical Research | Mary Beth Manning, MD, Joan Rothenberg, MD, Toby Briskin, MD, Denise Roadman, PAC, Sarah Dzigiel | Cleveland, OH |
| Velocity Clinical Research | J. Scott Overcash, MD, Adrienna Marquez, Hanh Chu, Kia Lee, Kim Quillin | La Mesa, CA |
| Velocity Clinical Research | Barbara Rizzardi, MD, Michelle King, NP, Vanessa Abad, NP, Jennifer Knowles, BS | West Jordan, UT |
| Velocity Clinical Research | Michael Waters, MD, Karla Zepeda, NP, Scott Overcash, MD, Jordan Coslet, NP, Dalia Tovar, MA | Chula Vista, CA |
| Velocity Clinical Research | Marian E. Shaw, MD, Mark A. Turner, MD, Cory J. Huffine, FNP-C, Esther S. Huffine, FNP-C | Meridian, ID |
| Walter Reed Army Institute of Research | Julie A. Ake, MD, MSc | Silver Spring, MD |
| Wayne State University / NIAID (UM1AI068614) | Elizabeth Secord, MD, Eric McGrath, MD, Phillip Levy, MD, Brittany Stewart, RD, PharmD, Charnell Cromer, RN, MSN, Ayanna Walters, RN, BSN | Detroit, MI |
| Weill Cornell Chelsea CRS / NIAID (UM1AI068619) | Kristen Marks, MS, MD, Grant Ellsworth, MD, MS, Caroline Greene, ANP-BC, Sarah Galloway, BA, Shashi Kapadia, MD, MS, Elliot DeHaan, MD | New York, NY |
| Willis-Knighton Health System / WKB Family Medicine Associates | Clint Wilson, MD, Jason Milligan, MD, Danielle Raley, MD, Joseph Bocchini, MD | Bossier City, LA |
| Womack Army Medical Center | Bruce McClenathan, MD, Mary Hussain, BS, Evelyn Lomasney, MD, Evelyn Hall, MMS, PA-C, Sherry Lamberth, PharmD | Fort Bragg, NC |
| WR Clinsearch | Mark McKenzie, MD, Teresa Deese, Christy Schmeck, Vickie Leathers, Christy Sweet | Chattanooga, TN |

* Funding of institutions by the National Institute of Allergy and Infectious Diseases (NIAID) and/or research support by the National Center for Advancing Translational Science (NCATS), as indicated. All other institutions were funded by Office of the Assistant Secretary for Preparedness and Response, Biomedical Advanced Research and Development Authority. The content of this publication is solely the responsibility of the authors and does not necessarily represent the official views of the funding sources.

#### 2019nCoV-301 Principal Investigators and Study Team (in alphabetical order)

| Principal Investigator | Study Team | Institution | Location |
|---|---|---|---|
| Ronald Ackerman, MD | Jamie Ackerman, Florida Aristy, Tomeko Heard, Diana Mann, Maureen Stewart, Cheryl Demczyk, Rohan Barron, Ashley Torres, Jennifer Gomez, Tiffany Potter | Comprehensive Clinical Research | West Palm Beach, FL |
| Jeffrey Adelglass, MD | Jerome (Jerry) G. Naifeh, Kristine Jane Kucera, Waseem Chughtai, Shireen Hasham Jaffer, Anuja Sathe, Cameron Galownia, Cheryl Hill, Ramiro Lopez, Erica Parker-Martinez, Helene Harrison, Chiedza Mutindori, Sabrina Flowers, Tamara Betters, Carolyn Ackley, Pamela Fox, Noelia Tejada James, Dorothy Saylor, Hallen Dao, Jon Etta Randolph, Jason Tentativa, Malaika Chughtai, Shanzae Chughtai, Maheen Shah, Hayyan Chughtai, Tyler Love, Ti'arah Love | Research Your Health | Plano, TX |
| Mohammed Allaw, MD | Georgettea Geuss, Chelsea Miles, Zachary Bittner, Melody Werne, Lyndsey Morrison, Stephanie Albin, Linda Frazier, Jacque Nalley, Christie Borin, Jacque Nalley | Synexus Clinical Research | Evansville, IN |
| James Andersen, MD | Szheckera Fearon, Rosa Negron, Amy Medina, Diana Holmes, Colleen Figueroa, Cristal Ruiz, Nancy Masseus Tare Floyd, Kenta Oliver, Candice Gerber, Mae Ann Francisco, Gilbert de la Cruz, Ginny McClanahan, Veronica Walker, David Irwin, Gloria Adejobi | Accel Research Sites | Lakeland, FL |
| Corey G. Anderson, MD | Naomi Devine, James Ramsey, Tyanna Montijo, Ashley Perez, David Tatelbaum, Lisa M. Dean, Angela D. Ledezma, Anthony Padilla, Cecilia M. Tanori, Georgina Lopez-Wood, Tasha C. Marriott, Ronald Hawkins, Hannah Spinks | Alliance for Multispecialty Research | Tempe, AZ |
| Elizabeth Barranco-Santana, MD | Michele Irizarry, Alice Grace Rodriguez, Irmaris Arroyo, Sara Cancel, Alejandra Román, Juan D. Lugo, Armando X. Torres, Marianne Hernandez, Brenda Garcia, Nancy Jiménez, Orlando Torres | Ponce School of Medicine | Ponce, Puerto Rico |
| Alejandro Quintín Barrat Hernández, MD | Sharzhaad Molina Guizar, Denisse Alejandra González Estrada, Silvano Omar Martínez Pérez, Zindy Yazmín Zárate Hinojosa, Norberto Daniel Vázquez Tinajero, Yessica Olivo Domínguez, Daniel Hernández León, Gloria Norma Ambrosio Lara, José Carlos Mateos Castro, Irving Nerí Leyva Ferrer, María Fernanda Hernández García, Heidy Jazmín Maldonado Pavón, Evelyn Monserrat Bravo Serralta, Edgar Iván Muñoz López, Karina Esmeralda García Mateo, Lorena Cruz Cruz, José Javier Zárate Hinojosa, Javier Torres Cole, Yareth Jiménez Barcenas, Andrea Anaid Rangel Huerta, Erika Guillén González, María de la Luz Rufina Martínez Lugo, Angélica Liliana Muñoz Solano, David Sena Gómez, Berenice Valera Montalvo, Moisés Miguel Ruíz Nogueira, Yoshira Montero Díaz, Francisco Javier Martínez Osorio, Alejandra Morales Arias, Sandra Itzel Solis Rivera, Alejandro Esteban Cortina, Aldo Miguel López Domínguez, María Fernanda Cortés Ruíz, Marilyn Yulissa Ramírez Domínguez, Lucero Moctezuma Juan, Francisco Barrales Arcos | FAICIC Clinical Research | Veracruz, Mexico |
| Maria I. Bermudez, MD | Noris Peraita, Ernesto Delgado, Alicia Arrazcaeta, Natalie Ramirez, Giovanna Salcedo, Aliana Amador, Elizabeth Martinez, Arleen Aspuru, Gabriella Gonzalez, Gabriella Alabaci, Livan Sanchez, Raul Tejeda, Adriana Bello, Barbara Vega-Aguera, Kassandra Martinez, Grettel Obregon, Oscar Alejandro Gutierrez Luna, Magela C. Dominguez, Lauren Pena | Suncoast Research Associates | Miami, FL |
| Vladimir Berthaud, MD, MPH | Toni Hall, Livette Johnson, Sylvia Eluhu, Ana Tomescu, Katharina Whitbeck, Rajbir Singh | Meharry Medical College | Nashville, TN |
| Donald M. Brandon, MD | William B. Davis, Daniel T. Lawler, Maria Aceves, Kathleen B. Anderson, Hana Berry, Janice E. Brandon, Jeffrey C. Brandon, Patricia A. Brandon, Lorraine Boggs, Charlene Cruz, Mairead Hawkins, Clarice Hranicky, Andrew J. McCrea, Karen G. McCrea, Kimberly Najera, Tierney J. O'Connor, Michelle L. Rios, Cindy F. Stevens, Hannah J. Zapata | California Research Foundation | San Diego, CA |
| David Browder, MD | Cortney Burch, Terri Moye, Michael Wright, Paul Bondy, Lesley Browder | Accellacare | Rocky Mount, NC |
| Michael B. Butcher, MD | James Fry, Julia Froschauer, Allison Deuel, Jeanne Piccola, Donna Percy, Karen Freudemann, Lois Rawe, Megan Bryant, Kurt Percy, Jon Marvin, Luann Corcoran | Sterling Research Group | Cincinnati, OH |
| Robert J. Buynak, MD | Mark Yarosz, Rachel McNeal, Megan Smith, Patricia Volom, Nicholas Hanna, Erica Lewis, Miranda Lee, Goldie Luna, Marilyn Idowu, Destiny Williams, Jessica Johnson, Consuelita Perez, Priscilla Dodson | Velocity Clinical Research | Valparaiso, IN |
| Cornell Calinescu, MD | Shannon Rodman, Joshua Rindt, Krystal Tyner, Lovelyn Vincente, Alejandro Osuna-Meda, Charmaine Brown, Matthew Derrick, Melodee Morrison, Marissa Washington | Synexus | Henderson, NV |
| Thomas B. Campbell, MD | Donna McGregor, Laurel Ware, Myron Levin, Steven Johnson, Sophia Quesada, Martin Krsak, Kristine Erlandson, Nicholas Sarchet, Vanessa Sutton, Lawrence Moran, Tracey Stevenson, Alaina Dougherty, Julianne Randlemon | University of Colorado Hospital CRS | Aurora, CO |
| Kevin D. Cannon, MD | Mesha M. Chadwick, Bailey Jordan, Taylor Fedorcha, Kathryn Zweier, Brettany Holt, Emily Johnson, Karen Ruggiero, Olivia Houghton, Courtney Christie, Allison Dunn, Courtney Boyce, Sasha Saint-Lot, Ashley Andrades, Ashley Miller, LaShaya Dunston, Russell Larkins, Brittany Savoca, Hannah Nevarez, Hannah Nevarez, Larkin Collins, Morgan Cyrus, Morgan Hussey, Christina MacNaughton, Heidi Kaufman, Sheila Gard, Alyssa Gaylor, Bethany Donelan-Wilson, Taylor Bayless, Anna McManus, Tracie Marlowe Bryant, Ben Manuel, Laura McMillan, Nicole Stigers, Prerana Zanke | PMG Research of Wilmington | Wilmington, NC |
| Jorge A. Chacon, MD | Juan J. Rivera, Erika A. Cutz, Maricruz E. Ortegon, María I. Rivera, Ricardo Cervera, Felipe Rivera, Daniela Pat, Daniela Cruz, Alberto Chacon, Kattia Borges, Aldo Borraz, Rebeca Ortegon, Karla Ic, Carmen Ojeda, Irvin Ortega, Mayra Jimenez, Cindy Novelo, Pharmacist, Mónica Pérez, Adriana Hernandez, Laura Martinez | Unidad de Atención Medica e Investigación en Salud (UNAMIS) | Merida, Mexico |
| Laurence Chu, MD | Michelle Chouteau, Lisa Johnson, Tambra Dora, Lamar Box, Michelle Listz, Katherine Davis, Jennifer Montes, Jessica Ruff, Jennifer Leyva, Pamela Fidler, Ruth Fitch, Sean Turnbow, Francesca Vigil, Maria Barrientes, Isaiah Knight, Cindy Duran, Lauren Christal, Breana Wade Liaison, Brooke Harris, Dean Skiles, Marisol Ramos, Brandon Newsom, Candace Gaitan, David Pereira | Benchmark Research | Austin, TX |

| | | | |
|---|---|---|---|
| Stuart H. Cohen, MD | Curtis Blankenship, Katelyn Trigg, Courtney Lymuel, Gursimran Mann, Zayan Musa, Hana Minsky, Eliseo Vasquez, Nicole Garza, Kaitlyn Low, Mehrab Hussain, William Li, Rahul Araza, Monique Conover, George Thompson, Hien Nguyen, Scott Crabtree, Bennett Penn, Minh-Vu Nguyen, Archana Reddy, Derek Bays, Kaitlyn Hardin, Matthew Boutros, Alan Koff, Natascha Tuznik, Angel Desai, Naomi Hauser, Sarah Waldman, Gauri Barlingay, Dean Blumberg | UC Davis Health | Sacramento, CA |
| Erin Cooksey, MD | Kristina Harrison, Deanna Cooper, Manisha Horton, Amanda Philyaw | Synexus Clinical Research | Anderson, SC |
| Aurelio Cruz-Valdez, PhD | Janeth, Pacheco-Flores, Anyela Lara, Secia Diaz-Miralrio | Instituto Nacional de Salud Pública | Cuernavaca, México |
| Dima Dandachi, MD, MPH | Tami Day, Britlyn Brown, Taylor Mathews | University of Missouri Health Care | Columbia, MO |
| Matthew G. Davis, MD | Therese Dayton, Joseph I. Mann, Patricia S. Larrabee, Jean C. Kelly, Tia. L Albro, Zerina Zornic, Susan J. Willer, Donna M. Willome, Kathleen K. Ebeling, Jaclyn P. Zona, Julie A. Mooney, Katherine A. Pagenkemper, Victoria F. Fink, Christine N. Hall, Chelsea Bork, Abigail Miller, Mackay Kanaley, Chelsey LoMonaco, Marie Musolino, Jessica Fisher, Katilyn Bergen, Rachel Bordonaro, Cassidy Glod, Liam Sullivan, Brandi Douglass, Ann Casey, Philip LaSpino, Maurice Holmes | Rochester Clinical Research | Rochester, NY |
| Michael E. Dever, MD | Michael Delgado, Tameika Scott, Laverne Denise Davila, Nelisa Frias, Anissa Hilton, Patricia Brown, Shana Caldwell, Martha Hendrix, Edmund Delgado, Mitul Shah, Gracemarie Rosario, Kaneitra Williamson, Taylor Lucier, Jaime Hawat, Matthew Stephens, Monica Cooper, Dante Canidate, Denise Pagan, Sierra Robinson, Pascal Nelson-Quiles, Anthony Perez, Chanel Adams, Keisha Foster, Scott Salmon, Andrew Lockwood, Priya Moorhouse, Paul Yi | Clinical Neuroscience Solutions | Orlando, FL |
| Matthew Doust, MD | Stephanie Catanzaro, Shana Harshell, Madison Mikulak, Bettie, D'Nise Corcoran, Susan DeCraene, Jasmin Redden, Brian DeCraene, Karen Wakefield, Adrian Aljeo, Denise Sample, Clarissa Lara, Stephanie Junker, Nathan Alderson, Kimberly Joshlin, Mia Munoz, Michele Aguirre, Dina Reyes Cordova, Neil Pearson | HOPE Research Institute | Phoenix, AZ |
| Daniel Duncanson, MD | Kelly Kush, Lori Nesbitt, Cora Sonnier, Jennifer McCarter, Thomas Buschbacher, Evie Zavala, Brittany Cooper, Abbey Mannings, Melissa Berrio, Erin Juhl, William Douglas, Timothy Elder, Linda Grover, Colleen Crabbe, Rachel Francis, Jesse Lipnick, Seldon Longley, Michael Rozboril, Madison Duncanson, Jakob Vaes, Michael Costa, Dhruv Panchal, Michelle Hendricks, Sergio Montalvo, Angel Dubois | SIMEDHealth / SIMEDResearch | Gainesville, FL |
| David Ensz, MD | Bruce Rankin, Tavane Harrison, Meagan Miller, Kayla Sturgeon, Jessica Knight, Janet Otto, Monica Salazar, Megan Howard, Carly Deges, Joseph Harris, Rylea Gulick, Melissa Wiseman, Sue Doty | Meridian Clinical Research | Sioux City, IA |
| Brandon Essink, MD | Roni Gray, Christine Wilson, Fritz Raiser, Akossiwa "Essi" Yovogan, Jessica Satorie Tiffany Nemecek, Hannah Harrington, Amy Lett-Brown,Chelsea Steinmetz, Tabitha Campbell, Carrie Essink, Jamie Meyer, Riley Brockman, Melissa Monarrez, Troy Humphries, Wynter Huffman, Brooke Dworak, Raquel Davis, Samantha Nocita, Heidi Smith, Carissa Schejbal, Kayla Flege, Joe Genoways, Jessa Swanson, Avery Dunn, Kevin Grimes, Phillip Astorino, Ashtynn Jarosz, Hailey Harper, Amy Nichols, Azra Bauman, Jessica Fellows, Courtney Heisey, Ginny McNew | Meridian Clinical Research | Omaha, NE |
| Carlos Fierro, MD | Natalia Leistner, Amy Thompson, Celia Gonzalez, Nathan Arthur, Mazen Zari, Mary Easley, Heather Barker, Manyvohn Rinehart, Monica Atwood, Natalya Amrine, Kelly Moen, Kaley Miller, Angela Eichler, Ann Geier, Christa Estrada, Amber Wolf, Denise Essix, Latoria Rios, Kasie Hickert, Kenny Nguyen, Karol Moore, Stefanie Uwah, Kaelyn Howell, Miranda Dean | Johnson County Clin-Trials | Lenexa, KS |
| Kenneth Finkelstein, DO | David Beckmann, Tanya Hutchins, Sebastian Garcia Escallon, Kristen Johnson, Athena Rivera, David Otuada, Jessica Bartlett, Lauren Wade, Tyler Will, Gina Nielsen-Grewe, Anita Suri | Providea Health Partners Elligo Health Research | Evergreen Park, IL |
| Jon Finley, MD | Nathan Segall, Mildred Stull, Michelle Sowell, Michelle Binns, Kiara Tyner, Karen Yangapatty, Elizabeth West, Cynthia Steele, Kwannda Whatley, Hannah Smith, Pamela Talbott, Kimberly Cobb, Donna Toepfer, Jennifer LeBrun, Susan Jones, Patrizia Greene, Cynthia Pinckney, Kim Banaski, Karen Hickson | CRA Headlands | Stockbridge, GA |
| Diana F Florescu, MD | Mark Rupp, Daniel Brailita, Adia Sikyta, Erica Stohs, Sara Hurtado Bares, Nada Fadul, Matthew Lunning, Elizabeth Schnaubelt, Molly Ferris, Andrew Buettner, Matthew Palmer, Bailee Lichter, Alison Lewis, Chase Kimberling, Jonathan Beck, Erin Iselin, Kimmai McClain, Andrew Schnaubelt | University of Nebraska Medical Center | Omaha, NE |
| Patrick Flume, MD | Gary Headden, Brandie Taylor, Ashley Warden, Amy Chamberlain, Kim Spencer, April Rasberry, Angela Millare, Angel Darrow, Abbey Grady, Max Lento, Allison Patterson, Caitlan LeMatty, Jhonatan Diaz, Andrew Stephens, Emalee Wood, Destri Eichman, Annie Cribb, Annelise Kauffman<br>Charnele Handy, Elizabeth Poindexter, Moira Chance, Anna Miller, Elizabeth Dickinson, Andrea Boan, Erin Klintworth | Medical University of South Carolina | Charleston, SC |
| Veronica Garcia-Fragoso, MD | Maria Gabriela Becerra, Cecilia Mckeown, Lisa Holloway, Toni White, Bonnie Colville, Frederic Santiago, Teresa Becker, Shakira Barr, Chen Ho Yang, Tracy Kowalski, Danitra Glasper, Diana Chehab Nazanin Zarinkamar, Joanna Quezon, Maryam Rabbani, Sadaf Batla, Ayla Perez, Berenice Ferrero, Dean Jang, Biman Goswami, Dustin McFadden, Elton Oliveira, Enya Rentas-Sherman, Julian Edmonson, Laura Plaza-Grisanty, Olga Konshina, Rachely Araujo-Gutierrez, Scott Ward, Teodoro Seminario, Patricia Matute, Sauleha Husain, Akram Assaf, Elisa Moralez, Frances Saubon, Jenny Torres, William Fernandez, Ashraf Jafri, Amy Anderson, Saji Mathew Perinjelil, Waheeda Sureshbabu, Kara Sikes, Joel Cano, Kendra Rogers, Quiana Wilson, Karina Sainz, Abdeali Dalal, Leena Mir, Misbah Baloch, Shammarrian Hampton, Crystal Reese, Lucia Almaguer,<br>Felicia Ardoin, Deep Patel, Bernardo Martinez Leal, Faryal Mahmood, Ana Rueda, Norma Gonzalez, Stacey Montero, Chandra Tobin, Abyssinia Moges, Ari Amirkhosravi, Herman | Texas Center for Drug Development | Houston, TX |

| | | | |
|---|---|---|---|
| | Ortiz, Matthew Joseph, Parul Mehta, Zain Rizvi, Diego Carrington, Blessing Feliz-Okoroji, Moez Talpur, Robert Krbashyan, Simeen Khan, Mary Rogers | | |
| George H. Freeman, MD | Esther Laverne Harmon, Marshall A. Cross, Kacie Sales, Catherine Q. Gular, Amanda Fronzaglio, Timothy O'Malley, Zaahin Huq, Jenna Johnson, Jessica Fuggett, Danielle Merian, Rita Quinn | Health Research of Hampton Roads | Newport News, VA |
| David L. Fried, MD | Lynne A. Haughey, Ariana C. Stanton, Lisa Stevens Rameaka | Omega Medical Research | Warwick, RI |
| Christopher Galloway, MD | Candice Montros, Lily Aleman, Samira Shairi, Robert Duran, Wesley Van Ever, Wasilah Suid, Sandra Torres, Taylor Rice, Wanda Estrada, Julie Castillo,Stephanie Cassidy, Ashleigh Ford, Thai Marie, Colon Maldonado, Amedaris Cordero, Zahra Somji, Rachel Morris | Headlands Research | Orlando, FL |
| Cynthia L. Gay, MD, MPH | David Wohl, Michelle Floris-Moore, Michael Herce, Danielle Clement, Arianna Morrison, Jan Busby-Whitehead, Michelle Hernandez, Zachary Willis, Allison, Burbank, Peyton Thompson, Chris Evans, Susan Pedersen, Becky Straub, Samantha Earnhardt, Erin Hoffman, Jonathan Oakes, Tevnan Keller, Victoria Rucinski, Camille O'Reilly, Kelsey Vollmer, Jennifer Rees, April Welch, Patti Vasquez, Joy Wannamaker, Tanailly Giralt Smith, India Pitts, Amanda Beaten, Ebony Harrington, Alex Bradley, Chidinma Okafor, Miriam Chicurel-Bayard, Kristina Shoffner, Polly Tsai, Chelsea Taylor, Susanne Hendersen, Emily Padgett, Debbie Pence, Jane Salm, Matt Campbell, Kirsten Haigler, Ekatherina Diadiuk, Mariam Ramzan, Pamela Miller, Julie Nelson, Nicole Maponga, Carmen Garcia, Charlie McGehee, Gloria Oyediran, Paul Alabanza, William Wolf, Hannah Munro, Rachael Turner, Dana Lapple, Grace Tillotson, Andrew Powell, Mandy Tipton, Catherine Kronk, Oesa Vinesette, Arti Malik, Kirby Caraballo, Maria Stetson, Charles West, Erin Cardot, Andy Thorne, Maria Bullis, William Zhao, Jennifer Thompson, Kristen Gray, Sarah Law, Holly Milner, Frederick Asamoah, Daniel Galeana, Marcia Gibson, Caressa Goss, Pamela Jones, Joshua Lee, Cheryl Hendrickson, Rachel Cook, Erin Daniel, Centhla Washington, Carolina Pastrana-Medina, Dayo Nylander-Thompson, William Johnson, Eliza Debose, Chloe Twomey, Rachel White, Grace Bailey, Hayley Meier, Jennifer Te Vazquez, Ascary Arias, Allison Castillo, Dynesha Perry, Gwen McKnight, Lucie Mangala, Jessica Gingles, Maggie Harman, Marie Oriol, Sean McMurray, Christy Litel, Noshima Darden-Tabb, Yerson Padilla, Danna Frederick | University of North Carolina | Chapel Hill, NC |
| Bruce C. Gebhardt, MD | Padma N. Mangu, Debra Beck Schroeck, Rajesh Kumar Davit, Gayle D. Hennekes, Donna Percy | Sterling Research Group | Cincinnati, OH |
| Carl P. Griffin, MD | Raymond Cornelison, Shanda Gower,William Schnitz, Angela Genovese, Ryan Morgan, Destiny S. Heinzig-Cartwright, April Green, Kim Hamilton, Chalimar Rojo, Lacey Dietz, Sharee Wright, Aja George, Karen Hames, Sharla Lister, Brandy Ball, Andrea Romero, Krystal Hightower, Dalia Tovar, Kim Calloway, Samelia Farni, Chris Hyatt, Linda Lopez, Kathi Shaw, Natacha Tull, Katelyn Hughes, Selwyn Oruh, Lauren Schwab, Samantha Ting | Lynn Health Science Institute | Oklahoma City, OK |
| Joseph Grillo, MD | Amy Potts, Julie White, Carla Bender, Debra Daugomah, Caitlin Harris, Brian White, Alannah Hill, Chelsea Lairson, Karen Blevins | Medical Research International | Oklahoma City, OK |
| Bernard Grunstra, MD | Donald Quinn, Shelby Olds, Phillip Claybrook, Amy Dye, Shai Perry, Joshua Bullen, Jennie Eller, Sandy Daggs, Nicole Everhart, Dennis Lee, Farrah Fuston | PMG Research of Bristol | Bristol, TN |
| Greg Hachigian, MD | Deborah Murray, Michael Cancilla, Logan Ledbetter, Masaru Oshita | Benchmark Research | Sacramento, CA |
| Charles Harper MD | Keith Vrbicky, Chelsie Nutsch, Sally Eppenbach, Wendell Lewis, Alisha Kiepke, Misty Appeldorn, Cyla Rohde, Catherine King, Kayla Andal, Ashley Frisch, Courtney Green, Kelsey Kelley, Katlyn Mace, Jordan Suckstorf, Torie Johnson, Linden DeBoer, Christy Lee, Eric Graber, Jeni Hoppe, Jill Smith, Heather Ebel, Taysha Hingst, Samantha Wieseler, Diahn Pekny, Elijah Schantz | Meridian Clinical Research | Norfolk, NE |
| C. Mary Healy, MD | Chianti Wade Bowers, Chanei Henry, Sheri Ordonez, Janet Brown, Cathy Faw, Shetel Anassi, Trent Davis, Kim Taylor | Baylor College of Medicine | Houston, TX |
| Jeffrey A. Henderson, MD, MPH | Jeffrey A. Henderson, MD, MPH, Marcia O'Leary, RN, Kendra Enright, RN, Jill Kessler, MS, Pete Ducheneaux, LPN, Asha Inniss, MS, APRN | Black Hills Center for American Indian Health | Rapid City, SD |
| Ripley Hollister, MD | Jeremy Brown, Melody Ronk, Jill York, Shelby Pickle, Jami Wagner, Lisa Jackson, Felipa Ramdeholl, Angelica Romero | Lynn Institute of the Rockies | Colorado Springs, CO |
| Matthew Hong, MD | Wayne Harper, Lisa Cohen, Priti Patel, Kendra Lisec, Makayla Dutton, Lynn Eckert, Aubrey Farray, Jenee Jiggetts, Emily Reilly, Jill Holmes, Aaron Deaver, Christine Grissom, Judith Shand, Brianca Farmer, Eric Henderson, Kristen Shireman, Brad Muskelley, Franziska Gassaway, Darian Lawrance, Sabine Ucik, Toni Bland, Katedra Dixon, Reginald Santiago, Caroline Zhu, Kathleen Sander, Brian Joseph, Marsha Peery, Lori Bridges, Sadia Khan, Adnan Nasir, Sofia Sequiera, Raquell Messick, Kyra Brown, James Hull | M3-Wake Research | Raleigh, NC |
| Gregory Holt, MD | Jennifer Denizard, Juanita Johnson, Sehrish Sikandar, Gisel Urdaneta, Silvana Cobain, Melyssa Sueiro, Precious Leaks, Evelyn Guadalupe, Rochelle Thompson, Dexter Peart, Leidi Paez, Krystal Hosang, Runxia Tian, Ali Vaeli Zadeh | Miami Veterans Affairs Medical Center | Miami, FL |
| Lilly Immergluck, MD | LaKesha Tables, Harold Gene Stringer, Jacquelyn Ali, Cristina Wilson, Noor Mohamed, Kay Woodson, Tiffany White | Morehouse School of Medicine | Atlanta, GA |
| Michael Jacobs, MD | Kathleen Menasche, Vincent Mirkil,Yazil Ramirez, Michael Yee, Laura Elio, Candice Garcia, Azucena Valdovino, Cristina Garcia, Sharla Peahi-Ching, Kristina Arcos | Alliance for Multispecialty Research | Las Vegas, NV |
| Jeffry Jacqmein, MD | Maggie Bowers, Dawn Robison,Victoria Mosteller, Janet Garvey, Alpa Patel, Darlene Bartilucci, Kenneth Aung-Din, Margaret Gannaway, Carolyn Tran, Michael Koren, Mitchell Rothstein, Sonia Gerardo, Cassie Lawler, Yvonne Douglas, Chris Ganzhorn, Emery Noles, Angela Morris, Lisa Carl, Andrea West, Laura Little, Ramil Castillo, Abbey Ras, Nalini Jones, Annan Nurrenbern, Deirdre Arrington, Jacob Wolfer, Brenda Anderson, Amanda Elwood, Amber DeVries, Cara Seifart, Jimmy Knowles, Vy Dang, Mary Strickland, Pam | Jacksonville Center for Clinical Research | Jacksonville, FL |

| | | | |
|---|---|---|---|
| | Garmon, Caron Whitelaw, Sharon Smith, Ivy Guillermo, Nate Grant, Khatija Hussein, Caron Whitelaw RN, Bernadette Moineau, Robert Nix | | |
| Robert Jeanfreau, MD | Susan Jeanfreau, Katelyn Jackson, Kynisha "Nicki" Johnson, RaeShanta McKendall, Shonna James, Calisha Sadiq, Susan Tortorich, Lori Goins, Steven Darden, Melissa Spedale, Kristen Robinson, Joseph Favret, Yordanka Koleva, April Spears, David Conroy | MedPharmics | Metairie, LA |
| William Jennings, MD | Hilario Alvarado, Michele Baka, Malina Regalado | Synexus Clinical Research | San Antonio, TX |
| Carol Kauffman, MD | Andrea Starnes, Andrea Woods, Karen Brudzinski | VA Ann Arbor Healthcare System | Ann Arbor, MI |
| Colleen F. Kelley, MD, MPH | Carlos del Rio, Sheetal Kandiah, Catherine Abrams, Erin Andrew, Felicia Atkinson, Erica Baker, Juliet Brown, Tucker Colvin, Natasha Renee Cook, Meena Dhir, Christopher Foster, Ronald Gaston, Gabriela Gerogial, John Gharbin, Betsy Hall, Valarie Hunter, Aastha KC, Kelly Likos, Bezuayehu Mandefro, Myles Mason, Humberto Orozco, Isaac Perez, Philip Powers, Christin Root, Brittany Spiegel, Pamela Weizel, Sarah Wiatrek, Felicia Wright | Ponce de Leon Center | Atlanta, GA |
| Jeff Kingsley, DO | April Pixler, LaKondria Curry, Sarah Afework, Austin Swanson, Alyssa Middlebrook, Christine Senn, Keyrhea Ritter, Katlin Salewski, Sierra Holmes, Jean Niles, Taylor Hernandez, Lacey Shaw, Kaila Maddox, Klarissa Bohnstedt, Emily Gilder, Cassandra Motley, Alyssa Middlebrook, Hephzibah Udo, Mattison Sherer, Wayman Petty, Joseph Surber | IACT Health | Columbus, GA |
| Judith Kirstein, MD | Marcia Bernard, Erica Sanchez, Nolan Mackey, Clarisse Baudelaire, Hanna He, Brenda Delgado, Brandon Steppe, Bonnie Goodale, Nicole Abels, Carol Remigio, Dipal Patel, Emily Zacarias, Nuvia Espinoza, Esmeralda Machado, Katia Talamante, Lizeth Romero | Velocity Clinical Research | Banning, CA |
| Susan Kline, MD, MPH | Sara Eischen, Rebecca Cote, Diondra Howard, Editha Jordan, Joyce Bolea, Annie McFarland, Asfaw Mesfin, Andrew Snyder, Darlette Luke, Derek LaBar, Theresa Christiansen, Beth Jorgenson, Christina Glasgow, Melissa Schedler | University of Minnesota | Minneapolis, MN |
| Mark E. Kutner, MD | Mark E. Kutner, Jorge Caso, Janet Mendez, Maria Hernandez, Carmen Amador, Amanda G. Colina, Alain Chang, Alondra Diaz, Arael Ayala, Carmen Ballester, Claudia Rodriguez, Dalila Del Valle, Eduardo Rodriguez, Gloria Moreno, Jennifer Ortega, Jhobana Vargas, Jonathan Fernandez, Juan Carlos Delgado, Laura Gonzalez, Leidy Montoya, Marianela Carvajal, Mariete Rendon, Maury Santos, Michelle Browne Mirnaya, Mujica, Neiner Enriquez, Noelio Hernandez, Paola Garcia<br>Raydel Valdes, Saray Carvajal, Susel M. Figueredo, Vanessa Hechevarria, Yanelis Dominguez, Yusleidy Diaz | Suncoast Research Group | Miami, FL |
| Mark Leibowitz, MD | Fernanda Morales, Rosario Sanchez, Mike Delgado, Norma Vega, Nelly Ayala, Iliana Gallaga, Cassandra Celis, Jennifer Muniz, Mariela Quiroz, Juan Frias, Rea Abaniel, John Nelson, Maricor Grio, Alejandro Moreno, Coralia Soto, Jose Espino, Daniel Vargas, Stephanie Lopez | National Research Institute | Los Angeles, CA |
| Derek Lewis, MD | Fred Newton, Aeiress Duhart, Breana Watkins, April Green, Chala Simpson, Briana Dean, Brandy Ball, Shakita Stevenson, Lashonda Stephenson | Lynn Institute of the Ozarks | Little Rock, AR |
| Gregg Lucksinger, MD | Jaleh Ostovar, Audrey Kuehl, Viviana Juncal, Avery Kerwin | Velocity Clinical Research | Medford, OR |
| Benjamin J. Luft, MD | Jorge Alves, Melissa Carr, Ryan Chacon, Barsha Chakraborhy, Aymon Faizi, Laurel Gumpert, Andrew Handel, Kayla Henkel, Erin Infanzon, Andrew Kanner, Lily Limsuvanrot, Michelle Miroddi, Jeanine Morelli, Sharon Nachman, Rena Nanan, Alexander Newman, Alison Pellecchia, Trisha Rush, Jennifer Russell, Stephanie Santiago-Michels, Jonathan Sicoli, Candace Smith, Michael Truhlar, Bruno Valenti, Jennifer Valentine, Kathy Vivas, Yasmine Brown-Williams | Stony Brook University - Stony Brook Medicine | Commack, NY |
| Siham Mahgoub, MD | Alice Ukaegbu, Immaculate Okonkwo, Shannon Gopaul, Tara Gibbons, Yuanxiu Chen, Debra Ordor, Linda Fletcher, Megan Ware, Florencia Gonzalez, Michael Perini, Carla Williams, Mulu Mengistab, Robert Postell, Yejide Obisesan, Adetokunbo Adedokun, Reyneir Magee, Jeremy Smith, Edward Bauer, Lora Collins, Urelda Allman, Deborah Clements, Sarah Shami, Nathaniel Blaboe, Pedro Lima, Michael Crawford | Howard University Hospital / Howard University College of Medicine | Washington, DC |
| Mary Beth Manning, MD | Toby Briskin, Denise Roadman, Sarah Dzigiel, Jennifer Gaston, Brooke Glivar, Brianna Arman, Briana Jackson, Brian Sharpe, Naqib Ahmad, Nicole Baitt | Velocity Clinical Cleveland | Cleveland, OH |
| Rickey D. Manning, MD | James Wilson Hurst, Rodney E. Sturgeon, Paul H. Wakefield, John A. Kirby | Accellacare | Knoxville, TN |
| Kristen Marks, MS, MD | Marshall Glesby, Roy Gulick, Timothy Wilkin, Ole Vielemeyer, Mary Vogler, Carrie Johnston, Rebecca Fry, Daniel Finn, Caitlin Rhoades, Noah Goss, Shaun Barcavage, Valery Hughes, Jonathan Berardi, Ashley Machado, Caique Mello, Mia Crowley, Monique Williams, Minkyung Lee, Mary Ann Zwiebel, Patrice Weller, Antonio Rivera-Lopez, Harrison Chan, Ruby Lee, Victoria Lesina, Vasilika Koci, Paul Kim, Steven Wang, Malissa Robinson, Edward Kenny, Danny Garcia, Venus Fernandez, Parul Shah, Celine Arar, Byron Bullough, Jonattan Rodriguez, Jessenia Fuentes, Jiamin Li, Arthur Goldbach, Genessi Rodriguez, Catherine Jerry, Nadi Islam, Madeline Gomez, Rajshri Hirpara, Ioanna Pahountis, Wayne Burns, Tahera Begum, Gianna Resso, Sophia Alvarez, Elizabeth Connolly, Roxanne Rosario, Sierra Derti, Britta Witting, Anna Gwak | Weill Cornell Chelsea CRS | New York, NY |
| Paul G. Matherne, MD | Cassie Beeks, Sarah Bowen, Deven Fejka, Nicole Guttierrez, Lakeyla Bates, Pam Taylor, Gigi Benoit, Micki Le | Medpharmics | Gulfport, MS |
| Monica McArthur, MD, PhD | Cheryl Young, Helen Powell, Levis Contreras, Panagiota Komninou, Christine Wade, Jumoke Oladapo, Kaitlin Mason, Robin Barnes, Leslie Howe, Cheilon Bolanos, Shannon Bittner, Elva Valle-Maldonado, Wanda Somrajit, Biraj Shrestha, Justin Ortiz, Nancy Greenberg, Kathleen Strauss, Lisa Chrisley, Melissa Billington, Sudhaunshu Joshi, Lavida Porter, Megan McGilvray, Daryl Grays, Shirley George, Jennifer Marron, Kelly Brooks, Natelaine Fripp, Mardi Reymann, Brenda Dorsey, Patricia Farley, Melissa Myers, Natasha Harris, Alyson Kwon, Marcela Pasetti, Daniel Cohen, Myounghee Lee, Laura Liberman, Sherry McCammon, James Campbell, Ana Herbert, Julia Silva, DeAnna Friedman-Klabanoff, Alythia Vo, Jennifer Winkler, Lisa Turek, Colleen Boyce, Anne Thurston, | University of Maryland School of Medicine | Baltimore, MD |

| | | | |
|---|---|---|---|
| | Daniele Nitkowski, Ginny Cummings, Sandra Molina, Susan Holian, Matthew Laurens, Rekha Rapaka, Megan Deming, Mark Travassos, Kirsten Lyke, Henry Seifert, George Escobar, Norma Martinez, Abigail Arias, Ana Maria Davila, Dolores Fontalvo, Elsa Aracely Vargas, Elva Jaldin, Gladis Lopez, Irma Justiniano, Judith Tenezaca, Julio Fernandez, Luz de Maria Osorio, Maria de los Angeles Pichardo, Morena Lemus, Maria Elena Rocha, Rosa Angelica Vigil, Sandra Herrera | | |
| R. Scott McClelland, MD, MPH | Devinder Garcha, Christopher P. Hawk, Bonnie Duran, Donna Lodge-Moore, Leigh Tao, Cristina J. Toledo-Cornell, Mona Jalili, Susan Lottimer, Sheila Samra, Seslee Alsup, Karlee Cooper, Theresa George-Greene, Spencer Hanson, Joni Hensley, Emily Barnett Highleyman, Jewell Jefferson, Jessica Lane, Jessica Long, Alex Martinez, Kerri Sloan, Kelly Smith, Kristee Lewis, Tara Babu, Dwyn Dithmer, Matthew Dustrude, McKenna Eastment, Emily Ford, Abir Hussein, Christine Johnston, Pamela Kohler, Debra Metter, Thepthara N. Pholsena, Meredith Potochnic, Tara Reid, Miko Robertson, Michelle Sabo, Helen Stankiewicz Karita, Jina Taub, Dana Varon, , Brian Wood, Alyssa Braun, David Crawford, Mark Drummond, Jess Heimonen, Lawrence Hemingway, Madelaine Humphreys, Bianca Kalia, Mary Kirk, Taylor Krause, Ray Larsen, Gisella Logioia, Cristina Luevano Santos, Anya Mathur, Lindsey McClellan, Jessica Moreno, Nicole Roed, Matthew Seymour, Katie Wicklander | University of Washington & Lummi Tribal Health Center | Seattle, WA |
| Bruce McClenathan, MD | Mary Hussain, Aaron Poch, Amy Santangelo, Anne Poch, David De Blasio, Evelyn Lomasney, Jacob Turnquist, Kathryn Lago, Laurie Housel, Sherry Lamberth, Sheryl Bedno, Lauren Blevins, Laura Brown, Alice Clay, Gervon Collins, Kaitlyn Covington, Amy Davis, Patricia Davis, Nicole Friedberg, Lacey Gazlay, Helen Gooden, Evelyn Hall, Kim Locklear, Kayla Majors, Shamona McRae, Bryce Meerhaeghe, Kendalyn Stephens, Jade Tran, Lisette Watkins, Katie Williams, Kema Matthews, Karrie Greive, Brandi Carroll, Amanda Williams, Brittany Garner, DeLisa Crosby, Jennifer Ritschl, Jamie Frahm, Karen Stewart, Priti Patel, Dilay Uras, Allison Northrop, Anika Uson, Olyvia Ray, CynDavia McKoy, April Beals, Deidre Turner, Christina Spooner | Womack Army Medical Center | Fort Bragg, NC |
| Mark McKenzie, MD | Teresa Deese, Christy Schmeck, Vickie Leathers, Christy Sweet, Misti Earwood, Erica Osmundse, Gisela Heintz, Lilian Nunkuna, Michelle Forgey, Shelly Brooks, Justian Jarrett, Elizabeth Michael, Lisa Guider, Zack Harmon, Diane Sproles, Randy Cooper, Jessica Benvenuto, Stefanie Mullins, Quinetrice Bennett, Corey Flack | WR Clinsearch | Chattanooga, TN |
| Jorge F. Méndez Galván, MD | Adriana Sordo Durán, Martha Yarelli Valencia Mejía, Froylan David Martínez Sánchez, Ana María Piña Rodríguez, Diana Alim Mena Martínez, Melany Susel Fernández Valdez, Laura Ruy Sánchez Guerrero, Ana Fabiola Ruiz Villagrana, Mónica B. Carrascal, Martha Cecilia Gomora Madrid, Anahí García Álvarez, Ismael Delgado Ginebra, Omar Alfonso Heredia Nieto, Yanni Maldonado Ventura, Jonathan E. Ramírez Salazar, Mariela Salgado Zagal, María Fernanda Espinosa García, Yolanda Albor Hernández, Ricardo Antonio González, Germán Alonso Lara, Marbella Rojas Ortega, Bernardo Kleinfinger Chayet, Victor Emmanuel Alva López, Diego Carlos Angel Perez, Alejandro Cortes Meda,Ivonne Hernandez Giron | Centro de Atención e Investigación Médica (CAIMED) | Mexico City, Mexico |
| Rajan Merchant, MD | Anelgine Crans Yoon, Janet Hill, Lucy Ng-Price, Teri Thompson-Seim, Alejandra Cazares Hernandez, Danielle Hornbuckle, Adriane Rubit, Ann Campbell, Dawn Diorio, Adeline Stabler, Jasdeep Shergill, Claudia Gross, Anne Nguyen | CommonSpirit Health Research Institute | Woodland, CA |
| Vicki E. Miller, MD, MPH | Amy Starr, Shiela Varghese, Sonia Guerrero, Monica Murray, Vanessa Gonzales, Blanca Gomez, Zainab Rizvi, Victoria Aguilar, Anna Pena, Madiha Baig, Dustin Watson, Pauline Ngban, Afifah Ayub, Laura Drampou, Shelby Danforth, Diana Avalos, Jacquelyn Gonzales, Ragen Powell, Sajjad Naqyi, Ambily Dileep, Alefiyah Motiwala, Heather Leary, Humera Siddiqui, Miatta George, Kastyn Kelly, Nicole Segura, Maryam Jamil, Husain Motiwala, Sandra Smith, Sally Hussein, Yousra Yousif, Carlyn Robinson, Cannon Lenfield, Luis Leal, Muhammad Irfan, Nayab Croher, Pattie Tate, Sandra Natalia Perez, Fredric Santiago, Syeda Riaz, Arsani Iskandar, Alefya Hussain | DM Clinical Research | Tomball, TX |
| Linda Murray, DO | Christy Delcamp, Monica Hoewt, Kristin Shade, Tara McTigue | Synexus Clinical Research | Pinellas Park, FL |
| David Musante, MD | William P. Silver, Linda R. Belhorn, Nicholas A. Viens, David Dellaero, Shandelle Parker, Andrew Zimmerman, Roger Ordronneau, Bryan Stanislaus, Kevrin Johnson, Megan Dice, Megan Heron, Sarah Wilkerson | M3-Emerging Medical Research | Durham, NC |
| Sherif Naguib, MD | Justin Singletary, Sha-Wanda Richmond, Sarah Omodele, Emily Oppenheim, Jalisha Hemphill, Marqueta Jones, Millat Gedefa, Janean Smith, Bonnie Raufman, Lesley Whitehead, Elia O'Dell, Sarah Omodele, David Taylor, ShaWanda Richmond, Alexis Melson, Justin Singletary | Synexus Clinical Research | Atlanta, GA |
| Joseph Newberg, MD | Laura Pearlman, Reuben Martinez, Victoria Andriulis, Jacquilyn McCormick, Anna Maddox, Rosalinda Vazquez, Nicole Leahy, Marian Padilla, Mary Reyes | Synexus Clinical Research | Chicago, IL |
| Paul J. Nugent, DO | Leonard Singer, Jeanne Blevins, Meagan Thomas, Christine Hull | Synexus Clinical Research | Cincinnati, OH |
| Yaa D. Oppong, MD | Ryan P. Starr, Scott N. Syndergaard, Nafisa Saleem, Cheryl Norris, Nicole Austin, Rozeli Shelly, Md Mashrur Islam Majumder, Annette Bunnells, Michelle Wallace, Avia McClain-Stocker, Rachel Ryan, Katie Wood, Arien Stebbins, Crystal Schmitt, Jeffrey Pemberton, Mitchel Arlidsen, Daniel Tomita, Geraldine McRae, Amy Sheets, Jeanette Mangual-Coughlin,<br>Margo Miller-Smith, Melinda Thomas | Carolina Institute for Clinical Research | Fayetteville, NC |
| J. Scott Overcash, MD | Adrienna Marquez, Hanh Chu, Kia Lee, Kim Quillin, Jordan Coslet, Yashveer Dubbula, Adam Prince, John Rodriguez, Lee Tomatsu, Erin Vawter, Michael Voskanian, Michael Waters, Gina Weaver, Karla Zepeda, Angela Anorve, Gordon Bovee, Jennifer Baker, Laura Castillo, Allie Davis, Jacob Esparza, Andrea Garcia, Jessica Gonzales, Lizette Gonzalez, Ashleigh Lindsay, Erica Marinelli, Cathy Meza, Shandel Odom, Makenna Orel, Grecia Perez, Helen Pu, Cesar Ramirez, Melania Riordan, Deidre Romines, Raquel Taitingfong, Katrina Tyler, Bernadette Wilson | Velocity Clinical Research | La Mesa, CA |

| | | | |
|---|---|---|---|
| Yogesh K. Paliwal, MD | Amit Paliwal, Renu Bhupathy, Krystle Edwards, Sarah Gordon, Cynthia Montano-Pereira, Blanca Gomez, Yazmin Nunez, Cassandra Martinez, Connie Navarrete, Mayra Casas, Ysabel Lopez, Anthony Macias, Alexandria Vasquez, Maria Gomez | Empire Clinical Research | Pomona, CA |
| Isabel Pereira, MD | Gina Rivero, Tracy Okonya, Frances Downing, Paulina Miller, Yasmin Camberos | Synexus Clinical Research | Vista, CA |
| Bruce Rankin, DO | John M. Hill, Steven Shinn, Vivek Rajasekhar, Marshall Nash, Michelle Tutt, Kimberlee Del Campo, Douglas F. Winter, Leandro Fernandez, Melissa Hodges, Michelle Jones, Sean Lemoine, Veronica Walker, Roy D. Richardson, Angeline Petracca. Katina Marchione, Michelle Morgan, Ashley McCaffrey, Amber Vasquez, Amy Houck-Dominy, Angela Hammerle, Antonio Rivera, Claxton Copeland, Crystal Paccione, Diana Toney, Fadhel Alyunis, Jennifer Dittman, Kriston Applewhite, Lora Parahovnik, Over Seijas, Ryan Hobbick, Samantha Watts, Shatonia Fields, Stacie Evans, Teresa Logsdon, Thais Truffa, Tifany Huertas, Vienna Bauer,William Serrano, Daisy Sawyer, Giovanni Urquilla, Tonya Toby, Albert Garcia, Alicia J. Cevera,Jeffery Hood, Hannah Hodges, Melissa Willard | Accel Research Sites | DeLand, FL |
| María José Reyes Fentanes, MD | Pablo Fermín González Limón, Luis Ricardo Acosta Beuló, Paulina Cleer García Valdovinos, Olivia de la Puente Flores, Eduardo Rugama Martel, Ana Gabriela Mier Flores, Ulises Abel Rodríguez Vargas, Diego Guillermo Muñoz Bolaños, Martha Alejandra Alonso Trejo, Elvia Ramírez Gutiérrez, Alberto Aaron del Rosal Medina, Jaime Chavez Baron, Ana Gabriela Guizar Zamora, Felipe Arredondo Saldaña, Juan De Dios Martín Luján Palacios, Juan José Pardo Moreno, Jorge Torres Ferrera, Itzetl Guzman Mendieta | PanAmerican Clinical Research México | Querétaro, Mexico |
| Margaret Rhee, MD | Jeffrey Klein, Katherine Stapleton, Stacy Collins, Dawn Greer, Kelli Meissner, Brenda Moore, Tylene Falkner, Celeste Blazy, Nicole Johnson, Christina Carter, Annette Pangle, Rosamond Hong | Synexus Clinical Research | Akron, OH |
| Robert Riesenberg, MD | Robert Riesenberg, Stanford Plavin, Mark Lerman, Leana Woodside, Maria Johnson | Atlanta Center for Medical Research | Atlanta, GA |
| Barbara Rizzardi, MD | Michelle King, Vanessa Abad, Jennifer Knowles, Benjamin Richeson, Denise Pessetto, Heather Holtman, Lori Luth, Wyatt Walsh, Andrea Johnson, Dreama Fackrell, Patrick O'Keefe, Sara Isolampi, Michelle Walkingshaw, Josh Carrillo, Renu Landage, Stephanie Wallace | Velocity Clinical Research | West Jordan, UT |
| Carina A. Rodriguez, MD | Patricia Emmanuel, Lucy Guerra, Asa Oxner, Alicia Marion, Reed Ryan, Tiffany Vasey, Susannah Hall, Amanda Morton, Emma Gonzalez, Elisabeth Ballans, Rachel Karlnoski, Luz Santamaria, Rosalinda Cruz, Joshua Finley, Michael Hayes, Oliver Emberger, Mark Pennington, Meghana Vankatesh, Kimberly Johnson, Marina Wassif, Janelle Perkins, Veroniya Winkfield, Amavyvis Garcia, John Jones, Lori Brock, Kyle Cesareo, Dilcina Dragon, Dominic Moore, Catherine Marten, Thi Nguyen, April Roberts, Kristi Bojaxhi, Chrestenie Mouse | University of South Florida, Morsani College of Medicine | Tampa, FL |
| David Rosenberg, MD | Lee Tomatsu, Viviana Gonzalez, Millie Manalo, Nicole Rudin | Pharmacology Research Institute | Los Alamitos, CA |
| Vida Veronica Ruiz Herrera, MD | Vida Veronica Ruiz Herrera, Eduardo Gabriel Vazquez Saldaña, Laura Julia Camacho Choza, Karen Sofia Vega Orozco, Sandra Janeth Ortega DominguezMaria, Carolina Molina Roman, Julian Camacho Choza, Rodolfo Fabian Lomeli Guerrero, Giuliana Magaña Garcia, Carlos Andres Perez Navarro, Cesar Alberto Lopez Martin, Luis Arturo Rico Godinez, Felipe de Jesus Lopez Cordova, Daniel Arroniz Bernal, Luisana Aldaco Cota, Edgar Cordova Pulido, David Aguila Rivera | PanAmerican Clinical Research México | Guadalajara, Mexico |
| Beth Safirstein, MD | Luz Zapata, Lazaro Gonzalez, Evelyn Quevedo, Farah Irani, Julio Vigil, Steven Rapp, Mark Firestone, Humberto Mucientes, Ali Yasells Garcia, Florence Baum, Robert Hacman, Martha Ravelo, Carlos Alzate, Keyanna Francois, Alberto Napoles, Jamie Lorenzo, Deandra Clarke, Disneydi Gutierrez, Yean Alfonso, Nestor Lopez, Ana Bustos, Ilya Faybishenko, Lynnette Perez, Evelyn Qlies, Maria del Valle, Natalie Joseph, Judith Powell, Jessica Hernandez, Rafael Sanchez, William Torres, Damaris Alonso, Dragos Juravle, Roberto Valledor, Veronica Valledor, Maria Pazos, Teresa Rios, Maria Lascano | MD Clinical | Hallandale Beach, FL |
| Howard Schwartz, MD | Nelia Sanchez-Crespo, Terry Piedra, Barbara Corral, Jennifer Schwartz | Cenexel RCA | Hollywood, FL |
| Elizabeth Secord, MD | Roy Collins, Marita Poff, Jamal Chehab, Sajith Matthews, Thomas Mazzocco, Chantel Karmo, Sarah Meram, Janie Faris, Valerie Mika, Shobi Mathew, Brian O'Neil, James Paxton, Amy Stolinski, Stacie Smith, Benjamin Wasinski, Lisa Palmer, Katherine Cross, Samuel Ceckowski, Theodore Falcon, Jeffrey Harrison, Abe Lovelace, Selmir Mahmutovic | Wayne State University | Detroit, MI |
| Marian E. Shaw, MD | Mark A. Turner, Cory J. Huffine, Esther S. Huffine, Jacqueline Hanson, Nicholas Tuttle, Shannon Veach, Antonio Navarrete, Jammie Smith | Velocity Clinical Research | Meridian, ID |
| Teresa S. Sligh, MD | Scott Sligh, Parul Desai, Vincent Huynh, Carlos Lopez, Erika Mendoza, Dennis Perez, Samuel Ceballos, Jennifer Gomez, Janneth Becerra, Tiffany Martinez, Erika Navarro Fausto | Providence Clinical Research | North Hollywood, CA |
| Joel Solis, MD | Carmen Medina, Westley Keating | Centex Studies | McAllen, TX |
| Jonathan Staben, MD | Jessica Horton, Hannah Neill-Gubitz, Hilary Koenigs, Autumn Dlugas, Stacie Rebar, Anne Reedy, Roslyn Pierce, Kali Karst, Jaimee Gribben, Sarah Troutt, Mimi Meipel, Ann Carson, Paige Ramos, Natosha Hardy, Zack Brownell, Dot Heid, Annie Estes, Andrea Fry, Veronica Navarro | MultiCare Institute for Research and Innovation | Cheney, WA |
| Kathryn E. Stephenson, MD, MPH | Karen A. Lorenc, Audrey B. Nathanson, Michelle Beck, Shaelah M. Huntington, Wendy Hori, Uyen Rasphoumy, Ashley Beckles, Jody Dushay, Vijai Bhola, Wilanda Gabriel, Annika Gompers, Halle Hall, Nicholas Manickas-Hill, Toluwanimi Ajayi, Nicole Magner, Conor Cronin, James Arrico, Heena Patel, Janet Mullington, Michael Seaman, Katherine Yanosick, Ariana Leonelli, Eric Dai | Beth Israel Deaconess Medical Center | Boston, MA |
| Danny Sugimoto, MD | Jeffrey Dugas Sr., Dolores Rijos, Sandra Shelton, Stephan Hong | Cedar Crosse Research Center | Chicago, IL |
| Suzanne Swan, MD | Sharine Phan, Tami Wahlin, Elizabeth Bennett, Amy Salzl, Jeannette Blaisdell, Stacie Mahowald, Dominick Thibodeau, Sophia Houser, Tammy Hanson | Synexus Clinical Research | Richfield, MN |
| Karen Tashima, MD | Helen Patterson, Stacey Chapman, Giselle Pinto, Jennifer Brashears, Evelyn Hipolito, Laura Elmasian, Timothy Flanigan, Joseph Garland, Britt Harrington, Anthony Harrison, Jenny Thai, Mazen Taman, Krista Kiser, Kay Rutherford, Shivani Patel, Jimin Shin, Kim Rapoza, | The Miriam Hospital | Providence, RI |

| | | | |
|---|---|---|---|
| | Sujata Sahu, Kristine Hauser, Kendra Vieira, Elliott Bosco, Christopher Federico, Kanika Malani, Christian Schroeder, Janet O'Connell, Meghan McCarthy, Anna Hippchen | | |
| Barbara S. Taylor, MD, MS | Bhoja Katipally, Jessica Blower, Kimberly Kone Ellis, Heta Javeri, Danielle Dixon, Anna Taranova, Diana Cavazos, Robin Tragus, Irma Scholler, Lisa Longoria, Laura Najvar, Meredith Hosek, Bridgette Soileau, Morgan Brown | University of Texas Health Science Center San Antonio | San Antonio, TX |
| Christine B. Turley, MD | Lewis McCurdy, Tonisha Brown, Martha Pawlicki, Jennifer Reeves, Jona Bauer, Cedrick Griner, Cameron Russell, Veena Sampathkumar, Zeynep Alimchandani, Robin Muller, Tracey Coakley, Mary Sours, Saifelnasr Mohamed, Sone Alanoh, Amy Yeh, Sahra Khan, Eleojo Abutu, Genena Buck, Sarah Hicks, Andreana Alexander, Tammy Patterson, Maria Martilnsalaco, Amy Clontz, Marina Leonidas, Zainab Shahid, Jay I. Patel, Ryan Bender | The Charlotte-Mecklenburg Hospital Authority d/b/a Atrium Health | Charlotte, NC |
| Lisa S. Usdan, MD | Lora J. McGill, Valerie K. Arnold, Carolyn Scatamacchia, Codi M. Anthony, Carol R. Marsh, Cathy T. Houpt, Charles L. Grandberry, Debra A. O'Brien, Kelsey N. Evans, Leslie M. Lazar, Mary J. Williams, Megann F. Fickle, Robyn M. Presley, Shelby R. McWhorter, Julia Sinatra, Irene W. Powell, Tavia S. Flagg, Melissa N. Flowers, Penny J. McCracken, Reagan A. Boone, Dominique L. Ross, Amber J. Jones, LaKeshia N. Pipkin, Victoria J. Neal, Monica Toor, Brandi Gruber, Erin L. Wells, Kelly Iskiwitz, Carolyn J. Scatamacchia, Codi M. Anthony, Lisa S. Usdan, Lora J. McGill, Valerie K. Arnold | Clinical Neuroscience Solutions | Memphis, TN |
| Larkin Tyler Wadsworth III, MD | Horacio Marafioti, Lyly Dang, Lauren Clement, Kristen Johnson, Anya Penly, Elizabeth Garner, Angie Kean, Sophia Bolakas, Andrea Deffenbaugh, Cerece Miles, Lindsay Nooter, Christy Shultz, George Cherniawski, Stephanie Tesson, Ash Dale, Laura Hartupee, Breanna Galibert, Karen Knapp | Sundance Clinical Research | St. Louis, MO |
| Michael Waters, MD | Dalia Tover, Scott Overcash, Jordan Coslet, Michael Voskanian, Giuliano Zolin, Matthew Petro, Gina Weaver, Kia Lee, Hanh Chu, Karla Zepeda, Crystle Rajania, John Rodriguez, Tracey Fabrega, Kaitlyn Sandler, Alex Tapia, Cecilia Barbabosa, Renee Pasion, Jacob Pineda, Rosalynn Landazuri, Angelica Franco, Estee Garcia, Marilynn Rodriguez, Joanna Ocampo | Velocity Clinical Research | Chula Vista, CA |
| Jordan Whatley, MD | Jordan Whatley, Christopher Dedon, Emily Best, Amie Breaux Shannon, Mary Margaret Dobson, Nicole Harrell, Lindsey Kobetz Hall, Kristen LeBleu Losavio, Patricia Whatley, Tana Bourgeois, Alexandra Caillouet, Samantha Brooke McMillon, Amy Thomassie, Donna Michelle Hurst, Michelle Symms, Lyndsea Folsom, Crystal Rowell, Loney Girod, Lauren Sternfels, Makaylea Truitt, Lori Martin, April Mims | Meridian Clinical Research | Baton Rouge, LA |
| Jewel Johnny White, MD | Amanda Occhino, Ruth Paiano, Morgan McLaughlin, Elisa Swieboda | Synexus Clinical Research | The Villages, FL |
| Hayes Williams, MD, PhD | LaShondra Cade, Mitzi Roberts, Aileen Cunningham, Rhodna Fouts, Connie Moya, Gary Boyd, Justina Owens, Abby Wellinghurst | Achieve Clinical Research | Birmingham, AL |
| Clint Wilson, MD | Jason Milligan, Danielle Raley, Joseph Bocchini, Carrie Kay, Shannon Saksa, Courtney Harmon, Ashley Primos, CJ McKenna, Star Roberts | Willis-Knighton Health System / WKB Family Medicine Associates | Bossier City, LA |
| Peter J. Winkle, MD | Amina Z. Haggag, Elizabeth Lee, Michelle Haynes, Marysol Villegas, Sabina Raja, Mary Grace Lejarde, Caroline Villanueva, Natalie Ureno, Jessica Cramer, Steven Garcia, Yesenia Barraza, Ashley Barajas, Lauren Ferreira, Lucy Rems, Zaki Abawi, Damon Pineda, Patricio Ordonez, Gaby Huizar, Lesbia Alarcon, Anna Luz Belarmino, Nenita Llarena, Alberto Heshike, Isabel Rangel, Karen Cruz, Rynel Villanueva, Matthew Rohrig, Han Tran, Axl Dyer, Maria Webb, Akihisa Kodama, Cynthia Juarez, Sandra Gaona, Moriah Wilson, Mark Gonzalez | Anaheim Clinical Trials | Anaheim, CA |
| Patricia L. Winokur, MD | N/A | University of Iowa Medical Center | Iowa City, IA |
| Paul E. Wylie, MD | Renea Henderson, Natasa Jenson, Fan Yang, Amy Kelley, Kelly Knight, Jessica Watson, Stacy Tierney, Emily Knight, Jessica Woosley, Faith Fields, Glen Scott Thrower | Preferred Research Partners | Little Rock, AR |
| Carmen D. Zorrilla, MD | Carmen Irizarry, Gloria Martino, Natalia Muler, Lázaro Valdés | Universidad de Puerto Rico - Recinto de Ciencias Médicas - Maternal Infant Studies Center (CEMI) | San Juan, Puerto Rico |

**Data and Safety Monitoring Board (DSMB) Members List**

| Member | Institution |
|---|---|
| Richard J. Whitley, MD (Chair) | School of Medicine, University of Alabama at Birmingham |
| Abdel Babiker, PhD | MRC Clinical Trials Unit at University College, London |
| Lisa A. Cooper, MD, MPH | Johns Hopkins University School of Medicine and Bloomberg School of Public Health |
| Susan S. Ellenberg, PhD | University of Pennsylvania School of Medicine, Center for Clinical Epidemiology and Biostatistics, Division of Biostatistics |
| Alan Fix, MD, MS | Vaccine Development Global Program, Center for Vaccine Innovation and Access, PATH |
| Marie Griffin, MD, MPH | Vanderbilt University Medical Center |
| Steven Joffe, MD, MPH | University of Pennsylvania, Perelman School of Medicine |
| Jorge Kalil, MD, MPH | Heart Institute, Hospital das Clínicas da Faculdade de Medicina da Universidade de São Paulo, Brazil |
| Myron M. Levine, MD, DTPH | University of Maryland School of Medicine |
| Malegapuru W. Makgoba, MB, ChB, DPhil, FRCP | University of KwaZulu-Natal, South Africa |
| Reneé H. Moore, PhD | Drexel University |
| Anastasios A. Tsiatis, PhD | North Carolina State University |
| Sally Hunsberger, PhD (Executive Secretary) | NIAID, NIH |

#### Objectives and End Points

Some secondary and exploratory objectives/end points (highlighted gray) were not addressed in this manuscript as they are still not yet available, including immunogenicity data and data from the period after blinded crossover. These results will be the subject of subsequent reports.

##### Table S1. Primary, Secondary, and Exploratory Objectives and End Points

| Objectives | End Points |
|---|---|
| **Primary:** | **Primary:** |
| • To evaluate the efficacy of a two-dose regimen of SARS-CoV-2 rS adjuvanted with Matrix-M™ compared to placebo against RT-PCR-confirmed symptomatic Covid-19 illness diagnosed ≥7 days after completion of the second injection in the initial set of vaccinations of adult participants ≥18 years of age. | **Primary End Point:**<br>• First episode of RT-PCR-positive mild, moderate, or severe Covid-19, where severity is defined as:<br>**Mild Covid-19 (≥1 of the following):**<br>• Fever (defined by subjective or objective measure, regardless of use of anti-pyretic medications)<br>• New onset cough<br>• ≥2 additional Covid-19 symptoms:<br>o New onset or worsening of shortness of breath or difficulty breathing compared to baseline.<br>o New onset fatigue.<br>o New onset generalized muscle or body aches.<br>o New onset headache.<br>o New loss of taste or smell.<br>o Acute onset of sore throat, congestion, or runny nose.<br>o New onset nausea, vomiting, or diarrhea.<br>**OR Moderate Covid-19 (≥1 of the following):**<br>• High fever (≥38.4°C) for ≥3 days (regardless of use of anti-pyretic medications, need not be contiguous days).<br>• Any evidence of significant LRTI:<br>o Shortness of breath (or breathlessness or difficulty breathing) with or without exertion (greater than baseline).<br>o Tachypnea: 24 to 29 breaths per minute at rest.<br>o $SpO_2$: 94% to 95% on room air.<br>o Abnormal chest X-ray or chest CT consistent with pneumonia or LRTI.<br>• Adventitious sounds on lung auscultation (e.g., crackles/rales, wheeze, rhonchi, pleural rub, stridor).<br>**OR Severe Covid-19 (≥1 of the following):**<br>• Tachypnea: ≥30 breaths per minute at rest.<br>• Resting heart rate ≥125 beats per minute.<br>• $SpO_2$: ≤93% on room air or $PaO_2/FiO_2$ <300 mmHg.<br>• High flow $O_2$ therapy or NIV/NIPPV (e.g., CPAP or BiPAP).<br>• Mechanical ventilation or ECMO.<br>• One or more major organ system dysfunction or failure to be defined by diagnostic testing/clinical syndrome/interventions, including any of the following:<br>o Acute respiratory failure, including ARDS.<br>o Acute renal failure.<br>o Acute hepatic failure.<br>o Acute right or left heart failure. |

| Objectives | End Points |
|---|---|
| | o Septic or cardiogenic shock (with shock defined as SBP <90 mm Hg OR DBP <60 mm Hg).<br>o Acute stroke (ischemic or hemorrhagic).<br>o Acute thrombotic event: AMI, DVT, PE.<br>o Requirement for: vasopressors, systemic corticosteroids, or hemodialysis.<br>• Admission to an ICU.<br>• Death. |
| **Key Secondary Objective:** | **Key Secondary End Point:** |
| • To evaluate the efficacy of a two-dose regimen of SARS-CoV-2 rS adjuvanted with Matrix-M™ compared to placebo against RT-PCR-confirmed symptomatic Covid-19 illness due to a SARS-CoV-2 variant not considered as a "variant of concern / interest" according to the CDC Variants Classification, diagnosed ≥7 days after completion of the second injection in the initial set of vaccinations of adult participants ≥18 years of age. | • First episode of RT-PCR-positive Covid-19, as defined under the primary end point, shown by gene sequencing to represent a variant not considered as a "variant of concern / interest" according to the CDC Variants Classification. |
| **Other Secondary Objectives:** | **Other Secondary End Points:** |
| • To evaluate the efficacy of a two-dose regimen of SARS-CoV-2 rS adjuvanted with Matrix-M™ compared to placebo against RT-PCR-confirmed moderate-to-severely symptomatic Covid-19 illness diagnosed ≥7 days after completion of the second vaccination in the initial set of vaccinations of adult participants ≥18 years of age.<br>• To assess VE against ANY symptomatic SARS-CoV-2 infection.<br>• To assess VE according to race and ethnicity.<br>• To assess VE in high-risk adults versus non-high-risk adults (high-risk is defined by age ≥65 years with or without comorbidities or age <65 years **with** comorbidities [e.g., obesity (BMI >30 kg/m$^2$), chronic kidney or lung disease, cardiovascular disease, and diabetes mellitus type 2] and/or by life circumstance [living or working conditions involving known frequent exposure to SARS-CoV-2 or to densely populated circumstances (e.g., factory or meat packing plants, essential retail workers, etc.)]).<br>• To assess the durability of vaccine efficacy (measured by all defined efficacy end points) in initial active vaccine recipients versus crossover (delayed) active vaccine recipients.*<br>• To describe the humoral immune response to vaccine in terms of neutralizing antibody to SARS-CoV-2 for all Immunogenicity Population Participants, and for subsets with and without prior SARS-CoV-2 exposure determined by detectable anti-NP antibodies at baseline.*<br>• To assess the immune response to vaccine by IgG antibody to SARS-CoV-2 S protein and hACE2 inhibiting antibodies at Day 35 and later for all Immunogenicity Population participants, and for subsets with and without prior SARS-CoV-2 exposure determined by detectable anti-NP antibodies at baseline.*<br>• To assess the durability of immune response (IgG antibody to SARS-CoV-2 S protein, hACE2 | • First episode of RT-PCR-positive moderate or severe Covid-19, as defined under the primary end point.<br>• ANY symptomatic SARS-CoV-2 infection, defined as: RT-PCR-positive nasal swab and ≥1 of any of the following symptoms:<br>o Fever.<br>o New onset cough.<br>o New onset or worsening of shortness of breath or difficulty breathing compared to baseline.<br>o New onset fatigue.<br>o New onset generalized muscle or body aches.<br>o New onset headache.<br>o New loss of taste or smell.<br>o Acute onset of sore throat, congestion, or runny nose.<br>o New onset nausea, vomiting, or diarrhea.<br>• Neutralizing antibody titers from Immunogenicity Population at Days 0, 35, and immediately prior to administration of the crossover set of vaccinations.*<br>• Serum IgG levels to SARS-CoV-2 S protein, hACE2 inhibition titers from Immunogenicity Population at Days 0, 35, and immediately prior to administration of the crossover set of vaccinations.*<br>• Serum IgG levels to SARS-CoV-2 S protein, MN, and hACE2 inhibition titers from Immunogenicity Population at Months 12, 18, and 24.*<br>• Description of course, treatment, and severity of Covid-19 reported after a RT-PCR-confirmed case via the Endpoint Form.<br>• Reactogenicity incidence and severity (mild, moderate, or severe) recorded by all participants on their eDiary on days of vaccination and subsequent 6 days (total 7 days after each vaccine injection in the initial set of vaccinations).<br>o Reactogenicity end points include injection site reactions:<br>▪ Pain.<br>▪ Tenderness.<br>▪ Erythema. |

| Objectives | End Points |
| --- | --- |
| inhibition, and MN) at 12, 18 and 24 months of study in all Immunogenicity Population participants, and for subsets with and without detectable anti-NP antibodies at baseline or prior to crossover set of vaccinations.<br>• To describe and compare the safety experience for the vaccine versus placebo in adult participants ≥18 years of age based on solicited short-term reactogenicity by toxicity grade for 7 days following each vaccination (Days 0 and 21) after the initial set of vaccinations.<br>• To assess overall safety through 49 days (28 days after second injection of each set of vaccinations [initial and crossover]) and to compare vaccine versus placebo for all unsolicited AEs and MAAEs.<br>• To assess the frequency and severity of MAAEs attributed to vaccine, AESIs, or SAEs through the EoS and to compare vaccine versus placebo after each set of vaccinations (initial and crossover).*<br>• To assess all-cause mortality in vaccine versus placebo recipients after each set of vaccinations (initial and crossover).*<br>• To describe the severity and course of Covid-19 in vaccine versus placebo recipients in terms of healthcare requirements, utilization, and medical assessments after each set of vaccinations (initial and crossover).*<br>• To assess the proportion of participants (vaccine versus placebo recipients) with SARS-CoV-2 infection determined by anti-SARS-CoV-2 NP antibodies, including specifically asymptomatic infection, across the 2 years of study follow-up.*<br>• To assess the VE against SARS-CoV-2 infection determined by anti-SARS-CoV-2 NP antibodies, regardless of whether the infection was symptomatic.*<br>• To assess in a subset of participants the immunogenicity of a new lot of SARS-CoV-2 rS with Matrix-M™ adjuvant in comparison to the lot utilized in the initial set of vaccinations (i.e., immunobridging).* | ▪ Swelling/induration.<br>o Systemic reactions:<br>▪ Fever.<br>▪ Malaise.<br>▪ Fatigue.<br>▪ Arthralgia.<br>▪ Myalgia.<br>▪ Headache.<br>▪ Nausea/vomiting.<br>• Incidence and severity of MAAEs through 49 days, i.e., 28 days after second injection of each set of vaccinations (initial and crossover).<br>• Incidence and severity of unsolicited AEs through 49 days, i.e., 28 days after second injection of each set of vaccinations (initial and crossover).<br>• Incidence and severity of MAAEs attributed to study vaccine, SAEs, and AESIs through Month 12.*<br>• Incidence and severity of SAEs, MAAEs attributed to study vaccine and AESIs during Month 12 through Month 24 or the EoS.*<br>• Death due to any cause.*<br>• Data points to be collected for healthcare requirements, utilization, and medical assessments from participants who become ill on study will be defined in a separate substudy protocol.*<br>• Antibodies to SARS-CoV-2 NP at Days 0 and 35, immediately prior to administration of the crossover set of vaccinations, and at Months 12, 18, and 24 will be used to determine natural infection and to determine the incidence of asymptomatic infection acquired during study follow-up.*<br>• Antibodies to SARS-CoV-2 NP, regardless of whether the infection was symptomatic.*<br>• IgG antibodies to SARS-CoV-2 rS at approximately 35 days after the first crossover vaccination in approximately 300 active vaccine recipients 18 to ≤64 years of age enrolled at selected study sites.*<br>• Neutralizing antibody response at Day 35 for all adolescent participants seronegative to anti-SARS-CoV-2 NP antibodies at baseline, compared with that observed in seronegative adult participants 18 to <26 years of age from the Adult Main Study (Immunogenicity Population participants before crossover).*<br>• Antibodies to SARS-CoV-2 NP, regardless of whether the infection was symptomatic.* |
| **Exploratory Objectives:** | **Exploratory End Points:** |
| • To evaluate the efficacy of study vaccine compared to placebo against RT-PCR-confirmed symptomatic Covid-19 illness due to a SARS-CoV-2 variant considered as a “variant of concern / interest” according to the CDC Variants Classification, diagnosed ≥7 days after completion of the second vaccination in the initial set of vaccinations of adult participants ≥18 years of age.<br>• To assess cell-mediated response:<br>o Th1 or Th2 predominance after initial set of vaccinations.* | • First episode of RT-PCR-positive Covid-19, as defined under the primary end point, shown by gene sequencing to represent a “variant of concern / interest” according to the CDC Variants Classification.<br>• Th1 or Th2 responses, e.g., IL-2, IL-4, IL-5, IL-13, TNF-α, IFN-γ in whole blood and/or harvested PBMCs prior to and on Day 35 after the initial set of vaccinations.*<br>• Serum samples from a designated subset of up to approximately 4500 Immunogenicity Population participants to be transferred to NIAID for testing and analysis to determine correlates of risk and protection. |

| Objectives | End Points |
|---|---|
| • To contribute to a larger cross-study NIH effort to define correlates of risk and protection against SARS-CoV-2 infection and disease.* <br>• To assess impact of vaccination on nasal viral load in nasal swabs of participants who develop symptoms of possible Covid-19.* <br>• To assess impact of vaccination on asymptomatic SARS-CoV-2 RT-PCR positivity and viral load at the time of the crossover set of vaccinations.* <br>• To describe sequences of the genetic material from SARS-CoV-2 viruses detected in Covid-19 cases to evaluate possible viral mutations that may be associated with breakthrough infections.* | End points will be described in a separate statistical analysis plan developed by external statistics groups (e.g., CoVPN, OWS).* <br>• Quantitative RT-PCR tests may be performed on nasal swabs collected from this trial to assess whether vaccination impacts viral shedding.* <br>• Quantitative RT-PCR tests performed on nasal swabs collected immediately prior to administration of blinded crossover vaccination to assess impact of initial vaccination on frequency of asymptomatic SARS-CoV-2 infection and level of viral shedding.* <br>• Next-generation sequencing of viral genomes detected in nasal swabs tested by RT-PCR to describe the genetic evolution of circulating SARS-CoV-2 strains during the conduct of the study.* |

Abbreviations: AESI = adverse event of special interest; AMI = acute myocardial infarction; ARDS = acute respiratory distress syndrome; BiPAP = bilevel positive airway pressure; BMI = body mass index; CDC = Centers for Disease Control and Prevention; Covid-19 = coronavirus disease 2019; CoVPN = Covid-19 Prevention Network; CPAP = continuous positive airway pressure; CT = computed tomography; DBP = diastolic blood pressure; DVT = deep vein thrombosis; ECMO = extracorporeal membrane oxygenation; eDiary = electronic patient-reported outcome diary; EoS = end of study; $FiO_2$ = fraction of inspired oxygen; hACE2 = human angiotensin-converting enzyme 2; ICU = intensive care unit; IgG = immunoglobulin G; IFN-γ = interferon gamma; IL = interleukin; LRTI = lower respiratory tract infection; MAAE = medically attended adverse event; MN = microneutralization; NIAID = National Institute of Allergy and Infectious Diseases; NIH = National Institutes of Health; NP = nucleocapsid; NIPPV = non-invasive positive pressure ventilation; NIV = non-invasive ventilation; $O_2$ = oxygen; OWS = Operation Warp Speed; $PaO_2$ = partial pressure of oxygen; PBMC = peripheral blood mononuclear cell; RT-PCR = reverse transcriptase-polymerase chain reaction; PE = pulmonary embolism; SAE = serious adverse event; SARS-CoV-2 = severe acute respiratory syndrome coronavirus 2; SARS-CoV-2 rS = severe acute respiratory syndrome coronavirus 2 recombinant spike protein nanoparticle vaccine; SBP = systolic blood pressure; $SpO_2$, oxygen saturation; Th1 = type 1 T helper; Th2 = type 2 T helper; TNF-α = tumor necrosis factor alpha; VE = vaccine efficacy.

* Objectives/end points not addressed in the interim report due to incompleteness or yet unavailable data are noted in the table; this includes immunogenicity data and data from the period after blinded crossover.

### Supplemental Methods

#### Recruitment Strategy

At least 25% of the study population was originally intended to be in the ≥65 years age group; however, availability of vaccines under the Emergency Use Authorization (EUA) during the first weeks of the study required the reprioritization of this population for public health and ethical reasons. Prioritization was given to enrollment of individuals at overall high risk for Covid-19, e.g., high risk for acquisition of Covid-19 of any severity due to living circumstances common to the Black/African American or American Indian/Alaska Native communities (including Native Americans of Mexican origin), Hispanic of Latino ethnicity, or other living or working conditions involving known frequent exposure to SARS-CoV-2 (e.g., factory or meat packing plants, essential retail workers, etc.), or at high risk of developing severe Covid-19 complications by virtue of comorbid conditions (e.g., obesity [BMI >30 kg/m$^2$], chronic kidney or lung disease, cardiovascular disease, and diabetes mellitus type 2).

#### Safety Assessments

Following collection of sufficient safety data to support application for EUA, i.e., 2 months' duration of safety follow-up, participants were scheduled for administration of 2 injections of the alternate study material 21 days apart ("blinded crossover"). That is, initial recipients of placebo did receive SARS-CoV-2 rS with Matrix-M™ adjuvant and initial recipients of SARS-CoV-2 rS with Matrix-M™ adjuvant did receive placebo. The same procedure for vaccine administration followed for the initial set of vaccinations was followed at the time of the blinded crossover to ensure that the integrity of the blinded study was maintained.

Solicited AEs of reactogenicity after the initial series of vaccinations was collected via participant reporting in the eDiary utilizing a smartphone application. All participants were trained on the use of these applications, and smartphone devices were provided for those participants who needed them at the initiation of their participation in the study (Day 0).

Safety assessments included collection of participant-recorded solicited (local and systemic reactogenicity) events through 7 days following each injection in the initial set of vaccinations collected via eDiary. Unsolicited AEs and MAAEs were collected through 49 days, i.e., 28 days after second injection of the initial and crossover sets of vaccinations. MAAEs attributed to vaccine, AESIs, SAEs, and investigator-assessed targeted physical examination findings, including vital sign measurements, will be collected through Month 12. Safety follow-up phone calls will be conducted at 3 and 6 months (±30 days) post-crossover to collect MAAEs attributed to vaccine, AESIs, and SAEs in all participants who received crossover vaccinations.

**Safety Monitoring**

Safety was monitored routinely by the Sponsor physicians and routinely by the 2019nCoV-301 Protocol Safety Review Team (PSRT). A centralized DSMB was established in collaboration with NIH, NIAID, Biomedical Advanced Research and Development Authority (BARDA), and Novavax according to the charter dictated by the participating groups. This group reviewed interim unblinded data periodically, made recommendations with respect to safety and emerging efficacy and needed changes to study design. The DSMB was to be informed immediately by the study unblinded statistician if the prespecified stopping boundary was met, indicating that the vaccine caused harm by increasing the rate of mild, moderate, or severe Covid-19. In addition, the DSMB monitored the study for high vaccine efficacy or for futility to detect vaccine activity.

**Prospective Surveillance of Covid-19**

For prospective surveillance, participants were provided with an oral thermometer on Day 0 and instructed to monitor their body temperature daily throughout the first 12 months of the study and to record temperature and any other relevant symptoms daily in their eDiary.

When fever or other specified symptoms were reported in the eDiary for at least 2 consecutive days for the same symptom, participants were directed via the eDiary to begin nasal self-swabbing at home for RT-PCR testing for a total of 3 days and to initiate daily completion of the InFLUenza Patient-Reported Outcome (FLU-PRO) symptom reporting instrument for 10 days after Covid-19 symptom onset or until the participant experienced 2 consecutive asymptomatic days. Participants were instructed at their enrollment visit on the methods of nasal self-swabbing for Covid-19 and completion of the FLU-PRO symptom reporting instrument. The self-swabs to be obtained by the participant were to be maintained according to directions provided in the 3-swab kit, and the designated courier was to be contacted to pick up the kit for shipping to the central laboratory, as directed.

In addition, the eDiary did alert the study site to contact the participant to schedule the in-person Acute Illness Visit for medical evaluation (to include oxygen [$O_2$] saturation and respiratory rate) and medically attended nasal swab. Participants were provided with a pulse oximeter and instructions for measuring and recording their $O_2$ saturation daily at home during their illness. Active surveillance for Covid-19 will continue after the blinded crossover through the first 12 months of study. Passive surveillance of safety and efficacy via remote contacts or the scheduled visits will continue during Months 12 to 24.

Study participants whose home nasal self-swab and/or medically attended nasal swabs were confirmed at the central laboratory to be RT-PCR-positive for SARS-CoV-2 at the Acute Illness Visit were contacted by the study site to arrange a Convalescent Visit. The Convalescent Visit was to occur approximately 1 month (or as soon thereafter, as feasible) after the onset of the RT-PCR-confirmed case of Covid-19 at the Acute Illness Visit to assess status of AEs, record the clinical course of the disease on the End Point Form and obtain a blood sample for convalescent serologic testing.

**Covid-19 End Point Assessment**

To ensure the quality and accuracy of investigator-recorded end point assessments collected on the Endpoint Assessment eCRF page, programmatic checking was performed prior to the data extraction for analysis. The algorithms and the data sources to be used for programming were determined and documented prior to unblinding. Using data elements relevant to the end point definition and captured in the eCRF, programmatic determination of potential end points and associated start date and severity were performed. Data elements used included the participant reported daily symptoms collected on the Daily Illness Symptoms Report eDiary, RT-PCR results by the central laboratory from participant self-swabbing, RT-PCR results by the central laboratory from the swab collected at the Acute Illness Visit, pulse oximeter readings reported by study participants, and pulse oximeter readings collected at the Acute Illness Visit. Disease episodes were constructed programmatically, including date of initial symptoms, date of positive RT-PCR test result, and preliminary severity based on symptoms reported and pulse oximeter readings. The programmatically determined end points were compared to the data collected on the Endpoint Assessment eCRF. Discrepancies such as missing or difference in date that illness started or difference in severity rating prompted Data Management to issue queries to the investigators. The Endpoint Assessment eCRF data, along with the official study RT-PCR results from the University of Washington, Seattle, WA, were used for analysis of the efficacy end points.

Potentially severe cases of symptomatic RT-PCR-positive Covid-19 were reviewed by an external Independent Endpoint Review Committee (ERC) established by the sponsor. The ERC consisted of physicians who have clinical and research experience (e.g., medical review and/or clinical trial experience) in infectious diseases. The committee's structure, responsibilities, and operation were specified within a charter. Potentially severe cases included Covid-19 reported as SAEs, programmatically identified end points consisting of at least 1 pulse oximeter reading ≤93%, and episodes identified as severe on the Endpoint Assessment eCRF. For pulse oximeter readings, when both participant-recorded values and site collected values were present, both were presented, but the site readings were given preference as to clinical utility. Participant profiles (as outlined in the charter) were provided to the committee members for review according to the process outlined in the charter. These participant profiles included demographics, medical history, AEs, concomitant medications, reported daily symptoms, and the Endpoint Assessment eCRF. The external reviewers documented the criteria used for their clinical review of the cases.

The results of the review were to confirm the case as severe or rule that the case was not severe. The outcome of the review for each case was stored in an electronic medical review system. A file was exported from the system and provided to the Biostatistics group for use in analysis. Cases that were ruled as not severe by the committee despite an investigator-entered severe grading were further reviewed and documented by Novavax clinician(s) prior to unblinding to determine the severity to use in analysis. Cases that were ruled as severe by the ERC but not severe by the investigator were analyzed as severe in the analysis.

**Nasal Swabs for Viral Detection**

Nasal swabs of the anterior nares were obtained at the study site on Day 0 (prior to study vaccination), at the Acute Illness Visit, and at the first crossover vaccination visit.

Participants who experienced an SAE of severe Covid-19 any time after Day 0 were to have a nasal swab obtained (by site personnel or other healthcare personnel) to be sent by the study site to the study central laboratory. Such a swab, if obtained, constituted the medically attended nasal swab recorded on the Acute Illness Visit form.

Participants in the adult portion of the trial were instructed at their enrollment visit on the method of self-swabbing for Covid-19 and procedure for arranging transport of swabs to the central laboratory. Quantitative RT-PCR was performed on RT-PCR-positive swabs using the Abbott RealTime RT-PCR to assess viral load and sequencing of viral genetic material detected in nasal swab RT-PCR testing to evaluate viral mutations.

**SARS-CoV-2 RT-PCR Testing**

The real-time reverse transcriptase-polymerase chain reaction (RT-PCR) test being used is the Abbott RealTime SARS-CoV-2 Assay, which was granted EUA by the US Food and Drug Administration (FDA) on March 18, 2020 (https://www.fda.gov/media/136255/download). The testing was performed at the University of Washington. The validation and verification of the Abbott RealTime SARS-CoV-2 Assay analytical and clinical performance has been published.[1] Dry swabs were used and have been validated for this assay with storage at 2-8°C for up to 7 days and then frozen at -80°C. Once the sample is received at the University of Washington, the dry swabs are eluted into Roswell Park Memorial Institute (RPMI) +2% fetal bovine serum (FBS) or phosphate buffered saline (PBS) prior to testing. The University of Washington conducted an analysis validating and verifying the performance of the Abbott RealTime SARS-CoV-2 Assay with varying viral dilutions. The outcome of the analysis was published elsewhere.[2]

**Whole-Genome Sequencing (WGS) and Clade/Lineage Assignment**

The key secondary end point of the trial was the first episode of RT-PCR-positive, symptomatic Covid-19 due to strain shown by gene sequencing to represent a variant not considered as a variant of concern (VOC) or variant of interest (VOI) according to the Centers for Disease Control and Prevention (CDC) SARS-CoV-2 Variant Classifications and Definitions,[3] starting at least 7 days post-vaccination 2 in the initial vaccination period. Baseline RT-PCR-positive samples as well as RT-PCR-positive samples from self-swabbing or the Acute Illness Visit with enough viral load were sent to the University of Washington Virology Laboratory for WGS, using methodology described elsewhere.[4] In case of multiple RT-PCR-positive samples for a given symptomatic episode, that with the highest viral load was chosen for sequencing.

Sequencing analysis included viral clade/lineage assignment using both the Nextstrain clade label system (https://nextstrain.org/blog/2021-01-06-updated-SARS-CoV-2-clade-naming) and the PANGO lineages designation system (https://cov-lineages.org/), and identification as a VOC/VOI/High Consequence, per the CDC (https://www.cdc.gov/coronavirus/2019-ncov/variants/variant-info.html). Evaluation of SARS-CoV-2 infection or disease was available by site and/or geographic region. The classification of variants was conducted by the University of Washington Virology Laboratory and provided for analysis.

**Analysis Populations**

There were 6 main analysis sets used in this trial:

- The **Intent-to-Treat (ITT) Analysis Set** included all participants who were randomized, regardless of protocol violations or missing data. The ITT analysis set was used for participant disposition summaries and were analyzed according to the treatment arm to which the participant was randomized.

- The **Full Analysis Set (FAS)** included all participants who were randomized and received at least 1 dose of study vaccine/placebo, regardless of protocol violations or missing data. Participants who were unblinded with an intention to receive other Covid-19 vaccines were censored at the time of unblinding. The FAS population was analyzed according to the treatment group to which participants were randomized. The FASs were used for supportive analyses. When the efficacy end points were analyzed using FAS, baseline SARS-CoV-2 seropositivity or nasal swab RT-PCR-positivity was ignored.

- The **Safety Analysis Set** included all randomized participants who received at least 1 dose of study vaccine/placebo. Participants in the Safety Analysis Set were analyzed according to the treatment actually received. In cases where information is available that indicated that a participant received both active vaccine and placebo during the initial vaccination series, the participant was analyzed as part of the active group.

- The **Per-Protocol Efficacy (PP-EFF) Analysis Set** included all participants who received the full prescribed regimen of trial vaccine and had no major protocol deviations that occurred before the first Covid-19 positive episode (i.e., participant was censored at the time of the protocol deviation) and were determined to affect the efficacy outcomes, including baseline SARS-CoV-2 seropositivity or nasal swab RT-PCR -positivity. Participants who were unblinded with an intention to receive other Covid-19 vaccines were censored at the time of unblinding. Although the study enrolled participants regardless of SARS-CoV-2 serologic status at the time of initial vaccination, any participants with confirmed infection or prior infection due to SARS-CoV-2 at baseline, by nasal swab RT-PCR or serology (assessed by anti-nucleocapsid [anti-NP]), were excluded from the PP-EFF population. PP-EFF was the primary set for all efficacy end points.
- A second **PP-EFF (PP-EFF-2) Analysis Set** was defined to allow evaluation of baseline serostatus analysis's impact on vaccine efficacy (VE). The PP-EFF-2 Analysis Set followed the same method described in the PP-EFF population with the exception that it included all participants regardless of baseline serostatus (anti-NP serology) or baseline virological status (RT-PCR).

**Statistical Method for Efficacy End Points**

The VE is defined as VE (%) = (1 – RR) × 100, where RR = relative risk of incidence rates between the 2 trial vaccine groups (SARS-CoV-2 rS / placebo). The RR was estimated by exponentiating the treatment group coefficient from a Poisson regression analysis with robust error variance.[5] The age strata was included in the model as a covariate. To assess incidence rates rather than absolute counts of cases, accounting for differences in follow-up times starting at 7 days after the second vaccination among participants, an offset was utilized in the Poisson regression. When a zero was reported in one of the treatment groups compared (e.g., Black or African Americans) or there were fewer than 5 cases total between groups, the exact conditional method was used instead. A two-sided, 95% confidence interval (CI) was constructed around the estimate.

A super superiority of the vaccine efficacy at each analysis was used to determine the success of the primary end point. A hypothesis test with a one-sided Type I error of 2.5% was conducted with the following hypotheses:

$H_0$: VE ≤0.30 (RR ≥0.70)

$H_1$: VE >0.30 (RR <0.70)

Rejection of the null hypothesis demonstrates a statistically significant VE with a lower bound of CI >30%. In order to be considered for EUA by the FDA, a vaccine must show super superiority where there is a minimum VE of 50% and a lower bound of two-sided 95% confidence bound of at least 30%. Based upon the number of primary efficacy end points planned for analysis, a lower bound of more than 30% corresponds with a VE point estimate of at least 50%.[6]

The RR and its CI was estimated using Poisson regression with robust error variance.[5] The generalized linear model with unstructured correlation matrix (robust error variances) was used. The explanatory variables in the model included the trial vaccine group. The dependent variable was the incidence rate of the end point of interest. The robust error variances were estimated using repeated statement and the participant identifier. The age strata were included in the model as a covariate. To account for the censoring in the analysis, the offset was defined as the natural log of the time from the start of follow-up (7 days post-second vaccination) to the outcome of interest or to the end of study in addition to censoring described in the analysis set definitions. Poisson distribution was used with a logarithmic link function. In the case where there were zero end points for one of the vaccine groups or the total number of end points in both treatment groups combined is less than 5, a Poisson model was substituted with an exact conditional binomial method using the Clopper-Pearson method. This method conditions on the total number of events across the treatment groups where the number of events in the active group are generated from a single binomial distribution. The point estimate form this single binomial distribution and the corresponding confidence intervals constructed using the Clopper-Pearson method were transformed back to relative risks.

A Cox proportional hazards model with the age strata as a covariate was also developed as a supportive analysis to the Poisson regression. The model followed the same explanatory and dependent variables as the Poisson model and censored participants based on their follow-up time available.

#### Supplemental Tables and Figures

*Primary Efficacy End Point*

The primary efficacy end point was the first episode of virologically confirmed (nasal swab RT-PCR-positive to SARS-CoV-2), symptomatic mild, moderate, or severe Covid-19 (see definitions in Table S1), with onset ≥7 days after completion of second study vaccination in serologically (to SARS-CoV-2 anti-Nucleoprotein, NP) and virologically (nasal swab RT-PCR) negative participants at baseline.

**Table S2. Symptoms Suggestive of Covid-19**

| |
|---|
| • Fever (body temperature >38.0°C, in the absence of other symptoms) or chills |
| • New onset or worsening of cough compared with baseline |
| • New onset or worsening of shortness of breath or difficulty breathing over baseline |
| • New onset of fatigue |
| • New onset of generalized muscle or body aches |
| • New onset of headache |
| • New loss of taste or smell |
| • Acute onset of sore throat |
| • Acute onset of congestion or runny nose |
| • New onset of nausea or vomiting |
| • New onset of diarrhea |

Abbreviations: Covid-19 = coronavirus disease 2019.

#### Table S3. End Point Definitions of Covid-19 Severity

| Covid-19 Severity | End Point Definitions |
|---|---|
| | **First episode of RT-PCR-positive mild, moderate, or severe Covid-19:** |
| **Mild** | **≥1 of the following:**<br>• Fever (defined by subjective or objective measure, regardless of use of anti-pyretic medications)<br>• New onset of cough<br>• ≥2 additional Covid-19 symptoms:<br> o New onset or worsening of shortness of breath or difficulty breathing compared to baseline.<br> o New onset of fatigue.<br> o New onset of generalized muscle or body aches.<br> o New onset of headache.<br> o New loss of taste or smell.<br> o Acute onset of sore throat, congestion, or runny nose.<br> o New onset of nausea, vomiting, or diarrhea. |
| **Moderate*** | **≥1 of the following:**<br>• High fever (≥38.4°C) for ≥3 days (regardless of use of anti-pyretic medications, need not be contiguous days).<br>• Any evidence of significant lower respiratory tract infection (LRTI):<br> o Shortness of breath (or breathlessness or difficulty breathing) with or without exertion (greater than baseline).<br> o Tachypnea: 24 to 29 breaths per minute at rest.<br> o $SpO_2$: 94% to 95% on room air.<br> o Abnormal chest X-ray or chest computerized tomography (CT) consistent with pneumonia or LRTI.<br>• Adventitious sounds on lung auscultation (e.g., crackles/rales, wheeze, rhonchi, pleural rub, stridor). |
| **Severe*** | **≥1 of the following:**<br>• Tachypnea: ≥30 breaths per minute at rest.<br>• Resting heart rate ≥125 beats per minute.<br>• $SpO_2$: ≤93% on room air or $PaO_2/FiO_2$ <300 mmHg.<br>• High flow oxygen ($O_2$) therapy or non-invasive ventilation (NIV)/non-invasive positive pressure ventilation (NIPPV) (e.g., continuous positive airway pressure [CPAP] or bilevel positive airway pressure [BiPAP]).<br>• Mechanical ventilation or extracorporeal membrane oxygenation (ECMO).<br>• One or more major organ system dysfunction or failure to be defined by diagnostic testing/clinical syndrome/interventions, including any of the following:<br> o Acute respiratory failure, including acute respiratory distress syndrome (ARDS).<br> o Acute renal failure.<br> o Acute hepatic failure.<br> o Acute right or left heart failure.<br> o Septic or cardiogenic shock (with shock defined as systolic blood pressure [SBP] <90 mm Hg OR diastolic blood pressure [DBP] <60 mm Hg).<br> o Acute stroke (ischemic or hemorrhagic).<br> o Acute thrombotic event: acute myocardial infarction (AMI), deep vein thrombosis (DVT), pulmonary embolism (PE).<br> o Requirement for: vasopressors, systemic corticosteroids, or hemodialysis.<br>• Admission to an intensive care unit (ICU).<br>• Death. |

Abbreviations: AMI = acute myocardial infarction; ARDS = acute respiratory distress syndrome; BiPAP = bi-level positive airway pressure; Covid-19 = coronavirus disease 2019; CPAP = continuous positive air pressure; CT = computerized tomography; DBP = diastolic blood pressure; DVT = deep vein thrombosis; ECMO = extracorporeal membrane oxygenation; $FiO_2$ = fraction of inspired oxygen; ICU = intensive care unit; LRTI = lower respiratory tract infection; NIV = non-invasive ventilation; NIPPV = non-invasive positive pressure ventilation; $PaO_2$ = partial pressure of oxygen in the alveolus; PE = pulmonary embolism; SBP = systolic blood pressure; $SpO_2$ = oxygen saturation.

* Participants with a single vital sign abnormality placing them in the moderate or severe categories must also meet the criteria for mild Covid-19.

#### Table S4. Demographics and Baseline Characteristics (Safety Analysis Set)

| Parameter | NVX-CoV2373<br>N = 19,729 | Placebo<br>N = 9853 | Total<br>N = 29,582 |
|---|---|---|---|
| **Age (years)** | | | |
| Mean (SD) | 46.5 (15.05) | 46.8 (14.95) | 46.6 (15.02) |
| Median | 47.0 | 47.0 | 47.0 |
| Min, max | 18 - 95 | 18 - 90 | 18 - 95 |
| **Age group, n (%)** | | | |
| 18 to ≤64 years | 17,251 (87.4) | 8616 (87.4) | 25,867 (87.4) |
| ≥65 years | 2478 (12.6) | 1237 (12.6) | 3715 (12.6) |
| **Sex, n (%)** | | | |
| Male | 10,409 (52.8) | 5038 (51.1) | 15,447 (52.2) |
| Female | 9320 (47.2) | 4815 (48.9) | 14,135 (47.8) |
| **Race, n (%)** | | | |
| White | 14,789 (75.0) | 7384 (74.9) | 22,173 (75.0) |
| Black or African American | 2320 (11.8) | 1167 (11.8) | 3487 (11.8) |
| American Indian or Alaska Native | 1309 (6.6) | 662 (6.7) | 1971 (6.7) |
| Asian | 811 (4.1) | 416 (4.2) | 1227 (4.1) |
| Multiple | 324 (1.6) | 158 (1.6) | 482 (1.6) |
| Native Hawaiian or Other Pacific Islander | 56 (0.3) | 12 (0.1) | 68 (0.2) |
| Not reported | 120 (0.6) | 54 (0.5) | 174 (0.6) |
| **Ethnicity, n (%)** | | | |
| Hispanic or Latino | 4333 (21.9) | 2154 (21.9) | 6487 (21.9) |
| Not Hispanic or Latino | 15,339 (77.7) | 7676 (77.9) | 23,015 (77.8) |
| Not reported | 32 (0.2) | 19 (0.2) | 51 (0.2) |
| Unknown | 25 (0.1) | 4 (< 0.1) | 29 (0.1) |
| **Country, n (%)** | | | |
| United States | 18,553 (94.0) | 9265 (94.0) | 27,818 (94.0) |
| Mexico | 1176 (6.0) | 588 (6.0) | 1764 (6.0) |
| **Occupation, n (%)** | | | |
| Currently working | 13,442 (68.1) | 6705 (68.1) | 20,147 (68.1) |
| Working in close proximity to others | 5019 (25.4) | 2524 (25.6) | 7543 (25.5) |
| Student attending school in person | 1135 (5.8) | 520 (5.3) | 1655 (5.6) |
| **In-person schooling/currently working/ working in close proximity to others, n (%)** | 14,965 (75.9) | 7466 (75.8) | 22,431 (75.8) |
| **Days/week at workplace, n (%)** | | | |
| 0 days/week | 3052 (15.5) | 1609 (16.3) | 4661 (15.8) |
| 1 day/week | 954 (4.8) | 448 (4.5) | 1402 (4.7) |
| 2–4 days/week | 3411 (17.3) | 1729 (17.5) | 5140 (17.4) |
| ≥5 days/week | 6009 (30.5) | 2915 (29.6) | 8924 (30.2) |
| **PPE used by people at workplace** | 10,261 (52.0) | 5078 (51.5) | 15,339 (51.9) |
| **Living situation, mean (SD)** | | | |
| Number of people living with participant | 2.0 (3.65) | 1.9 (3.28) | 2.0 (3.53) |
| Number of co-habitants under 18 years | 0.6 (1.77) | 0.6 (1.38) | 0.6 (1.65) |
| Number of co-habitants 18 to 64 years | 1.2 (2.71) | 1.2 (3.00) | 1.2 (2.81) |

| Parameter | NVX-CoV2373<br>N = 19,729 | Placebo<br>N = 9853 | Total<br>N = 29,582 |
|---|---|---|---|
| Number of co-habitants ≥65 years | 0.2 (0.46) | 0.2 (0.45) | 0.2 (0.46) |
| **Lifestyle, n (%)** | | | |
| History of smoking/vaping | 6180 (31.3) | 3090 (31.4) | 9270 (31.3) |
| Currently smoking/vaping | 3086 (15.6) | 1528 (15.5) | 4614 (15.6) |
| **BMI category, n (%)** | | | |
| Underweight (<18.0 $kg/m^2$) | 142 (0.7) | 60 (0.6) | 202 (0.7) |
| Normal (18.0–24.9 $kg/m^2$) | 5676 (28.8) | 2804 (28.5) | 8480 (28.7) |
| Overweight (25.0–29.9 $kg/m^2$) | 6475 (32.8) | 3243 (32.9) | 9718 (32.9) |
| Obese (≥30.0 $kg/m^2$) | 7339 (37.2) | 3708 (37.6) | 11,047 (37.3) |
| **Comorbidities, n (%)** | | | |
| Obesity (BMI ≥ $kg/m^2$) | 7339 (37.2) | 3708 (37.6) | 11,047 (37.3) |
| Chronic lung disease | 2745 (13.9) | 1434 (14.6) | 4179 (14.1) |
| Diabetes mellitus type 2 | 1517 (7.7) | 813 (8.3) | 2330 (7.9) |
| Cardiovascular disease | 222 (1.1) | 121 (1.2) | 343 (1.2) |
| Chronic kidney disease | 132 (0.7) | 58 (0.6) | 190 (0.6) |
| **Overall high-risk adults,* n (%)** | | | |
| Yes | 18,805 (95.3) | 9387 (95.3) | 28,192 (95.3) |
| No | 924 (4.7) | 466 (4.7) | 1390 (4.7) |
| **High risk for developing severe Covid-19,† n (%)** | | | |
| Yes | 10,411 (52.8) | 5248 (53.3) | 15,659 (52.9) |
| No | 9318 (47.2) | 4605 (46.7) | 13,923 (47.1) |
| **HIV-positive, n (%)** | 148 (0.8) | 61 (0.6) | 209 (0.7) |
| **Baseline serostatus, n (%)** | | | |
| Seronegative and RT-PCR negative | 18,489 (93.7) | 9178 (93.1) | 27,667 (93.5) |
| Seropositive or RT-PCR positive | 1240 (6.3) | 675 (6.9) | 1915 (6.5) |

Abbreviations: BMI = body mass index; max = maximum; min = minimum; NP = nucleocapsid; NVX-CoV2373 = 5 µg SARS-CoV-2 rS with 50 µg Matrix-M™ adjuvant; RT-PCR = reverse transcription-polymerase chain reaction; PPE = personal protective equipment; SARS-CoV-2 rS = severe acute respiratory syndrome coronavirus 2 recombinant spike protein nanoparticle vaccine; SD = standard deviation.

* Overall high-risk adults were defined as 1) age ≥65 years with or without comorbidities and/or living or working conditions involving known frequent exposure to SARS-CoV-2 or to densely populated circumstances; 2) age <65 years with comorbidities and/or living or working conditions involving known frequent exposure to SARS-CoV-2 or to densely populated circumstances.

† High risk for development of severe Covid-19 include participants 1) age ≥65 years with or without comorbidities and/or 2) age <65 years with comorbidities.[7]

#### Table S5. Vaccine Efficacy Against RT-PCR-Confirmed Symptomatic Mild, Moderate, or Severe Covid-19 at Least 7 Days After Second Vaccination in Adult Participants Not Previously Exposed to SARS-CoV-2 (PP-EFF Analysis Set)

| Parameter | NVX-CoV2373<br>N = 17,312 | Placebo<br>N = 8140 |
|---|---|---|
| Participants with no occurrence of event,* n (%) | 17,298 (99.9) | 8077 (99.2) |
| Participants with occurrence of event,[†] n (%) | 14 (0.1) | 63 (0.8) |
| Severity of first occurrence, n (%) | | |
| Mild | 14 (0.1) | 49 (0.6) |
| Moderate | 0 | 10 (0.1) |
| Severe | 0 | 4 (<0.1) |
| Median surveillance time[‡] (days) | 64.0 | 58.0 |
| Log-linear model using modified Poisson regression[§] | | |
| Mean disease incidence rate per year in 1000 people | 3.26 | 34.01 |
| 95% CI | 1.55, 6.89 | 20.70, 55.87 |
| Relative risk | 0.10 | |
| 95% CI | 0.05, 0.17 | |
| Vaccine efficacy (%) | 90.40 | |
| 95% CI | 82.88, 94.62 | |
| P-value[¶] | <0.001 | |
| Cox proportional hazard model (sensitivity analysis)[‖] | | |
| Vaccine efficacy (%) | 90.44 | |
| 95% CI | 82.94, 94.64 | |
| P-value[#] | <0.001 | |

Abbreviations: CI = confidence interval; Covid-19 = coronavirus disease 2019; NVX-CoV2373 = 5 µg SARS-CoV-2 rS with 50 µg Matrix-M™ adjuvant; RT-PCR = reverse transcriptase-polymerase chain reaction; PP-EFF = Per-Protocol Efficacy; SARS-CoV-2 rS = severe acute respiratory syndrome coronavirus 2 recombinant spike protein nanoparticle vaccine; VE = vaccine efficacy.

* Includes participants with RT-PCR-confirmed infection who did not meet mild, moderate, or severe Covid-19 criteria.

† Event = first occurrence of RT-PCR-confirmed mild, moderate, or severe Covid-19 with onset of illness episode from at least 7 days after second vaccination within the surveillance period.

‡ Surveillance time was defined as the difference between the date at end of surveillance period (onset of first occurrence of event/ censoring) and date at start of surveillance period (7 days after the second injection) + 1.

§ Modified Poisson regression with logarithmic link function, treatment group, and age strata as fixed effects and robust error variance.[6]

¶ This P-value corresponded to a one-sided hypothesis test with significance level 0.025. If the VE P-value <0.025, then reject H0: VE ≤30%.

‖ Cox-proportional hazard model with Efron's method for tie handling with vaccine group and age strata. Hazard ratio was used to estimate relative risk.

\# This P-value corresponded to a one-sided hypothesis test with significance level 0.025. If the VE P-value <0.025, then reject H0: VE ≤0%.

#### Table S6. Vaccine Efficacy Against RT-PCR-Confirmed Symptomatic Mild, Moderate, or Severe Covid-19 Due to a SARS-CoV-2 Variant Not Considered as a Variant of Concern or Variant of Interest at Least 7 Days After Second Vaccination in Adult Participants Not Previously Exposed to SARS-CoV-2 (PP-EFF Analysis Set)

| Parameter | NVX-CoV2373<br>N = 17,312 | Placebo<br>N = 8140 |
|---|---|---|
| Participants with no occurrence of event,* n (%) | 17,312 (100.0) | 8127 (99.8) |
| Participants with occurrence of event,[†] n (%) | 0 (0.0) | 13 (0.2) |
| Severity of first occurrence, n (%) | | |
| Mild | 0 (0.0) | 10 (0.1) |
| Moderate | 0 (0.0) | 2 (<0.1) |
| Severe | 0 (0.0) | 1 (<0.1) |
| Median surveillance time[‡] (days) | 64.0 | 58.0 |
| Exact conditional binomial method substituted for log-linear model using modified Poisson regression[§] | | |
| Mean disease incidence rate per year in 1000 people | 0.00 | 10.18 |
| 95% CI | <0.01, 1.25 | 5.42, 17.41 |
| Relative risk | 0.00 | |
| 95% CI | <0.01, 0.14 | |
| Vaccine efficacy (%) | 100.00 | |
| 95% CI | 85.84, 100.00 | |
| P-value[⁋] | <0.001 | |

Abbreviations: CDC = Centers for Disease Control and Prevention; CI = confidence interval; Covid-19 = coronavirus disease 2019; NVX-CoV2373 = 5 µg SARS-CoV-2 rS with 50 µg Matrix-M™ adjuvant; RT-PCR = reverse transcriptase-polymerase chain reaction; PP-EFF = Per-Protocol Efficacy; SARS-CoV-2 = severe acute respiratory syndrome coronavirus 2; SARS-CoV-2 rS = severe acute respiratory syndrome coronavirus 2 recombinant spike protein nanoparticle vaccine; SIG = SARS-CoV-2 Interagency Group; VE = vaccine efficacy; VOC = variant of concern; VOI = variant of interest.

* Includes participants with RT-PCR-confirmed infection who did not meet mild, moderate, or severe Covid-19 criteria and not considered a VOC or VOI.

† Event = first occurrence of RT-PCR-confirmed mild, moderate, or severe Covid-19 due to a SARS-CoV-2 variant not considered as a VOC or VOI with onset of illness episode from at least 7 days after second vaccination within the surveillance period.

‡ Surveillance time was defined as the difference between the date at end of surveillance period (onset of first occurrence of event/censoring) and date at start of surveillance period (7 days after the second injection) + 1.

§ In the event when there were zero cases in either group or the total number of cases in both treatment groups combined <5, VE and 95% CI were estimated with 1 – ratio of incidence rates using the exact method conditional on the total number of cases.

⁋ This P-value corresponded to a one-sided hypothesis test with significance level 0.025. If the VE P-value <0.025, then reject $H_0$: VE ≤30%.

Note: VOC/VOI were established by SIG and CDC for SARS-CoV-2 Variant Classifications and Definitions.[3]

#### Table S7. Vaccine Efficacy Against RT-PCR-Confirmed Symptomatic Moderate or Severe Covid-19 at Least 7 Days After Second Vaccination in Adult Participants Not Previously Exposed to SARS-CoV-2 (PP-EFF Analysis Set)

| Parameter | NVX-CoV2373<br>N = 17,312 | Placebo<br>N = 8140 |
|---|---|---|
| Participants with no occurrence of event,* n (%) | 17,312 (100.0) | 8126 (99.8) |
| Participants with occurrence of event,[†] n (%) | 0 (0.0) | 14 (0.2) |
| Severity of first occurrence, n (%) | | |
| Moderate | 0 (0.0) | 10 (0.1) |
| Severe | 0 (0.0) | 4 (<0.1) |
| Median surveillance time[‡] (days) | 64.0 | 58.0 |
| Exact conditional binomial method substituted for log-linear model using modified Poisson regression[§] | | |
| Mean disease incidence rate per year in 1000 people | 0.00 | 10.96 |
| 95% CI | 0.00, 1.25 | 5.99, 18.40 |
| Relative risk | 0.00 | |
| 95% CI | 0.00, 0.13 | |
| Vaccine efficacy (%) | 100.00 | |
| 95% CI | 86.99, 100.00 | |
| P-value[¶] | <0.001 | |

Abbreviations: CI = confidence interval; COVID-19 = coronavirus disease 2019; NVX-CoV2373 = 5 µg SARS-CoV-2 rS with 50 µg Matrix-M™ adjuvant; RT-PCR = reverse transcriptase-polymerase chain reaction; PP-EFF = Per-Protocol Efficacy; SARS-CoV-2 rS = severe acute respiratory syndrome coronavirus 2 recombinant spike protein nanoparticle vaccine; VE = vaccine efficacy.

* Includes participants with RT-PCR-confirmed infection who did not meet moderate or severe Covid-19 criteria.

† Event = first occurrence of RT-PCR-confirmed moderate or severe Covid-19 with onset of illness episode from at least 7 days after second vaccination within the surveillance period.

‡ Surveillance time was defined as the difference between the date at end of surveillance period (onset of first occurrence of event/censoring) and date at start of surveillance period (7 days after the second injection) + 1.

§ In the event when there were zero cases in either group or the total number of cases in both treatment groups combined <5, VE and 95% CI were estimated with 1 – ratio of incidence rates using the exact method conditional on the total number of cases. NE = not estimable in the event the test for exact binomial proportion could not be conducted.

¶ This P-value corresponds to a one-sided hypothesis test with significance level 0.025. If the VE P-value <0.025, then reject $H_0$: VE ≤0%.

**Table S8. Vaccine Efficacy Against RT-PCR-Confirmed Symptomatic Mild, Moderate or Severe Covid-19 Due to a SARS-CoV-2 Variant Considered as a Variant of Concern or Variant of Interest at Least 7 Days After Second Vaccination in Adult Participants Not Previously Exposed to SARS-CoV-2 (PP-EFF Analysis Set)**

| Parameter | NVX-CoV2373<br>N = 17,312 | Placebo<br>N = 8140 |
|---|---|---|
| Participants with no occurrence of event,* n (%) | 17,305 (100.0) | 8099 (99.5) |
| Participants with occurrence of event,† n (%) | 7 (<0.1) | 41 (0.5) |
| Severity of first occurrence, n (%) | | |
| Mild | 7 (<0.1) | 31 (0.4) |
| Moderate | 0 (0.0) | 8 (0.1) |
| Severe | 0 (0.0) | 2 (<0.1) |
| Median surveillance time‡ (days) | 64.0 | 58.0 |
| Log-linear model using modified Poisson regression§ | | |
| Mean disease incidence rate per year in 1000 people | 1.47 | 19.93 |
| 95% CI | 0.50, 4.30 | 9.95, 39.94 |
| Relative risk | 0.07 | |
| 95% CI | 0.03, 0.16 | |
| Vaccine efficacy (%) | 92.62 | |
| 95% CI | 83.56, 96.69 | |
| P-value⁋ | <0.001 | |

Abbreviations: CI = confidence interval; Covid-19 = coronavirus disease 2019; NVX-CoV2373 = 5 µg SARS-CoV-2 rS with 50 µg Matrix-M™ adjuvant; RT-PCR = reverse transcriptase-polymerase chain reaction; PP-EFF = Per-Protocol Efficacy; SARS-CoV-2 rS = severe acute respiratory syndrome coronavirus 2 recombinant spike protein nanoparticle vaccine; VE = vaccine efficacy; VOC = variant of concern; VOI = variant of interest.

* Includes participants with RT-PCR-confirmed infection who did not meet mild, moderate, or severe Covid-19 criteria and not considered a VOC or VOI.

† Event = first occurrence of RT-PCR-confirmed mild, moderate, or severe Covid-19 due to a VOC or VOI with onset of illness episode from at least 7 days after second vaccination within the surveillance period.

‡ Surveillance time was defined as the difference between the date at end of surveillance period (onset of first occurrence of event/censoring) and date at start of surveillance period (7 days after the second injection) + 1.

§ Modified Poisson regression with logarithmic link function, treatment group, and age strata as fixed effects and robust error variance.[6]

⁋ This P-value corresponded to a one-sided hypothesis test with significance level 0.025. If the VE P-value <0.025, then reject $H_0$: VE ≤0%.

Note: VOC/VOI were established by SIG and CDC for SARS-CoV-2 Variant Classifications and Definitions.[3]

#### Figure S1. SARS-CoV-2 Clade/Variant Identified in Per-Protocol Efficacy Covid-19 End Point Cases by Disease Severity

| | NVX-CoV2373 (n=17,312) | | Placebo (n=8,140) | |
|---|---|---|---|---|
| Total | 14 | | 63 | |
| | Variant | N (%) | Variant | N (%) |
| Mild Disease | *Alpha (B.1.1.7)*<br>*Beta (B.1.351)*<br>*Iota (B.1.526)*<br>*No sequence available* | *4*<br>*1*<br>*2*<br>*7* | *Alpha (B.1.1.7)*<br>*Beta (B.1.351)*<br>*Gamma (P.1)*<br>*Epsilon (B.1.429)*<br>*Iota (B.1.526)*<br>*Kappa (B.1.617.1)*<br>*Zeta (P.2)*<br>*B.1*<br>*B.1.1*<br>*B.1.1.316*<br>*B.1.1.519*<br>*B.1.2*<br>*B.1.243*<br>*B.1.311*<br>*B.1.596*<br>*No sequence available* | *20*<br>*1*<br>*2*<br>*1*<br>*5*<br>*1*<br>*1*<br>*1*<br>*1*<br>*1*<br>*1*<br>*1*<br>*2*<br>*2*<br>*1*<br>*8* |
| Moderate Disease | n/a | 0 | *Alpha (B.1.1.7)*<br>*Epsilon (B.1.429)*<br>*B.1.2* | *6*<br>*2*<br>*2* |
| Severe Disease | n/a | 0 | *Alpha (B.1.1.7)*<br>*Iota (B.1.526)*<br>*B.1.2*<br>*No sequence available* | *1*<br>*1*<br>*1*<br>*1* |

Abbreviations: Covid-19 = coronavirus disease 2019; NVX-CoV2373 = 5 µg SARS-CoV-2 rS with 50 µg Matrix-M™ adjuvant; SARS-CoV-2 = severe acute respiratory syndrome coronavirus 2, SARS-CoV-2 rS = severe acute respiratory syndrome coronavirus 2 recombinant spike protein nanoparticle vaccine.

Red: VOC, variant of concern, Orange: VOI, variant of interest, Green: not a VOC/VOI, as per CDC Classification.[3]

**Table S9. Overall Summary of Treatment-Emergent Adverse Events Reported From After Start of First Vaccination to Blinded Crossover or Early Withdrawal (Safety Analysis Set)**

| TEAE Category | NVX-CoV2373<br>N = 19,729 | | Placebo<br>N = 9853 | |
|---|---|---|---|---|
| | n (%) | E | n (%) | E |
| Any TEAE | 3216 (16.3) | 5647 | 1456 (14.8) | 2491 |
| Any severe TEAE* | 250 (1.3) | 359 | 108 (1.1) | 165 |
| Any treatment-related TEAE * | 798 (4.0) | 1422 | 239 (2.4) | 365 |
| Any severe treatment-related TEAE* | 59 (0.3) | 81 | 11 (0.1) | 19 |
| Any MAAE | 1387 (7.0) | 1995 | 651 (6.6) | 938 |
| Any treatment-related MAAE* | 92 (0.5) | 129 | 30 (0.3) | 36 |
| Any serious treatment-related MAAE* | 5 (< 0.1) | 8 | 0 | 0 |
| Any serious TEAE | 169 (0.9) | 228 | 94 (1.0) | 128 |
| Any TEAE leading to vaccination discontinuation | 57 (0.3) | 102 | 16 (0.2) | 26 |
| Any treatment-related TEAE leading to vaccination discontinuation* | 10 (0.1) | 17 | 3 (<0.1) | 4 |
| Any TEAE leading to study discontinuation | 60 (0.3) | 65 | 13 (0.1) | 14 |
| Any treatment-related TEAE leading to study discontinuation* | 14 (0.1) | 17 | 2 (<0.1) | 2 |
| Any AESI: PIMMC | 16 (0.1) | 21 | 3 (<0.1) | 4 |
| Any treatment-related AESI: PIMMC* | 10 (0.1) | 15 | 1 (<0.1) | 1 |
| Any AESI: relevant to Covid-19 | 4 (<0.1) | 6 | 4 (<0.1) | 7 |
| Any treatment-related AESI: relevant to Covid-19* | 0 | 0 | 0 | 0 |

Abbreviations: AESI = adverse event of special interest; Covid-19 = coronavirus disease 2019; E = number of events at each level of summarization; MAAE = medically attended adverse event; NVX-CoV2373 = 5 µg SARS-CoV-2 rS with 50 µg Matrix-M™ adjuvant; PIMMC = potential immune-mediated medical conditions; SARS-CoV-2 = severe acute respiratory syndrome coronavirus 2; SARS-CoV-2 rS = severe acute respiratory syndrome coronavirus 2 recombinant spike protein nanoparticle vaccine; TEAE = treatment-emergent adverse event.

* Relationship and severity were based on the data reported by site, i.e., missing information was not imputed.

#### Table S10. Summary of Solicited Local Adverse Events Within 7 Days After Dose 1 and Dose 2 in All Participants (Safety Analysis Set)

| Solicited Local Adverse Events | All Participants | |
|---|---|---|
| | NVX-CoV2373<br>N = 19,729/19,104 (%) | Placebo<br>N = 9853/9422 (%) |
| **Any local adverse event, N1/N2** | **18,072/17,139** | **8904/8278** |
| Dose 1 (Grade ≥1) | 10,475 (57.96) | 1881 (21.13) |
| *Grade 3* | 197 (1.09) | 22 (0.25) |
| *Grade 4* | 1 (< 0.01) | 1 (0.01) |
| Dose 2 (Grade ≥1) | 13,525 (78.91) | 1797 (21.71) |
| *Grade 3* | 1140 (6.65) | 25 (0.30) |
| *Grade 4* | 7 (0.04) | 1 (0.01) |
| **Any pain, N1/N2** | **18,072/17,139** | **8904/8278** |
| Dose 1 (Grade ≥1) | 6211 (34.37) | 986 (11.07) |
| *Grade 3* | 55 (0.30) | 3 (0.03) |
| *Grade 4* | 0 | 0 |
| Dose 2 (Grade ≥1) | 10,227 (59.67) | 1141 (13.78) |
| *Grade 3* | 297 (1.73) | 7 (0.08) |
| *Grade 4* | 5 (0.03) | 1 (0.01) |
| **Any tenderness, N1/N2** | **18,072/17,139** | **8904/8278** |
| Dose 1 (Grade ≥1) | 9450 (52.29) | 1494 (16.78) |
| *Grade 3* | 156 (0.86) | 18 (0.20) |
| *Grade 4* | 1 (<0.01) | 1 (0.01) |
| Dose 2 (Grade ≥1) | 12,584 (73.42) | 1312 (15.85) |
| *Grade 3* | 834 (4.87) | 18 (0.22) |
| *Grade 4* | 3 (0.02) | 0 |
| **Any erythema, N1/N2** | **18,072/17,139** | **8904/8278** |
| Dose 1 (Grade ≥1) | 164 (0.91) | 27 (0.30) |
| *Grade 3* | 3 (0.02) | 0 |
| *Grade 4* | 0 | 0 |
| Dose 2 (Grade ≥1) | 1138 (6.64) | 29 (035) |
| *Grade 3* | 143 (0.83) | 2 (0.02) |
| *Grade 4* | 0 | 0 |
| **Any swelling, N1/N2** | **18,072/17,139** | **8904/8278** |
| Dose 1 (Grade ≥1) | 154 (0.85) | 24 (0.27) |
| *Grade 3* | 7 (0.04) | 3 (0.03) |
| *Grade 4* | 0 | 0 |
| Dose 2 (Grade ≥1) | 1056 (6.16) | 25 (0.30) |
| *Grade 3* | 91 (0.53) | 2 (0.02) |
| *Grade 4* | 0 | 0 |

Abbreviations: FDA = US Food and Drug Administration; N = number of participants in the Safety Analysis Set following Dose 1/Dose 2; N1 = number of participants in the Safety Analysis Set who received the first dose and completed at least 1 day of the reactogenicity diary; N2 = number of participants in the Safety Analysis Set who received the second dose and completed at least 1 day of the reactogenicity diary; NVX-CoV2373 = 5 µg SARS-CoV-2 rS with 50 µg Matrix-M™ adjuvant; SARS-CoV-2 rS = severe acute respiratory syndrome coronavirus 2 recombinant spike protein nanoparticle vaccine.

Note: Data are presented as number (%) of participants experiencing a solicited event. Percentages were based on n/N1 × 100 and n/N2 × 100. At each level of participant summarization, a participant was counted once if they indicated the event.

Note: Grading of solicited adverse events was based on FDA Toxicity Grading Scale for Clinical Abnormalities.[8]

**Table S11. Duration (Days) of Solicited Local Adverse Events Within 7 Days After Dose 1 and Dose 2 in All Participants (Safety Analysis Set)**

| Solicited Local Adverse Events | NVX-CoV2373<br>N = 19,729/19,104 | Placebo<br>N = 9853/9422 |
|---|---|---|
| **Pain (# of days ≥ grade 1), N1/N2** | **18,072/17,139** | **8904/8278** |
| Dose 1, n | 6210 | 986 |
| Median | 1.0 | 1.0 |
| Minimum - maximum | 1 - 7 | 1 - 7 |
| Dose 2, n | 10226 | 1141 |
| Median | 2.0 | 1.0 |
| Minimum - maximum | 1 - 7 | 1 - 7 |
| **Tenderness (# of days ≥ grade 1), N1/N2** | **18,072/17,139** | **8904/8278** |
| Dose 1, n | 9450 | 1494 |
| Median | 2.0 | 1.0 |
| Minimum - maximum | 1 - 7 | 1 - 7 |
| Dose 2, n | 12584 | 1312 |
| Median | 2.0 | 1.0 |
| Minimum - maximum | 1 - 7 | 1 - 7 |
| **Erythema (# of days ≥ grade 1), N1/N2** | **18,072/17,139** | **8904/8278** |
| Dose 1, n | 164 | 27 |
| Median | 1.0 | 1.0 |
| Minimum - maximum | 1 - 7 | 1 - 5 |
| Dose 2, n | 1138 | 29 |
| Median | 2.0 | 1.0 |
| Minimum - maximum | 1 - 7 | 1 - 6 |
| **Swelling (# of days ≥ grade 1), N1/N2** | **18,072/17,139** | **8904/8278** |
| Dose 1, n | 154 | 24 |
| Median | 1.0 | 1.0 |
| Minimum - maximum | 1 - 7 | 1 - 5 |
| Dose 2, n | 1056 | 25 |
| Median | 2.0 | 1.0 |
| Minimum - maximum | 1 - 7 | 1 - 5 |

Abbreviations: n = number of participants who reported the solicited event; N = number of participants in the Safety Analysis Set following Dose 1/Dose 2; N1 = number of participants in the Safety Analysis Set who received the first dose and completed at least 1 day of the reactogenicity diary; N2 = number of participants in the Safety Analysis Set who received the second dose and completed at least 1 day of the reactogenicity diary; NVX-CoV2373 = 5 µg SARS-CoV-2 rS with 50 µg Matrix-M™ adjuvant; SARS-CoV-2 rS = severe acute respiratory syndrome coronavirus 2 recombinant spike protein nanoparticle vaccine.

Note: Duration is calculated as the number of days the solicited event was greater than grade 0. n = number of subjects who reported the event.

**Table S12. Summary of Solicited Systemic Adverse Events Within 7 Days After Dose 1 and Dose 2 by Age Group (Safety Analysis Set)**

| Solicited Systemic Adverse Events | All Participants | |
|---|---|---|
| | **NVX-CoV2373 N = 19,729/19,104 (%)** | **Placebo N = 9853/9422 (%)** |
| **Any solicited systemic TEAE, N1/N2** | **18,072/17,139** | **8904/8278** |
| Dose 1 (Grade ≥1) | 8614 (47.66) | 3562 (40.00) |
| *Grade 3* | 422 (2.34) | 183 (2.06) |
| *Grade 4* | 17 (0.09) | 5 (0.06) |
| Dose 2 (Grade ≥1) | 11,906 (69.47) | 2969 (35.87) |
| *Grade 3* | 2056 (12.00) | 165 (1.99) |
| *Grade 4* | 21 (0.12) | 5 (0.06) |
| **Headache, N1/N2** | **18,072/17,139** | **8904/8278** |
| Dose 1 (Grade ≥1) | 4505 (24.93) | 2028 (22.78) |
| *Grade 3* | 146 (0.81) | 62 (0.70) |
| *Grade 4* | 5 (0.03) | 1 (0.01) |
| Dose 2 (Grade ≥1) | 7618 (44.45) | 1625 (19.63) |
| *Grade 3* | 512 (2.99) | 36 (0.43) |
| *Grade 4* | 6 (0.04) | 2 (0.02) |
| **Fatigue, N1/N2** | **18,072/17,139** | **8904/8278** |
| Dose 1 (Grade ≥1) | 4632 (25.63) | 1993 (22.38) |
| *Grade 3* | 224 (1.24) | 100 (1.12) |
| *Grade 4* | 3 (0.02) | 1 (0.01) |
| Dose 2 (Grade ≥1) | 8486 (49.51) | 1811 (21.88) |
| *Grade 3* | 1419 (8.28) | 108 (1.30) |
| *Grade 4* | 4 (0.02) | 3 (0.04) |
| **Malaise, N1/N2** | **18,072/17,139** | **8904/8278** |
| Dose 1 (Grade ≥1) | 2660 (14.72) | 1037 (11.65) |
| *Grade 3* | 137 (0.76) | 53 (0.60) |
| *Grade 4* | 7 (0.04) | 2 (0.02) |
| Dose 2 (Grade ≥1) | 6674 (38.94) | 1018 (12.30) |
| *Grade 3* | 1073 (6.26) | 57 (0.69) |
| *Grade 4* | 9 (0.05) | 2 (0.02) |
| **Muscle pain, N1/N2** | **18,072/17,139** | **8904/8278** |
| Dose 1 (Grade ≥1) | 4102 (22.70) | 1188 (13.34) |
| *Grade 3* | 81 (0.45) | 35 (0.39) |
| *Grade 4* | 2 (0.01) | 2 (0.02) |
| Dose 2 (Grade ≥1) | 8240 (48.08) | 1001 (12.09) |
| *Grade 3* | 841 (4.91) | 29 (0.35) |
| *Grade 4* | 5 (0.03) | 4 (0.05) |
| **Joint pain, N1/N2** | **18,072/17,139** | **8904/8278** |
| Dose 1 (Grade ≥1) | 1388 (7.68) | 590 (6.63) |

| Solicited Systemic Adverse Events | All Participants | |
|---|---|---|
| | **NVX-CoV2373**<br>**N = 19,729/19,104 (%)** | **Placebo**<br>**N = 9853/9422 (%)** |
| *Grade 3* | 51 (0.28) | 29 (0.33) |
| *Grade 4* | 1 (< 0.01) | 0 |
| Dose 2 (Grade ≥1) | 3809 (22.22) | 567 (6.85) |
| *Grade 3* | 411 (2.40) | 24 (0.29) |
| *Grade 4* | 6 (0.04) | 2 (0.02) |
| **Fever, N1/N2** | **18,072/17,139** | **8904/8278** |
| Dose 1 (Grade ≥1) | 66 (0.37) | 33 (0.37) |
| *Grade 3* | 8 (0.04) | 6 (0.07) |
| *Grade 4* | 6 (0.03) | 1 (0.01) |
| Dose 2 (Grade ≥1) | 973 (5.68) | 23 (0.28) |
| *Grade 3* | 62 (0.36) | 3 (0.04) |
| *Grade 4* | 2 (0.01) | 0 |
| **Nausea/Vomiting, N1/N2** | **18,072/17,139** | **8904/8278** |
| Dose 1 (Grade ≥1) | 1152 (6.37) | 488 (5.48) |
| *Grade 3* | 17 (0.09) | 7 (0.08) |
| *Grade 4* | 4 (0.02) | 3 (0.03) |
| Dose 2 (Grade ≥1) | 1929 (11.26) | 450 (5.44) |
| *Grade 3* | 29 (0.17) | 7 (0.08) |
| *Grade 4* | 7 (0.04) | 2 (0.02) |

Abbreviations: FDA = US Food and Drug Administration; N = number of participants in the Safety Analysis Set following Dose 1/Dose 2; N1 = number of participants in the Safety Analysis Set who received the first dose and completed at least 1 day of the reactogenicity diary; N2 = number of participants in the Safety Analysis Set who received the second dose and completed at least 1 day of the reactogenicity diary; NVX-CoV2373 = 5 µg SARS-CoV-2 rS with 50 µg Matrix-M™ adjuvant; SARS-CoV-2 rS = severe acute respiratory syndrome coronavirus 2 recombinant spike protein nanoparticle vaccine.

Note: Data are presented as number (%) of participants experiencing a solicited event. Percentages were based on n/N1 × 100 and n/N2 × 100. At each level of participant summarization, a participant was counted once if they indicated the event occurred and provided a severity during the reactogenicity period. The highest severity experienced during the reactogenicity period is summarized in this table.

Note: Grading of solicited adverse events was based on FDA Toxicity Grading Scale for Clinical Abnormalities.[8]

**Table S13. Duration (Days) of Solicited Systemic Adverse Events Within 7 Days After Dose 1 and Dose 2 in All Participants (Safety Analysis Set)**

| Solicited Systemic Adverse Events | NVX-CoV2373<br>N = 19,729/19,104 | Placebo<br>N = 9853/9422 |
|---|---|---|
| **Headache (# of days ≥ grade 1), N1/N2** | **18,072/17,139** | **8904/8278** |
| Dose 1, n | 4505 | 2028 |
| Median | 1.0 | 1.0 |
| Minimum - maximum | 1 - 7 | 1 - 7 |
| Dose 2, n | 7618 | 1625 |
| Median | 1.0 | 1.0 |
| Minimum - maximum | 1 - 7 | 1 - 7 |
| **Fatigue (# of days ≥ grade 1), N1/N2** | **18,072/17,139** | **8904/8278** |
| Dose 1, n | 4632 | 1993 |
| Median | 1.0 | 1.0 |
| Minimum - maximum | 1 - 7 | 1 - 7 |
| Dose 2, n | 8486 | 1811 |
| Median | 1.0 | 1.0 |
| Minimum - maximum | 1 - 7 | 1 - 7 |
| **Malaise (# of days ≥ grade 1), N1/N2** | **18,072/17,139** | **8904/8278** |
| Dose 1, n | 2660 | 1037 |
| Median | 1.0 | 1.0 |
| Minimum - maximum | 1 - 7 | 1 - 7 |
| Dose 2, n | 6674 | 1018 |
| Median | 1.0 | 1.0 |
| Minimum - maximum | 1 - 7 | 1 - 7 |
| **Muscle pain (# of days ≥ grade 1), N1/N2** | **18,072/17,139** | **8904/8278** |
| Dose 1, n | 4102 | 1188 |
| Median | 1.0 | 1.0 |
| Minimum - maximum | 1 - 7 | 1 - 7 |
| Dose 2, n | 8240 | 1000 |
| Median | 1.0 | 1.0 |
| Minimum - maximum | 1 - 7 | 1 - 7 |
| **Joint pain (# of days ≥ grade 1), N1/N2** | **18,072/17,139** | **8904/8278** |
| Dose 1, n | 1388 | 590 |
| Median | 1.0 | 1.0 |
| Minimum - maximum | 1 - 7 | 1 - 7 |
| Dose 2, n | 3809 | 567 |
| Median | 1.0 | 1.0 |
| Minimum - maximum | 1 - 7 | 1 - 7 |
| **Fever (# of days ≥ grade 1), N1/N2** | **18,072/17,139** | **8904/8278** |
| Dose 1, n | 66 | 33 |
| Median | 1.0 | 1.0 |
| Minimum - maximum | 1 - 5 | 1 - 2 |
| Dose 2, n | 973 | 23 |
| Median | 1.0 | 1.0 |

| Solicited Systemic Adverse Events | NVX-CoV2373<br>N = 19,729/19,104 | Placebo<br>N = 9853/9422 |
|---|---|---|
| Minimum - maximum | 1 - 6 | 1 - 4 |
| **Nausea/Vomiting (# of days ≥ grade 1), N1/N2** | **18,072/17,139** | **8904/8278** |
| Dose 1, n | 1152 | 488 |
| Median | 1.0 | 1.0 |
| Minimum - maximum | 1 - 7 | 1 - 6 |
| Dose 2, n | 1929 | 450 |
| Median | 1.0 | 1.0 |
| Minimum - maximum | 1 - 7 | 1 - 7 |

Abbreviations: n = number of participants who reported the solicited event; N = number of participants in the Safety Analysis Set following Dose 1/Dose 2; N1 = number of participants in the Safety Analysis Set who received the first dose and completed at least 1 day of the reactogenicity diary; N2 = number of participants in the Safety Analysis Set who received the second dose and completed at least 1 day of the reactogenicity diary; NVX-CoV2373 = 5 µg SARS-CoV-2 rS with 50 µg Matrix-M™ adjuvant; SARS-CoV-2 rS = severe acute respiratory syndrome coronavirus 2 recombinant spike protein nanoparticle vaccine.

Note: Duration is calculated as the number of days the solicited event was greater than grade 0. n=number of subjects who reported the event.

**Table S14. Summary of Unsolicited Adverse Events by System Organ Class and Preferred Term Reported From After Start of First Vaccination Through 28 Days After Second Vaccination (e.g., Day 49) for Events With an Incidence Rate ≥1 Event per 100 Person-Years by Age Strata (Safety Analysis Set)**

| System Organ Class/Preferred Term (MedDRA, Version 23.1) | Participants ≥18 Years | | | | Participants 18 to ≤64 Years | | | | Participants ≥65 Years | | | |
|---|---|---|---|---|---|---|---|---|---|---|---|---|
| | NVX-CoV2373 N = 19,729 | | Placebo N = 9853 | | NVX-CoV2373 N = 17,251 | | Placebo N = 8616 | | NVX-CoV2373 N = 2478 | | Placebo N = 1237 | |
| | E | IR | E | IR | E | IR | E | IR | E | IR | E | IR |
| **Number of participants experiencing an event** | **4299** | **156.37** | **1794** | **131.98** | **3781** | **156.87** | **1590** | **133.15** | **518** | **152.84** | **204** | **123.56** |
| **General disorders and administration site conditions** | **925** | **33.65** | **228** | **16.77** | **831** | **34.48** | **203** | **17.00** | **94** | **27.73** | **25** | **15.14** |
| Fatigue | 196 | 7.13 | 69 | 5.08 | 173 | 7.18 | 63 | 5.28 | 23 | 6.79 | 6 | 3.63 |
| Injection site pain | 160 | 5.82 | 38 | 2.80 | 139 | 5.77 | 31 | 2.60 | 21 | 6.20 | 7 | 4.24 |
| Pyrexia | 116 | 4.22 | 23 | 1.69 | 109 | 4.52 | 22 | 1.84 | 7 | 2.07 | 1 | 0.61 |
| Pain | 68 | 2.47 | 21 | 1.54 | 61 | 2.53 | 21 | 1.76 | 7 | 2.07 | 0 | 0 |
| Chills | 65 | 2.36 | 6 | 0.44 | 61 | 2.53 | 6 | 0.50 | 4 | 1.18 | 0 | 0 |
| Malaise | 48 | 1.75 | 14 | 1.03 | 44 | 1.83 | 12 | 1.00 | 4 | 1.18 | 2 | 1.21 |
| Injection site pruritus | 44 | 1.60 | 2 | 0.15 | 39 | 1.62 | 2 | 0.17 | 5 | 1.48 | 0 | 0 |
| Injection site erythema | 29 | 1.05 | 4 | 0.29 | 28 | 1.16 | 4 | 0.33 | 1 | 0.30 | 0 | 0 |
| Edema peripheral | 15 | 0.55 | 4 | 0.29 | 10 | 0.41 | 2 | 0.17 | 5 | 1.48 | 2 | 1.21 |
| **Nervous system disorders** | **514** | **18.70** | **246** | **18.10** | **461** | **19.13** | **223** | **18.67** | **53** | **15.64** | **23** | **13.93** |
| Headache | 293 | 10.66 | 130 | 9.56 | 271 | 11.24 | 118 | 9.88 | 22 | 6.49 | 12 | 7.27 |
| Dizziness | 45 | 1.64 | 23 | 1.69 | 37 | 1.54 | 20 | 1.67 | 8 | 2.36 | 3 | 1.82 |
| Tension headache | 27 | 0.98 | 15 | 1.10 | 21 | 0.87 | 13 | 1.09 | 6 | 1.77 | 2 | 1.21 |
| **Infections and infestations** | **507** | **18.44** | **284** | **20.89** | **445** | **18.46** | **250** | **20.94** | **62** | **18.29** | **34** | **20.59** |
| Upper respiratory tract infection | 63 | 2.29 | 31 | 2.28 | 55 | 2.28 | 27 | 2.26 | 8 | 2.36 | 4 | 2.42 |
| Urinary tract infection | 52 | 1.89 | 19 | 1.40 | 43 | 1.78 | 14 | 1.17 | 9 | 2.66 | 5 | 3.03 |
| Covid-19 | 49 | 1.78 | 38 | 2.80 | 42 | 1.74 | 36 | 3.01 | 7 | 2.07 | 2 | 1.21 |
| Viral infection | 31 | 1.13 | 17 | 1.25 | 27 | 1.12 | 16 | 1.34 | 4 | 1.18 | 1 | 0.61 |
| Sinusitis | 29 | 1.05 | 18 | 1.32 | 23 | 0.95 | 17 | 1.42 | 6 | 1.77 | 1 | 0.61 |
| Cellulitis | 15 | 0.55 | 8 | 0.59 | 11 | 0.46 | 5 | 0.42 | 4 | 1.18 | 3 | 1.82 |
| Tooth infection | 4 | 0.15 | 15 | 1.10 | 4 | 0.17 | 13 | 1.09 | 0 | 0 | 2 | 1.21 |
| **Respiratory, thoracic, and mediastinal disorders** | **415** | **15.09** | **210** | **15.45** | **364** | **15.10** | **189** | **15.83** | **51** | **15.05** | **21** | **12.72** |

| System Organ Class/Preferred Term (MedDRA, Version 23.1) | Participants ≥18 Years | | | | Participants 18 to ≤64 Years | | | | Participants ≥65 Years | | | |
|---|---|---|---|---|---|---|---|---|---|---|---|---|
| | NVX-CoV2373 N = 19,729 | | Placebo N = 9853 | | NVX-CoV2373 N = 17,251 | | Placebo N = 8616 | | NVX-CoV2373 N = 2478 | | Placebo N = 1237 | |
| | **E** | **IR** | **E** | **IR** | **E** | **IR** | **E** | **IR** | **E** | **IR** | **E** | **IR** |
| Nasal congestion | 111 | 4.04 | 67 | 4.93 | 97 | 4.02 | 62 | 5.19 | 14 | 4.13 | 5 | 3.03 |
| Cough | 78 | 2.84 | 41 | 3.02 | 68 | 2.82 | 38 | 3.18 | 10 | 2.95 | 3 | 1.82 |
| Rhinorrhea | 55 | 2.00 | 20 | 1.47 | 45 | 1.87 | 17 | 1.42 | 10 | 2.95 | 3 | 1.82 |
| Oropharyngeal pain | 48 | 1.75 | 27 | 1.99 | 42 | 1.74 | 27 | 2.26 | 6 | 1.77 | 0 | 0 |
| Dyspnea | 32 | 1.16 | 18 | 1.32 | 31 | 1.29 | 14 | 1.17 | 1 | 0.30 | 4 | 2.42 |
| **Musculoskeletal and connective tissue disorders** | **378** | **13.75** | **161** | **11.84** | **326** | **13.53** | **132** | **11.05** | **52** | **15.34** | **29** | **17.56** |
| Myalgia | 109 | 3.96 | 30 | 2.21 | 93 | 3.86 | 24 | 2.01 | 16 | 4.72 | 6 | 3.63 |
| Arthralgia | 64 | 2.33 | 36 | 2.65 | 56 | 2.32 | 32 | 2.68 | 8 | 2.36 | 4 | 2.42 |
| Pain in extremity | 51 | 1.86 | 17 | 1.25 | 47 | 1.95 | 15 | 1.26 | 4 | 1.18 | 2 | 1.21 |
| Back pain | 38 | 1.38 | 21 | 1.54 | 33 | 1.37 | 17 | 1.42 | 5 | 1.48 | 4 | 2.42 |
| Osteoarthritis | 11 | 0.40 | 5 | 0.37 | 8 | 0.33 | 2 | 0.17 | 3 | 0.89 | 3 | 1.82 |
| **Gastrointestinal disorders** | **376** | **13.68** | **189** | **13.90** | **335** | **13.90** | **174** | **14.57** | **41** | **12.10** | **15** | **9.09** |
| Diarrhea | 89 | 3.24 | 60 | 4.41 | 73 | 3.03 | 57 | 4.77 | 16 | 4.72 | 3 | 1.82 |
| Nausea | 86 | 3.13 | 39 | 2.87 | 80 | 3.32 | 34 | 2.85 | 6 | 1.77 | 5 | 3.03 |
| Vomiting | 35 | 1.27 | 12 | 0.88 | 34 | 1.41 | 11 | 0.92 | 1 | 0.30 | 1 | 0.61 |
| Gastroesophageal reflux disease | 15 | 0.55 | 6 | 0.44 | 14 | 0.58 | 4 | 0.33 | 1 | 0.30 | 2 | 1.21 |
| **Skin and subcutaneous tissue disorders** | **218** | **7.93** | **71** | **5.22** | **197** | **8.17** | **67** | **5.61** | **21** | **6.20** | **4** | **2.42** |
| Rash | 61 | 2.22 | 22 | 1.62 | 55 | 2.28 | 21 | 1.76 | 6 | 1.77 | 1 | 0.61 |
| **Injury, poisoning, and procedural complications** | **205** | **7.46** | **102** | **7.50** | **173** | **7.18** | **87** | **7.29** | **32** | **9.44** | **15** | **9.09** |
| Ligament sprain | 11 | 0.40 | 8 | 0.59 | 9 | 0.37 | 6 | 0.50 | 2 | 0.59 | 2 | 1.21 |
| Fall | 10 | 0.36 | 7 | 0.51 | 6 | 0.25 | 4 | 0.33 | 4 | 1.18 | 3 | 1.82 |
| **Psychiatric disorders** | **109** | **3.96** | **48** | **3.53** | **103** | **4.27** | **47** | **3.94** | **6** | **1.77** | **1** | **0.61** |
| Anxiety | 32 | 1.16 | 14 | 1.03 | 31 | 1.29 | 13 | 1.09 | 1 | 0.30 | 1 | 0.61 |
| **Vascular disorders** | **108** | **3.93** | **50** | **3.68** | **82** | **3.40** | **43** | **3.60** | **26** | **7.67** | **7** | **4.24** |
| Hypertension | 71 | 2.58 | 40 | 2.94 | 54 | 2.24 | 36 | 3.01 | 17 | 5.02 | 4 | 2.42 |
| **Metabolism and nutrition disorders** | **79** | **2.87** | **47** | **3.46** | **62** | **2.57** | **44** | **3.68** | **17** | **5.02** | **3** | **1.82** |
| **Blood and lymphatic disorders** | **78** | **2.84** | **22** | **1.62** | **76** | **3.15** | **21** | **1.76** | **2** | **0.59** | **1** | **0.61** |
| Lymphadenopathy | 53 | 1.93 | 13 | 0.96 | 52 | 2.16 | 12 | 1.00 | 1 | 0.30 | 1 | 0.61 |
| **Investigations** | **67** | **2.44** | **28** | **2.06** | **54** | **2.24** | **23** | **1.93** | **13** | **3.84** | **5** | **3.03** |

| System Organ Class/Preferred Term (MedDRA, Version 23.1) | Participants ≥18 Years | | | | Participants 18 to ≤64 Years | | | | Participants ≥65 Years | | | |
|---|---|---|---|---|---|---|---|---|---|---|---|---|
| | NVX-CoV2373 N = 19,729 | | Placebo N = 9853 | | NVX-CoV2373 N = 17,251 | | Placebo N = 8616 | | NVX-CoV2373 N = 2478 | | Placebo N = 1237 | |
| | E | IR | E | IR | E | IR | E | IR | E | IR | E | IR |
| Blood pressure increased | 15 | 0.55 | 5 | 0.37 | 12 | 0.50 | 3 | 0.25 | 3 | 0.89 | 2 | 1.21 |
| SARS-CoV-2 test positive | 12 | 0.44 | 12 | 0.88 | 10 | 0.41 | 10 | 0.84 | 2 | 0.59 | 2 | 1.21 |
| **Eye disorders** | **62** | **2.26** | **13** | **0.96** | **54** | **2.24** | **10** | **0.84** | **8** | **2.36** | **3** | **1.82** |
| **Reproductive system and breast disorders** | **55** | **2.00** | **17** | **1.25** | **51** | **2.12** | **15** | **1.26** | **4** | **1.18** | **2** | **1.21** |
| **Cardiac disorders** | **45** | **1.64** | **21** | **1.54** | **34** | **1.41** | **14** | **1.17** | **11** | **3.25** | **7** | **4.24** |
| **Ear and labyrinth disorders** | **44** | **1.60** | **16** | **1.18** | **39** | **1.62** | **13** | **1.09** | **5** | **1.48** | **3** | **1.82** |
| **Immune system disorders** | **29** | **1.05** | **7** | **0.51** | **27** | **1.12** | **7** | **0.59** | **2** | **0.59** | **0** | **0** |
| **Neoplasms benign, malignant and unspecified (including cysts and polyps)** | **26** | **0.95** | **7** | **0.51** | **19** | **0.79** | **4** | **0.33** | **7** | **2.07** | **3** | **1.82** |
| **Renal and urinary disorders** | **26** | **0.95** | **10** | **0.74** | **17** | **0.71** | **9** | **0.75** | **9** | **2.66** | **1** | **0.61** |

Abbreviations: E = number of AEs reported; IR = incidence rate is defined as number of events per 100 person-years = e/100 PY; MedDRA = Medical Dictionary for Regulatory Activities; NVX-CoV2373 = 5 µg SARS-CoV-2 rS with 50 µg Matrix-M™ adjuvant; SARS-CoV-2 = severe acute respiratory syndrome coronavirus 2; SARS-CoV-2 rS = severe acute respiratory syndrome coronavirus 2 recombinant spike protein nanoparticle vaccine.

Note: Events and IRs at the system organ class level represent all events.

**Table S15. Summary of Unsolicited Serious Adverse Events by System Organ Class and Preferred Term From Start of First Vaccination to Blinded Crossover or Early Termination by Age Strata (Safety Analysis Set)**

| System Organ Class/ Preferred Term (MedDRA, Version 23.1) | Participants ≥18 Years | | | | Participants 18 to ≤64 Years | | | | Participants ≥65 Years | | | |
|---|---|---|---|---|---|---|---|---|---|---|---|---|
| | NVX-CoV2373 N = 19,729 | | Placebo N = 9853 | | NVX-CoV2373 N = 17,251 | | Placebo N = 8616 | | NVX-CoV2373 N = 2478 | | Placebo N = 1237 | |
| | **E** | **IR** | **E** | **IR** | **E** | **IR** | **E** | **IR** | **E** | **IR** | **E** | **IR** |
| **Number of participants experiencing an event** | **228** | **4.32** | **128** | **4.89** | **169** | **3.68** | **103** | **4.52** | **59** | **8.48** | **25** | **7.41** |
| **Infections and infestations** | **33** | **0.62** | **35** | **1.34** | **26** | **0.57** | **28** | **1.23** | **7** | **1.01** | **7** | **2.08** |
| Appendicitis | 5 | 0.09 | 4 | 0.15 | 4 | 0.09 | 4 | 0.18 | 1 | 0.14 | 0 | 0 |
| Covid-19 | 5 | 0.09 | 9 | 0.34 | 3 | 0.07 | 8 | 0.35 | 2 | 0.29 | 1 | 0.30 |
| Pneumonia | 4 | 0.08 | 2 | 0.08 | 2 | 0.04 | 1 | 0.04 | 2 | 0.29 | 1 | 0.30 |
| Cellulitis | 2 | 0.04 | 2 | 0.08 | 1 | 0.02 | 1 | 0.04 | 1 | 0.14 | 1 | 0.30 |
| Sepsis | 2 | 0.04 | 2 | 0.08 | 1 | 0.02 | 1 | 0.04 | 1 | 0.14 | 1 | 0.30 |
| Abscess limb | 1 | 0.02 | 1 | 0.04 | 1 | 0.02 | 1 | 0.04 | 0 | 0 | 0 | 0 |
| Appendicitis perforated | 1 | 0.02 | 1 | 0.04 | 1 | 0.02 | 1 | 0.04 | 0 | 0 | 0 | 0 |
| Arthritis bacterial | 1 | 0.02 | 1 | 0.04 | 1 | 0.02 | 0 | 0 | 0 | 0 | 1 | 0.30 |
| Empyema | 1 | 0.02 | 0 | 0 | 1 | 0.02 | 0 | 0 | 0 | 0 | 0 | 0 |
| Localized infection | 1 | 0.02 | 0 | 0 | 1 | 0.02 | 0 | 0 | 0 | 0 | 0 | 0 |
| Mastitis | 1 | 0.02 | 0 | 0 | 1 | 0.02 | 0 | 0 | 0 | 0 | 0 | 0 |
| Necrotizing soft tissue infection | 1 | 0.02 | 0 | 0 | 1 | 0.02 | 0 | 0 | 0 | 0 | 0 | 0 |
| Osteomyelitis | 1 | 0.02 | 0 | 0 | 1 | 0.02 | 0 | 0 | 0 | 0 | 0 | 0 |
| Perineal abscess | 1 | 0.02 | 0 | 0 | 1 | 0.02 | 0 | 0 | 0 | 0 | 0 | 0 |
| Post-procedural infection | 1 | 0.02 | 0 | 0 | 1 | 0.02 | 0 | 0 | 0 | 0 | 0 | 0 |
| Pyelonephritis | 1 | 0.02 | 0 | 0 | 1 | 0.02 | 0 | 0 | 0 | 0 | 0 | 0 |
| Septic shock | 1 | 0.02 | 1 | 0.04 | 1 | 0.02 | 1 | 0.04 | 0 | 0 | 0 | 0 |
| Subcutaneous abscess | 1 | 0.02 | 0 | 0 | 1 | 0.02 | 0 | 0 | 0 | 0 | 0 | 0 |
| Urosepsis | 1 | 0.02 | 0 | 0 | 1 | 0.02 | 0 | 0 | 0 | 0 | 0 | 0 |
| Appendicitis perforated | 1 | 0.02 | 1 | 0.04 | 1 | 0.02 | 1 | 0.04 | 0 | 0 | 0 | 0 |
| Arthritis bacterial | 1 | 0.02 | 1 | 0.04 | 1 | 0.02 | 0 | 0 | 0 | 0 | 1 | 0.30 |
| Abdominal wall abscess | 0 | 0 | 1 | 0.04 | 0 | 0 | 1 | 0.04 | 0 | 0 | 0 | 0 |
| Covid-19 pneumonia | 0 | 0 | 6 | 0.23 | 0 | 0 | 5 | 0.22 | 0 | 0 | 1 | 0.30 |
| Diverticulitis | 0 | 0 | 2 | 0.08 | 0 | 0 | 2 | 0.09 | 0 | 0 | 0 | 0 |
| Groin abscess | 0 | 0 | 1 | 0.04 | 0 | 0 | 1 | 0.04 | 0 | 0 | 0 | 0 |
| Pneumonia fungal | 0 | 0 | 1 | 0.04 | 0 | 0 | 1 | 0.04 | 0 | 0 | 0 | 0 |
| Streptococcal bacteremia | 0 | 0 | 1 | 0.04 | 0 | 0 | 0 | 0 | 0 | 0 | 1 | 0.30 |
| **Cardiac disorders** | **30** | **0.57** | **14** | **0.54** | **16** | **0.35** | **10** | **0.44** | **14** | **2.01** | **4** | **1.19** |
| Atrial fibrillation | 7 | 0.13 | 2 | 0.08 | 4 | 0.09 | 1 | 0.04 | 3 | 0.43 | 1 | 0.30 |
| Myocardial infarction | 4 | 0.08 | 2 | 0.08 | 2 | 0.04 | 1 | 0.04 | 2 | 0.29 | 1 | 0.30 |

| System Organ Class/ Preferred Term (MedDRA, Version 23.1) | Participants ≥18 Years | | | | Participants 18 to ≤64 Years | | | | Participants ≥65 Years | | | |
|---|---|---|---|---|---|---|---|---|---|---|---|---|
| | NVX-CoV2373 N = 19,729 | | Placebo N = 9853 | | NVX-CoV2373 N = 17,251 | | Placebo N = 8616 | | NVX-CoV2373 N = 2478 | | Placebo N = 1237 | |
| | **E** | **IR** | **E** | **IR** | **E** | **IR** | **E** | **IR** | **E** | **IR** | **E** | **IR** |
| Acute left ventricular failure | 3 | 0.06 | 0 | 0 | 1 | 0.02 | 0 | 0 | 2 | 0.29 | 0 | 0 |
| Cardiac arrest | 3 | 0.06 | 3 | 0.11 | 2 | 0.04 | 3 | 0.13 | 1 | 0.14 | 0 | 0 |
| Cardiac failure congestive | 3 | 0.06 | 1 | 0.04 | 1 | 0.02 | 1 | 0.04 | 2 | 0.29 | 0 | 0 |
| Acute myocardial infarction | 2 | 0.04 | 2 | 0.08 | 2 | 0.04 | 1 | 0.04 | 0 | 0 | 1 | 0.30 |
| Angina pectoris | 1 | 0.02 | 0 | 0 | 4 | 0.09 | 0 | 0 | **0** | **0** | **0** | **0** |
| Bradycardia | 1 | 0.02 | 0 | 0 | 0 | 0 | **0** | **0** | 1 | 0.14 | 0 | 0 |
| Cardiac pseudoaneurysm | 1 | 0.02 | 0 | 0 | 1 | 0.02 | 0 | 0 | **0** | **0** | **0** | **0** |
| Coronary artery disease | 1 | 0.02 | 2 | 0.08 | 2 | 0.04 | 1 | 0.04 | 0 | 0 | 1 | 0.30 |
| Myocardial ischemia | 1 | 0.02 | 0 | 0 | 0 | 0 | 1 | 0.04 | 1 | 0.14 | 0 | 0 |
| Myocarditis | 1 | 0.02 | 1 | 0.04 | 0 | 0 | 0 | 0 | 1 | 0.14 | 0 | 0 |
| Palpitations | 1 | 0.02 | 0 | 0 | 1 | 0.02 | **0** | **0** | 0 | 0 | 0 | 0 |
| Ventricular tachycardia | 1 | 0.02 | 0 | 0 | 0 | 0 | 0 | 0 | 1 | 0.14 | 0 | 0 |
| Cardio-respiratory arrest | 0 | 0 | 1 | 0.04 | 0 | 0.00 | 1 | 0.04 | **0** | **0** | **0** | **0** |
| **Injury, poisoning, and procedural complications** | **25** | **0.47** | **14** | **0.54** | **18** | **0.39** | **13** | **0.57** | **7** | **1.01** | **1** | **0.30** |
| Alcohol poisoning | 2 | 0.04 | 1 | 0.04 | 2 | 0.04 | 1 | 0.04 | 0 | 0 | 0 | 0 |
| Femur fracture | 2 | 0.04 | 0 | 0 | 0 | 0 | 0 | 0 | 2 | 0.29 | 0 | 0 |
| Rib fracture | 2 | 0.04 | 1 | 0.04 | 1 | 0.02 | 1 | 0.04 | 1 | 0.14 | 0 | 0 |
| Accidental overdose | 1 | 0.02 | 0 | 0 | 1 | 0.02 | 0 | 0 | 0 | 0 | 0 | 0 |
| Ankle fracture | 1 | 0.02 | 0 | 0 | 1 | 0.02 | 0 | 0 | 0 | 0 | 0 | 0 |
| Burns third degree | 1 | 0.02 | 0 | 0 | 1 | 0.02 | 0 | 0 | 0 | 0 | 0 | 0 |
| Concussion | 1 | 0.02 | 0 | 0 | 1 | 0.02 | 0 | 0 | 0 | 0 | 0 | 0 |
| Exposure to toxic agent | 1 | 0.02 | 0 | 0 | 0 | 0 | 0 | 0 | 1 | 0.04 | 0 | 0 |
| Fall | 1 | 0.02 | 2 | 0.08 | 1 | 0.02 | 1 | 0.04 | 0 | 0 | 1 | 0.30 |
| Fibula fracture | 1 | 0.02 | 1 | 0.04 | 1 | 0.02 | 1 | 0.04 | 0 | 0 | 0 | 0 |
| Gunshot wound | 1 | 0.02 | 0 | 0 | 1 | 0.02 | 0 | 0 | 0 | 0 | 0 | 0 |
| Hip fracture | 1 | 0.02 | 0 | 0 | 0 | 0 | 0 | 0 | 1 | 0.14 | 0 | 0 |
| Incisional hernia | 1 | 0.02 | 0 | 0 | 1 | 0.02 | 0 | 0 | 0 | 0 | 0 | 0 |
| Jaw fracture | 1 | 0.02 | 0 | 0 | 0 | 0 | 0 | 0 | 1 | 0.14 | 0 | 0 |
| Overdose | 1 | 0.02 | 2 | 0.08 | 1 | 0.02 | 2 | 0.09 | 0 | 0 | 0 | 0 |
| Radius fracture | 1 | 0.02 | 0 | 0 | 1 | 0.02 | 0 | 0 | **0** | **0** | **0** | **0** |
| Snake bite | 1 | 0.02 | 0 | 0 | 1 | 0.02 | 0 | 0 | 0 | 0 | 0 | 0 |
| Spinal fracture | 1 | 0.02 | 0 | 0 | 1 | 0.02 | 0 | 0 | 0 | 0 | 0 | 0 |

| System Organ Class/ Preferred Term (MedDRA, Version 23.1) | Participants ≥18 Years | | | | Participants 18 to ≤64 Years | | | | Participants ≥65 Years | | | |
|---|---|---|---|---|---|---|---|---|---|---|---|---|
| | NVX-CoV2373 N = 19,729 | | Placebo N = 9853 | | NVX-CoV2373 N = 17,251 | | Placebo N = 8616 | | NVX-CoV2373 N = 2478 | | Placebo N = 1237 | |
| | E | IR | E | IR | E | IR | E | IR | E | IR | E | IR |
| Splenic rupture | 1 | 0.02 | 0 | 0 | 1 | 0.02 | 0 | 0 | 0 | 0 | 0 | 0 |
| Tibia fracture | 1 | 0.02 | 1 | 0.04 | 1 | 0.02 | 1 | 0.04 | 0 | 0 | 0 | 0 |
| Traumatic hematoma | 1 | 0.02 | 0 | 0 | 1 | 0.02 | 0 | 0 | 0 | 0 | 0 | 0 |
| Wrist fracture | 1 | 0.02 | 0 | 0 | 0 | 0 | 0 | 0 | 1 | 0.14 | 0 | 0 |
| Foot fracture | 0 | 0 | 1 | 0.04 | 0 | 0 | 1 | 0.04 | 0 | 0 | 0 | 0 |
| Foreign body in gastrointestinal tract | 0 | 0 | 1 | 0.04 | 0 | 0 | 1 | 0.04 | 0 | 0 | 0 | 0 |
| Injury | 0 | 0 | 1 | 0.04 | 0 | 0 | 1 | 0.04 | 0 | 0 | 0 | 0 |
| Joint injury | 0 | 0 | 1 | 0.04 | 0 | 0 | 1 | 0.04 | 0 | 0 | 0 | 0 |
| Lumbar vertebral fracture | 0 | 0 | 1 | 0.04 | 0 | 0 | 1 | 0.04 | 0 | 0 | 0 | 0 |
| Road traffic accident | 0 | 0 | 1 | 0.04 | 0 | 0 | 1 | 0.04 | 0 | 0 | 0 | 0 |
| **Nervous system disorders** | **19** | **0.36** | **9** | **0.34** | **16** | **0.35** | **8** | **0.35** | **3** | **0.43** | **1** | **0.30** |
| Cerebrovascular accident | 7 | 0.13 | 1 | 0.04 | 5 | 0.11 | 0 | 0 | 2 | 0.29 | 1 | 0.30 |
| Ischemic stroke | 2 | 0.04 | 0 | 0 | 1 | 0.02 | 0 | 0 | 1 | 0.14 | 0 | 0 |
| Seizure | 2 | 0.04 | 1 | 0.04 | 2 | 0.04 | 1 | 0.04 | 0 | 0 | 0 | 0 |
| Alcoholic seizure | 1 | 0.02 | 0 | 0 | 1 | 0.02 | 8 | 0.35 | 0 | 0 | 0 | 0 |
| Altered state of consciousness | 1 | 0.02 | 0 | 0 | 1 | 0.02 | 0 | 0 | 0 | 0 | 0 | 0 |
| Central nervous system inflammation | 1 | 0.02 | 0 | 0 | 1 | 0.02 | 1 | 0.04 | 0 | 0 | 0 | 0 |
| Cervicogenic headache | 1 | 0.02 | 0 | 0 | 1 | 0.02 | 0 | 0 | 0 | 0 | 0 | 0 |
| Neuropathy peripheral | 1 | 0.02 | 0 | 0 | 1 | 0.02 | 0 | 0 | 0 | 0 | 0 | 0 |
| Peroneal nerve palsy | 1 | 0.02 | 0 | 0 | 1 | 0.02 | 0 | 0 | 0 | 0 | 0 | 0 |
| Presyncope | 1 | 0.02 | 0 | 0 | 1 | 0.02 | 0 | 0 | 0 | 0 | 0 | 0 |
| Transient ischemic attack | 1 | 0.02 | 1 | 0.04 | 1 | 0.02 | 1 | 0.04 | 0 | 0 | 0 | 0 |
| Carotid artery stenosis | 0 | 0 | 1 | 0.04 | 0 | 0 | 0 | 0 | 0 | 0 | 0 | 0 |
| Cerebellar infarction | 0 | 0 | 1 | 0.04 | 0 | 0 | 0 | 0 | 0 | 0 | 0 | 0 |
| Generalized tonic-clonic seizure | 0 | 0 | 1 | 0.04 | 0 | 0 | 1 | 0.04 | 0 | 0 | 0 | 0 |
| Hypoesthesia | 0 | 0 | 1 | 0.04 | 0 | 0 | 1 | 0.04 | 0 | 0 | 0 | 0 |
| Syncope | 0 | 0 | 2 | 0.08 | 0 | 0 | 2 | 0.09 | 0 | 0 | 0 | 0 |
| **Gastrointestinal disorders** | **16** | **0.30** | **6** | **0.23** | **14** | **0.31** | **4** | **0.18** | **2** | **0.29** | **2** | **0.59** |
| Intestinal obstruction | 2 | 0.04 | 0 | 0 | 2 | 0.04 | 0 | 0 | 0 | 0 | 0 | 0 |
| Abdominal pain | 1 | 0.02 | 0 | 0 | 1 | 0.02 | 0 | 0 | 0 | 0 | 0 | 0 |
| Alcoholic pancreatitis | 1 | 0.02 | 0 | 0 | 1 | 0.02 | 0 | 0 | 0 | 0 | 0 | 0 |
| Ascites | 1 | 0.02 | 0 | 0 | 1 | 0.02 | 0 | 0 | 0 | 0 | 0 | 0 |

| System Organ Class/ Preferred Term (MedDRA, Version 23.1) | Participants ≥18 Years | | | | Participants 18 to ≤64 Years | | | | Participants ≥65 Years | | | |
|---|---|---|---|---|---|---|---|---|---|---|---|---|
| | NVX-CoV2373 N = 19,729 | | Placebo N = 9853 | | NVX-CoV2373 N = 17,251 | | Placebo N = 8616 | | NVX-CoV2373 N = 2478 | | Placebo N = 1237 | |
| | **E** | **IR** | **E** | **IR** | **E** | **IR** | **E** | **IR** | **E** | **IR** | **E** | **IR** |
| Colitis ulcerative | 1 | 0.02 | 0 | 0 | 0 | 0 | 0 | 0 | 1 | 0.14 | 0 | 0 |
| Gastritis | 1 | 0.02 | 0 | 0 | 1 | 0.02 | 0 | 0 | 0 | 0 | 0 | 0 |
| Gastrointestinal hemorrhage | 1 | 0.02 | 0 | 0 | 0 | 0 | 0 | 0 | 1 | 0.14 | 0 | 0 |
| Hematemesis | 1 | 0.02 | 1 | 0.04 | 1 | 0.02 | 0 | 0 | 0 | 0 | 0 | 0 |
| Hiatus hernia | 1 | 0.02 | 0 | 0 | 1 | 0.02 | 0 | 0 | 0 | 0 | 0 | 0 |
| Impaired gastric emptying | 1 | 0.02 | 1 | 0.04 | 1 | 0.02 | 1 | 0.04 | 0 | 0 | 0 | 0 |
| Mallory-Weiss syndrome | 1 | 0.02 | 0 | 0 | 1 | 0.02 | 0 | 0 | 0 | 0 | 0 | 0 |
| Pancreatitis | 1 | 0.02 | 0 | 0 | 1 | 0.02 | 0 | 0 | 0 | 0 | 0 | 0 |
| Pancreatitis acute | 1 | 0.02 | 0 | 0 | 1 | 0.02 | 0 | 0 | 0 | 0 | 0 | 0 |
| Peptic ulcer | 1 | 0.02 | 0 | 0 | 1 | 0.02 | 0 | 0 | 0 | 0 | 0 | 0 |
| Rectal hemorrhage | 1 | 0.02 | 0 | 0 | 1 | 0.02 | 0 | 0 | 0 | 0 | 0 | 0 |
| Duodenal ulcer | 0 | 0 | 1 | 0.04 | 0 | 0 | 0 | 0 | 0 | 0 | 0 | 0 |
| Gastric hemorrhage | 0 | 0 | 1 | 0.04 | 0 | 0 | 1 | 0.04 | 0 | 0 | 0 | 0 |
| Nausea | 0 | 0 | 1 | 0.04 | 0 | 0 | 0 | 0 | 0 | 0 | 1 | 0.30 |
| Vomiting | 0 | 0 | 1 | 0.04 | 0 | 0 | 0 | 0 | 0 | 0 | 1 | 0.30 |
| **Respiratory, thoracic, and mediastinal disorders** | **16** | **0.30** | **8** | **0.31** | **11** | **0.24** | **5** | **0.22** | **5** | **0.72** | **3** | **0.89** |
| Acute respiratory failure | 3 | 0.06 | 0 | 0 | 1 | 0.02 | 0 | 0 | 2 | 0.29 | 0 | 0 |
| Pneumonia aspiration | 3 | 0.06 | 0 | 0 | 3 | 0.07 | 0 | 0 | 0 | 0 | 0 | 0 |
| Pulmonary embolism | 3 | 0.06 | 2 | 0.08 | 1 | 0.02 | 1 | 0.04 | 2 | 0.29 | 1 | 0.30 |
| Chronic obstructive pulmonary disease | 2 | 0.04 | 0 | 0 | 1 | 0.02 | 0 | 0 | 1 | 0.14 | 0 | 0 |
| Dyspnea | 2 | 0.04 | 2 | 0.08 | 2 | 0.04 | 1 | 0.04 | 0 | 0 | 1 | 0.30 |
| Asthma | 1 | 0.02 | 2 | 0.08 | 1 | 0.02 | 1 | 0.04 | 0 | 0 | 1 | 0.30 |
| Pulmonary hypertension | 1 | 0.02 | 0 | 0.00 | 1 | 0.02 | 0 | 0 | 0 | 0 | 0 | 0 |
| Respiratory failure | 1 | 0.02 | 1 | 0.04 | 1 | 0.02 | 1 | 0.04 | 0 | 0 | 0 | 0 |
| Pneumothorax | 0 | 0 | 1 | 0.04 | 0 | 0 | 1 | 0.04 | 0 | 0 | 0 | 0 |
| **Neoplasms benign, malignant, and unspecified (including cysts and polyps)** | **14** | **0.27** | **4** | **0.15** | **8** | **0.17** | **4** | **0.18** | **6** | **0.86** | **0** | **0** |
| Prostate cancer | 6 | 0.11 | 0 | 0 | 2 | 0.04 | 0 | 0 | 4 | 0.58 | 0 | 0 |
| Breast cancer | 2 | 0.04 | 0 | 0 | 2 | 0.04 | 0 | 0 | 0 | 0 | 0 | 0 |
| Breast cancer metastatic | 1 | 0.02 | 0 | 0 | 1 | 0.02 | 0 | 0 | 1 | 0.14 | 0 | 0 |
| Breast cancer stage III | 1 | 0.02 | 0 | 0 | 1 | 0.02 | 0 | 0 | 0 | 0 | 0 | 0 |
| Chronic myeloid leukemia | 1 | 0.02 | 0 | 0 | 0 | 0 | 0 | 0 | 0 | 0 | 0 | 0 |

| System Organ Class/ Preferred Term (MedDRA, Version 23.1) | Participants ≥18 Years | | | | Participants 18 to ≤64 Years | | | | Participants ≥65 Years | | | |
|---|---|---|---|---|---|---|---|---|---|---|---|---|
| | NVX-CoV2373 N = 19,729 | | Placebo N = 9853 | | NVX-CoV2373 N = 17,251 | | Placebo N = 8616 | | NVX-CoV2373 N = 2478 | | Placebo N = 1237 | |
| | **E** | **IR** | **E** | **IR** | **E** | **IR** | **E** | **IR** | **E** | **IR** | **E** | **IR** |
| Malignant melanoma | 1 | 0.02 | 0 | 0 | 8 | 0.17 | 0 | 0 | 0 | 0 | 0 | 0 |
| Non-Hodgkin's lymphoma | 1 | 0.02 | 0 | 0 | 0 | 0 | 0 | 0 | 1 | 0.14 | 0 | 0 |
| Testis cancer | 1 | 0.02 | 0 | 0 | 1 | 0.02 | 0 | 0 | 0 | 0 | 0 | 0 |
| Anal squamous cell carcinoma | 0 | 0 | 1 | 0.04 | 0 | 0 | 1 | 0.04 | 0 | 0 | 0 | 0 |
| Endometrial adenocarcinoma | 0 | 0 | 2 | 0.08 | 0 | 0 | 2 | 0.09 | 0 | 0 | 0 | 0 |
| Invasive ductal breast carcinoma | 0 | 0 | 1 | 0.04 | 1 | 0.02 | 1 | 0.04 | 0 | 0 | 0 | 0 |
| **Vascular disorders** | **12** | **0.23** | **4** | **0.15** | **8** | **0.17** | **4** | **0.18** | **4** | **0.58** | **0** | **0** |
| Hypertension | 3 | 0.06 | 0 | 0 | 3 | 0.07 | 0 | 0 | 0 | 0 | 0 | 0 |
| Deep vein thrombosis | 2 | 0.04 | 0 | 0 | 0 | 0 | 0 | 0 | 2 | 0.29 | 0 | 0 |
| Hypertensive crisis | 2 | 0.04 | 0 | 0 | 1 | 0.02 | 2 | 0.09 | 0 | 0 | 0 | 0 |
| Arterial occlusive disease | 1 | 0.02 | 0 | 0 | 0 | 0 | 0 | 0 | 1 | 0.14 | 0 | 0 |
| Circulatory collapse | 1 | 0.02 | 0 | 0 | 1 | 0.02 | 0 | 0 | 0 | 0 | 0 | 0 |
| Hematoma | 1 | 0.02 | 0 | 0 | 0 | 0 | 0 | 0 | 1 | 0.14 | 0 | 0 |
| Hypertension | 1 | 0.02 | 0 | 0 | 3 | 0.07 | 0 | 0 | 0 | 0 | 0 | 0 |
| Intermittent claudication | 1 | 0.02 | 0 | 0 | 1 | 0.02 | 0 | 0 | 0 | 0 | 0 | 0 |
| Embolism | 0 | 0 | 1 | 0.04 | 0 | 0 | 1 | 0.04 | 0 | 0 | 0 | 0 |
| Hypotension | 0 | 0 | 1 | 0.04 | 2 | 0.04 | 1 | 0.04 | 0 | 0 | 0 | 0 |
| **Hepatobiliary disorders** | **11** | **0.21** | **0** | **0** | **11** | **0.24** | **0** | **0** | **0** | **0** | **0** | **0** |
| Cholecystitis acute | 5 | 0.09 | 0 | 0 | 5 | 0.11 | 0 | 0 | 0 | 0 | 0 | 0 |
| Bile duct stone | 2 | 0.04 | 0 | 0 | 2 | 0.04 | 0 | 0 | 0 | 0 | 0 | 0 |
| Cholecystitis | 2 | 0.04 | 0 | 0 | 2 | 0.04 | 0 | 0 | 0 | 0 | 0 | 0 |
| Cholelithiasis | 1 | 0.02 | 0 | 0 | 1 | 0.02 | 0 | 0 | 0 | 0 | 0 | 0 |
| Cirrhosis alcoholic | 1 | 0.02 | 0 | 0 | 1 | 0.02 | 0 | 0 | 0 | 0 | 0 | 0 |
| **Psychiatric disorders** | **11** | **0.21** | **8** | **0.31** | **11** | **0.24** | **6** | **0.26** | **0** | **0** | **2** | **0.59** |
| Suicidal ideation | 3 | 0.06 | 3 | 0.11 | 3 | 0.07 | 2 | 0.09 | 0 | 0 | 1 | 0.30 |
| Bipolar disorder | 2 | 0.04 | 1 | 0.04 | 2 | 0.04 | 0 | 0 | 0 | 0 | 1 | 0.30 |
| Depression | 2 | 0.04 | 0 | 0 | 2 | 0.04 | 0 | 0 | 0 | 0 | 0 | 0 |
| Drug abuse | 1 | 0.02 | 1 | 0.04 | 2 | 0.04 | 1 | 0.04 | 0 | 0 | 0 | 0 |
| Homicidal ideation | 1 | 0.02 | 0 | 0.00 | 1 | 0.02 | 0 | 0.00 | 0 | 0 | 0 | 0 |
| Psychiatric symptom | 1 | 0.02 | 0 | 0.00 | 0 | 0.00 | 0 | 0.00 | 0 | 0 | 0 | 0 |
| Substance abuse | 1 | 0.02 | 0 | 0.00 | 1 | 0.02 | 0 | 0.00 | 0 | 0 | 0 | 0 |
| Alcohol abuse | 0 | 0 | 1 | 0.04 | 0 | 0 | 1 | 0.04 | 0 | 0 | 0 | 0 |

| System Organ Class/ Preferred Term (MedDRA, Version 23.1) | Participants ≥18 Years | | | | Participants 18 to ≤64 Years | | | | Participants ≥65 Years | | | |
|---|---|---|---|---|---|---|---|---|---|---|---|---|
| | NVX-CoV2373 N = 19,729 | | Placebo N = 9853 | | NVX-CoV2373 N = 17,251 | | Placebo N = 8616 | | NVX-CoV2373 N = 2478 | | Placebo N = 1237 | |
| | E | IR | E | IR | E | IR | E | IR | E | IR | E | IR |
| Delusion | 0 | 0 | 1 | 0.04 | 0 | 0 | 1 | 0.04 | 0 | 0 | 0 | 0 |
| Panic attack | 0 | 0 | 1 | 0.04 | 1 | 0.02 | 1 | 0.04 | 0 | 0 | 0 | 0 |
| **General disorders and administration site conditions** | **7** | **0.13** | **6** | **0.23** | **3** | **0.07** | **4** | **0.18** | **4** | **0.58** | **2** | **0.59** |
| Asthenia | 2 | 0.04 | 0 | 0 | 0 | 0 | 0 | 0 | 2 | 0.29 | 0 | 0 |
| Chest pain | 1 | 0.02 | 3 | 0.11 | 1 | 0.02 | 2 | 0.09 | 0 | 0 | 1 | 0.30 |
| Drug withdrawal syndrome | 1 | 0.02 | 0 | 0 | 1 | 0.02 | 0 | 0 | 0 | 0 | 0 | 0 |
| Mucosal inflammation | 1 | 0.02 | 0 | 0 | 0 | 0 | 0 | 0 | 1 | 0.14 | 0 | 0 |
| Edema peripheral | 1 | 0.02 | 1 | 0.04 | 0 | 0 | 1 | 0.04 | 1 | 0.14 | 0 | 0 |
| Sudden death | 1 | 0.02 | 0 | 0 | 1 | 0.02 | 0 | 0 | 0 | 0 | 0 | 0 |
| Catheter site thrombosis | 0 | 0 | 1 | 0.04 | 0 | 0 | 1 | 0.04 | 0 | 0 | 0 | 0 |
| Edema | 0 | 0 | 1 | 0.04 | 0 | 0 | 0 | 0 | 0 | 0 | 1 | 0.30 |
| **Musculoskeletal and connective tissue disorders** | **7** | **0.13** | **2** | **0.08** | **4** | **0.09** | **2** | **0.09** | **3** | **0.43** | **0** | **0** |
| Intervertebral disc protrusion | 2 | 0.04 | 0 | 0 | 1 | 0.02 | 0 | 0 | 1 | 0.14 | 0 | 0 |
| Arthralgia | 1 | 0.02 | 0 | 0 | 1 | 0.02 | 0 | 0 | 0 | 0 | 0 | 0 |
| Cervical spinal stenosis | 1 | 0.02 | 0 | 0 | 1 | 0.02 | 0 | 0 | **0** | **0** | 0 | 0 |
| Osteoarthritis | 1 | 0.02 | 0 | 0 | 0 | 0 | 2 | 0.09 | 1 | 0.14 | 0 | 0 |
| Osteolysis | 1 | 0.02 | 0 | 0 | 0 | 0 | 0 | 0 | 1 | 0.14 | 0 | 0 |
| Rhabdomyolysis | 1 | 0.02 | 0 | 0 | 1 | 0.02 | 0 | 0 | 0 | 0 | 0 | 0 |
| Intervertebral disc disorder | 0 | 0 | 1 | 0.04 | 0 | 0 | 1 | 0.04 | 1 | 0.14 | 0 | 0 |
| Neck pain | 0 | 0 | 1 | 0.04 | 0 | 0 | 1 | 0.04 | 0 | 0 | 0 | 0 |
| **Pregnancy, puerperium, and perinatal conditions** | **6** | **0.11** | **1** | **0.04** | **6** | **0.13** | **1** | **0.04** | **0** | **0** | **0** | **0** |
| Abortion spontaneous | 3 | 0.06 | 1 | 0.04 | 3 | 0.07 | 1 | 0.04 | 0 | 0 | 0 | 0 |
| Pregnancy | 2 | 0.04 | 0 | 0 | 2 | 0.04 | 0 | 0 | 0 | 0 | 0 | 0 |
| Abortion spontaneous complete | 1 | 0.02 | 0 | 0 | 1 | 0.02 | 0 | 0 | 0 | 0 | 0 | 0 |
| **Blood and lymphatic system disorders** | **5** | **0.09** | **1** | **0.04** | **4** | **0.09** | **1** | **0.04** | **1** | **0.14** | **0** | **0** |
| Blood loss anemia | 1 | 0.02 | 0 | 0 | 1 | 0.02 | 0 | 0 | 0 | 0 | 0 | 0 |
| Iron deficiency anemia | 1 | 0.02 | 0 | 0 | 1 | 0.02 | 0 | 0 | 0 | 0 | 0 | 0 |
| Neutropenia | 1 | 0.02 | 0 | 0 | 1 | 0.02 | 0 | 0 | 1 | 0.14 | 0 | 0 |
| Normocytic anemia | 1 | 0.02 | 0 | 0 | 1 | 0.02 | 0 | 0 | 0 | 0 | 0 | 0 |
| Thrombocytopenia | 1 | 0.02 | 0 | 0 | 1 | 0.02 | 0 | 0 | 0 | 0 | 0 | 0 |
| Leukocytosis | 0 | 0 | 1 | 0.04 | 0 | 0 | 1 | 0.04 | 0 | 0 | 0 | 0 |
| **Renal and urinary disorders** | **4** | **0.08** | **4** | **0.15** | **2** | **0.04** | **3** | **0.13** | **2** | **0.29** | **1** | **0.30** |

| System Organ Class/ Preferred Term (MedDRA, Version 23.1) | Participants ≥18 Years | | | | Participants 18 to ≤64 Years | | | | Participants ≥65 Years | | | |
|---|---|---|---|---|---|---|---|---|---|---|---|---|
| | NVX-CoV2373 N = 19,729 | | Placebo N = 9853 | | NVX-CoV2373 N = 17,251 | | Placebo N = 8616 | | NVX-CoV2373 N = 2478 | | Placebo N = 1237 | |
| | E | IR | E | IR | E | IR | E | IR | E | IR | E | IR |
| Acute kidney injury | 2 | 0.04 | 1 | 0.04 | 1 | 0.02 | 1 | 0.04 | 1 | 0.14 | 0 | 0 |
| Chronic kidney disease | 1 | 0.02 | 0 | 0 | 0 | 0 | 0 | 0 | 1 | 0.14 | 0 | 0 |
| Nephrolithiasis | 1 | 0.02 | 2 | 0.08 | 1 | 0.02 | 2 | 0.09 | 0 | 0 | 0 | 0 |
| Renal failure | 0 | 0 | 1 | 0.04 | 0 | 0 | 0 | 0 | 0 | 0 | 1 | 0.30 |
| **Endocrine disorders** | **3** | **0.06** | **0** | **0** | **3** | **0.07** | **0** | **0** | **0** | **0** | **0** | **0** |
| Basedow's disease | 1 | 0.02 | 0 | 0 | 1 | 0.02 | 0 | 0 | 0 | 0 | 0 | 0 |
| Hyperparathyroidism | 1 | 0.02 | 0 | 0 | 1 | 0.02 | 0 | 0 | 0 | 0 | 0 | 0 |
| Hyperthyroidism | 1 | 0.02 | 0 | 0 | 1 | 0.02 | 0 | 0 | 0 | 0 | 0 | 0 |
| **Metabolism and nutrition disorders** | **3** | **0.06** | **7** | **0.27** | **3** | **0.07** | **7** | **0.31** | **0** | **0** | **0** | **0** |
| Diabetic ketoacidosis | 1 | 0.02 | 0 | 0 | 1 | 0.02 | 0 | 0 | 0 | 0 | 0 | 0 |
| Electrolyte imbalance | 1 | 0.02 | 0 | 0 | 1 | 0.02 | 0 | 0 | 0 | 0 | 0 | 0 |
| Type 2 diabetes mellitus | 1 | 0.02 | 0 | 0 | 1 | 0.02 | 0 | 0 | 0 | 0 | 0 | 0 |
| Dehydration | 0 | 0 | 2 | 0.08 | 0 | 0 | 2 | 0.09 | 0 | 0 | 0 | 0 |
| Hyperglycemia | 0 | 0 | 1 | 0.04 | 0 | 0 | 1 | 0.04 | 0 | 0 | 0 | 0 |
| Hypoglycemia | 0 | 0 | 1 | 0.04 | 0 | 0 | 1 | 0.04 | 0 | 0 | 0 | 0 |
| Hypokalemia | 0 | 0 | 2 | 0.08 | 0 | 0 | 2 | 0.09 | 0 | 0 | 0 | 0 |
| Hyponatremia | 0 | 0 | 1 | 0.04 | 0 | 0 | 1 | 0.04 | 0 | 0 | 0 | 0 |
| **Skin and subcutaneous tissue disorders** | **2** | **0.04** | **0** | **0** | **1** | **0.02** | **0** | **0** | **1** | **0.14** | **0** | **0** |
| Angioedema | 1 | 0.02 | 0 | 0 | 1 | 0.02 | 0 | 0 | 1 | 0.14 | 0 | 0 |
| Dermatitis | 1 | 0.02 | 0 | 0 | 1 | 0.02 | 0 | 0 | 0 | 0 | 0 | 0 |
| **Uncoded** | **2** | **0.04** | **0** | **0** | **2** | **0.04** | **0** | **0** | **0** | **0** | **0** | **0** |
| Cholecystitis and cholelithiasis | 1 | 0.02 | 0 | 0 | 1 | 0.02 | 0 | 0 | 0 | 0 | 0 | 0 |
| Syncope leading to focal seizure disorder | 1 | 0.02 | 0 | 0 | 1 | 0.02 | 0 | 0 | 0 | 0 | 0 | 0 |
| **Eye disorders** | 1 | 0.02 | 0 | 0 | 1 | 0.02 | 0 | 0 | 0 | 0 | 0 | 0 |
| Diplopia | 1 | 0.02 | 0 | 0 | 1 | 0.02 | 0 | 0 | 0 | 0 | 0 | 0 |
| **Surgical and medical procedures** | **1** | **0.02** | **2** | **0.08** | **1** | **0.02** | **1** | **0.04** | **0** | **0** | **1** | **0.30** |
| Coronary arterial stent insertion | 1 | 0.02 | 0 | 0 | 1 | 0.02 | 0 | 0 | 0 | 0 | 0 | 0 |
| Hip arthroplasty | 0 | 0 | 1 | 0.04 | 0 | 0 | 1 | 0.04 | 0 | 0 | 0 | 0 |
| Spinal fusion surgery | 0 | 0 | 1 | 0.04 | 0 | 0 | 0 | 0 | 0 | 0 | 1 | 0.30 |
| **Investigations** | **0** | **0** | **2** | **0.08** | **0** | **0** | **2** | **0.09** | **0** | **0** | **0** | **0** |
| Oxygen saturation decreased | 0 | 0 | 1 | 0.04 | 0 | 0 | 1 | 0.04 | 0 | 0 | 0 | 0 |
| SARS-CoV-2 test positive | 0 | 0 | 1 | 0.04 | 0 | 0 | 1 | 0.04 | 0 | 0 | 0 | 0 |

| System Organ Class/ Preferred Term (MedDRA, Version 23.1) | Participants ≥18 Years | | | | Participants 18 to ≤64 Years | | | | Participants ≥65 Years | | | |
|---|---|---|---|---|---|---|---|---|---|---|---|---|
| | NVX-CoV2373 N = 19,729 | | Placebo N = 9853 | | NVX-CoV2373 N = 17,251 | | Placebo N = 8616 | | NVX-CoV2373 N = 2478 | | Placebo N = 1237 | |
| | E | IR | E | IR | E | IR | E | IR | E | IR | E | IR |
| **Reproductive system and breast disorders** | **0** | **0** | **1** | **0.04** | **0** | **0** | **0** | **0** | **0** | **0** | **1** | **0.30** |
| Benign prostatic hyperplasia | 0 | 0 | 1 | 0.04 | 0 | 0 | 0 | 0 | 0 | 0 | 1 | 0.30 |

Abbreviations: E = number of AEs reported; IR = incidence rate is defined as number of events per 100 person-years = e/100 PY; MedDRA = Medical Dictionary for Regulatory Activities; NVX-CoV2373 = 5 µg SARS-CoV-2 rS with 50 µg Matrix-M™ adjuvant; SARS-CoV-2 = severe acute respiratory syndrome coronavirus 2; SARS-CoV-2 rS = severe acute respiratory syndrome coronavirus 2 recombinant spike protein nanoparticle vaccine.

#### Table S16. Serious AEs of Interest Reported in PREVENT-19, Including Safety Signals Observed During Use of Other Covid-19 Vaccines (Safety Analysis Set)

| System Organ Class/ Preferred Term (MedDRA, Version 23.1) | Participants ≥18 Years | | | | Participants 18 to ≤64 Years | | | | Participants ≥65 Years | | | |
|---|---|---|---|---|---|---|---|---|---|---|---|---|
| | NVX-CoV2373 N = 19,729 | | Placebo N = 9853 | | NVX-CoV2373 N = 17,251 | | Placebo N = 8616 | | NVX-CoV2373 N = 2478 | | Placebo N = 1237 | |
| | E | IR | E | IR | E | IR | E | IR | E | IR | E | IR |
| **Cardiac disorders** | | | | | | | | | | | | |
| Myocardial infarction | 4 | 0.08 | 2 | 0.08 | 2 | 0.04 | 1 | 0.04 | 2 | 0.29 | 1 | 0.30 |
| Myocarditis | 1 | 0.02 | 1 | 0.04 | 0 | 0 | 0 | 0 | 1 | 0.14 | 0 | 0 |
| Pericarditis | 0 | 0 | 0 | 0 | 0 | 0 | 0 | 0 | 0 | 0 | 0 | 0 |
| **Respiratory, thoracic, and mediastinal disorders** | | | | | | | | | | | | |
| Pulmonary embolism | 3 | 0.06 | 2 | 0.08 | 1 | 0.02 | 1 | 0.04 | 2 | 0.29 | 1 | 0.30 |
| **Vascular disorders** | | | | | | | | | | | | |
| Deep vein thrombosis | 2 | 0.04 | 0 | 0 | 0 | 0 | 0 | 0 | 2 | 0.29 | 0 | 0 |
| Embolism | 0 | 0 | 1 | 0.04 | 0 | 0 | 1 | 0.04 | 0 | 0 | 0 | 0 |
| Cerebral venous sinus thrombosis | 0 | 0 | 0 | 0 | 0 | 0 | 0 | 0 | 0 | 0 | 0 | 0 |
| **Blood and lymphatic system disorders** | | | | | | | | | | | | |
| Thrombocytopenia | 1 | 0.02 | 0 | 0 | 1 | 0.02 | 0 | 0 | 0 | 0 | 0 | 0 |
| Disseminated intravascular coagulation | 0 | 0 | 0 | 0 | 0 | 0 | 0 | 0 | 0 | 0 | 0 | 0 |
| **Immune system disorders** | | | | | | | | | | | | |
| Guillain-Barré syndrome | 0 | 0 | 0 | 0 | 0 | 0 | 0 | 0 | 0 | 0 | 0 | 0 |

Abbreviations: E = number of AEs reported; IR = incidence rate is defined as number of events per 100 person-years = e/100 PY; MedDRA = Medical Dictionary for Regulatory Activities; NVX-CoV2373 = 5 µg SARS-CoV-2 rS with 50 µg Matrix-M™ adjuvant; SARS-CoV-2 = severe acute respiratory syndrome coronavirus 2; SARS-CoV-2 rS = severe acute respiratory syndrome coronavirus 2 recombinant spike protein nanoparticle vaccine.

#### References

1. Degli-Angeli E, Dragavon J, Huang M-L, et al. Validation and verification of the Abbott RealTime SARS-CoV-2 assay analytical and clinical performance. J Clin Virol 2020;129:104474. doi: 10.1016/j.jcv.2020.104474.
2. Skalina KA, Goldstein DY, Sulail J, et al. Extended storage of SARS-CoV-2 nasopharyngeal swabs does not negatively impact results of molecular-based testing across three clinical platforms. J Clin Pathol 2020 Nov 3;jclinpath-2020-206738. doi: 10.1136/jclinpath-2020-206738.
3. US Centers for Disease Control and Prevention. SARS-CoV-2 Variant Classifications and Definitions. (https://www.cdc.gov/coronavirus/2019-ncov/variants/variant-info.html).
4. Addetia A, Lin MJ, Peddu V, Roychoudhury P, Jerome KR, Greninger AL. Sensitive recovery of complete SARS-CoV-2 genomes from clinical samples by use of Swift Biosciences' SARS-CoV-2 Multiplex Amplicon Sequencing Panel. J Clin Microbiol 2020;59(1):e02226-20. doi: 10.1128/JCM.02226-20.
5. Zou G. A modified poisson regression approach to prospective studies with binary data. Am J Epidemiol 2004;159:702-6. (https://doi.org/10.1093/aje/kwh090).
6. US Food and Drug Administration. Guidance for Industry: Development and Licensure of Vaccines to Prevent COVID-19; June 2020. (https://www.fda.gov/media/139638/download).
7. Centers for Disease Control and Prevention. People with certain medical conditions. Updated May 13, 2021. (https://www.cdc.gov/coronavirus/2019-ncov/need-extra-precautions/people-with-medical-conditions.html).

8. US Food and Drug Administration, Center for Biologics Evaluation and Research (US). Guidance for Industry: Toxicity Grading Scale for Healthy Adult and Adolescent Volunteers Enrolled in Preventive Vaccine Clinical Trials. September 2007. (https://www.fda.gov/media/73679/download).
